# The cost-effectiveness of life after stroke services and the impact of these services on health and social care resource use: a rapid review

**DOI:** 10.1101/2024.11.21.24317699

**Authors:** Kalpa Pisavadia, Bethany Fern Anthony, Jacob Davies, Sofie Roberts, Rachel Granger, Llinos Haf Spencer, Elizabeth Gillen, Juliet Hounsome, Jayne Noyes, Deborah Fitzsimmons, Rhiannon Tudor Edwards, Adrian Edwards, Alison Cooper, Ruth Lewis

## Abstract

The UK is expected to see a 60% increase in first-time strokes over the next 20 years. This translates to about 2.1 million stroke survivors living in the UK by 2035. Life after stroke services aim to support peoples physical and emotional state, are complimentary to rehabilitation and take a non-medical holistic approach to living well after a stroke. This rapid review aimed to identify evidence on the cost-effectiveness of life after stroke services, and the impact of these interventions on health or social care resource use. The review included evidence from 12 studies (7 economic evaluations and 5 randomised controlled trials), published between January 2000 and August 2024.

The economic evaluations assessed a number of interventions to support stroke survivors, their families and caregivers. Two of the randomised controlled trials were partial economic evaluations, reporting on cost and resource use data related to training caregivers, and an arts and health-based intervention. The other three trials reported on resource use but not cost data, and assessed family support interventions, and a telehealth intervention to assist stroke survivors and their carers. There was a lack of evidence on the cost-effectiveness of life after stroke as a comprehensive service. However, this review identified evidence on the cost-effectiveness and resource utilisation of specific interventions within these services.

Findings include that a community-based Individual Management Program for post-stroke survivors was cost-effective from a societal perspective at 24-month follow-up. A carer training intervention, delivered whilst the stroke patient was in hospital, was associated with a reduction in health and social care resource use when evaluated at a single hospital. However, this intervention was not cost-effective when rolled out and assessed across multiple hospitals. Other findings included that a combination of speech and language therapy with voluntary support services had a lower cost per session compared to NHS speech and language therapy alone. Family support organiser interventions for stroke survivors and carers were associated with reduced healthcare utilisation.

To fully understand the effectiveness and cost-effectiveness of life after stroke interventions, research is needed to assess potential long-term impacts. A reduction in resource use may be associated with cost savings and reduced burden on the NHS. However, an increase in health care and social care use may also be appropriate due to better signposting or identification of peoples’ needs.

While traditional cost-effectiveness analysis provides a valuable framework for evaluating healthcare interventions, it may not fully capture the individual experiences of stroke survivors or the broader impact on their ongoing recovery, wellbeing and the lives of their carers. Future research should seek to quantify the effects of life after stroke support on patients and carers. Evidence is also needed on the cost-effectiveness of supporting stroke survivors in returning to the workforce. Many of the sample sizes in the included studies lacked ethnic diversity. Stroke trials need strategies to achieve equity of access. This review focused on evidence of cost-effectiveness and resource utilisation. Decisions relating to policy and practice should also consider evidence on clinical effectiveness and patient preferences.

**Funding Statement:** The authors were funded for this work by the Health and Care Research Wales Evidence Centre, itself funded by Health and Care Research Wales on behalf of Welsh Government.

**Report Contributors:** *Review Team:* Kalpa Pisavadia, Bethany Fern Anthony, Jacob Davies, Sofie Roberts, Rachel Granger, Llinos Haf Spencer, Elizabeth Gillen, Juliet Hounsome, Jayne Noyes, Deborah Fitzsimmons, and Rhiannon Tudor Edwards.

*Methodological Advice:* Ruth Lewis

*Evidence Centre Team:* Ruth Lewis, Adrian Edwards, Alison Cooper, Micaela Gal and Elizabeth Doe involved in Stakeholder engagement, review of report and editing.

*Public Partner:* Robert Hall

*Stakeholder(s):* Angela Contestabile, Heidi James, Lynn Preece, Llinos Wyn Parry, Katie Chappelle, Jonathan Hewitt. Evidence need submitted to the Evidence Centre: 30.10.2023 Initial Stakeholder Consultation Meeting: 29.04.24 Final Report issued: June 2025 The review should be cited as: Health and Care Research Wales Evidence Centre. A rapid review of cost-effectiveness of life after stroke services and the impact of these services on health and social care resource use_(RR0027). June 2025.

*Disclaimer:* The views expressed in this publication are those of the authors, not necessarily Health and Care Research Wales. The Health and Care Research Wales Evidence Centre and authors of this work declare that they have no conflict of interest.

**EXECUTIVE SUMMARY:** *What is a Rapid Review?:* Our rapid reviews use a variation of the systematic review approach, abbreviating or omitting some components to generate the evidence to inform stakeholders promptly whilst maintaining attention to bias.

*Who is this Rapid Review for?:* This Rapid Review was conducted as part of the Health and Care Research Wales Evidence Centre Work Programme. The review question was suggested by representatives of the Stroke Association UK.

*Background / Aim of Rapid Review:* The UK is expected to see a 60% increase in first-time strokes over the next 20 years. This translates to about 2.1 million stroke survivors living in the UK by 2035. Approximately 70,000 stroke survivors are currently living in Wales. Life after stroke services take a non-medical holistic approach to living well after a stroke and are complimentary to rehabilitation. Life after stroke services encompass services that aim to support people’s physical and emotional state. Some of these services are specific to communication and emotional support, providing tools and information, reassurance, coaching and peer support. Life after stroke services have the potential to reduce the cost burden to health and social care. This rapid review aimed to identify evidence on the cost-effectiveness of life after stroke services and the impact of these interventions on health or social care resource use (it did not review the evidence on the clinical effectiveness or peoples lived experience of life after stroke services).

*Results of the Rapid Review:* The evidence base:

- The review included evidence from studies published between January 2000 and August 2024 (at the time when the searches were conducted).
- Of the 12 studies included, seven were economic evaluations, and five were randomised controlled trials reporting on cost and or resource use data only.
- The seven economic evaluations assessed the following interventions: a needs assessment tool for caregivers (Patchwood et al, 2021); training for caregivers (Forster et al, 2013); an exercise and education reintegration programme for stroke survivors and their families (Harrington et al, 2010), a community-based Individual Management Program for post-stroke survivors (Orman et al, 2024); combined speech and language therapy and support services for stroke survivors and their families (van der Gaag and Brooks, 2008); peer-befriending for psychological distress (Flood et al, 2022), and a post-discharge structured assessment (Forster et al, 2015).
- Two randomised controlled trials were partial economic evaluations which reported on cost and resource use data. These studies specifically related to the following interventions: training caregivers (Kalra et al, 2004); and an arts and health-based intervention (Ellis-Hill et al, 2019).
- The remaining three randomised controlled trials reported on resource use but not cost data and assessed family support organiser service (Mant et al, 2000; Tilling et al, 2005), and a telehealth support intervention designed to assist stroke survivors and their carers (Bishop et al, 2014). Key findings:

- This review highlighted a lack of studies evaluating the cost-effectiveness of life after stroke as a comprehensive service as per the national stoke model, particularly when delivered as intended in a holistic multicomponent format. However, this review identified evidence on the cost-effectiveness and resource utilisation of specific interventions within these services, that support both stroke survivors and carers.
- A community-based Individual Management Program for post-stroke survivors was found to be cost-effective from a societal perspective at 24-month follow-up.
- A combination of speech and language therapy with voluntary support services had a lower cost per session than reported costs for equivalent NHS services However, cost-effectiveness was not assessed in this study
- A telehealth support intervention was associated with a reduction in healthcare utilisation.
- A caregiver training programme, delivered whilst the stroke patient was still in hospital, was associated with a reduction in health and social care resource use when evaluated at a single hospital. However, this intervention was not found to be cost-effective when rolled out and assessed across multiple hospitals.
- A community-based arts and health group intervention led to a reduction in outpatient contacts and home care worker contacts compared to usual care.
- A Family Support Organiser service was associated with a reduction in physiotherapy contact, but the use of other services were comparable with usual care.
- The addition of a new post-discharge system to an existing community-based Stroke Care Coordinators service was not cost-effective from either a health or societal perspective when compared to the Stroke Care Coordinator usual practice.
- Peer-befriending was found to be more costly and less effective than usual care alone.
- The use of a Carer Support Needs Assessment Tool (CSNAT) was not cost-effective compared to usual care.
- The cost per patient of a community-based combined exercise and education reintegration programme was greater than that of the control group.

*Policy and Practice Implications:*

- There is some evidence that interventions such as Individual Management Programme; telehealth support, community-based arts and health group, Family Support Organiser, and caregiver training can be cost-effective or lead to a reduction in service utilisation.
- While the gold standard model of traditional cost-effectiveness analysis proves a robust and valuable framework for assessing the cost-effectiveness of healthcare interventions, it does not capture the individual impact on the recovery of a stroke survivor and the ongoing health and wellbeing of them or their family.
- This review focused on evidence of cost-effectiveness and resource utilisation, which are helpful in the context of resource allocation for future roll-out of services, however, decisions relating to policy and practice must also consider the wider evidence base on clinical effectiveness and patient preferences going forward.
- Although a reduction in resource use may be associated with cost savings and reduced burden on the NHS, importantly, an increase in health care and social care use may also be appropriate due to better signposting or identification of peoples’ needs.

*Research Implications and Evidence Gaps:*

- Further research is needed on cost-effectiveness of life after stroke as a comprehensive service as per the national stoke model
- To fully understand the effectiveness and cost-effectiveness of life after stroke interventions, research is needed to assess potential long-term impacts.
- Evidence is needed on the cost-effectiveness of supporting stroke survivors in returning to the workforce.
- The timing of life after stroke interventions appeared to be important in terms of how they are received by stroke survivors, their carers and their families.
- Many of the samples in the included studies lacked ethnic diversity. Stroke trials need strategies to achieve equity of access, given that a large portion of UK stroke admissions are from Black, Asian and minority ethnic communities (Office for National Statistics, 2021).

*Economic considerations:*

- The average cost to society per stroke survivor in the first year post-stroke in the UK is £45,409. The key drivers of this cost are informal care and lost productivity costs (Stroke Association, 2020). Appropriate and timely post-stroke support for stroke survivors, their families and or caregivers could help alleviate some of this economic impact.
- Stroke costs NHS Wales £220 million per year. When considering a wider societal economic cost, this figure rises to £1.6 billion per year. If current trends persist with no action taken, this figure is forecast to increase to £2.8 billion per annum by 2035 (Welsh Government, 2024b).

**Glossary:** **Economic evaluation:** an assessment of the costs and effects of alternate healthcare interventions. The aim of an economic evaluation is to help decision makers maximise the level of health benefits relative to the finite resources available.

**Health and Social Care resource use or utilisation:** refers to the use of healthcare resources by end users (patients) and intervention deliverers. This can take the form of contacts with health professionals across health and social care, medicines or healthcare consumables used. In economic evaluations, resource use is collected, and costs are assigned to them to identify the costs an intervention places on the healthcare system.

**Cost-effectiveness analysis:** costs are compared with a treatment’s common therapeutic goal, expressed in terms of one main outcome measured in natural units (e.g., improvement in blood pressure or cholesterol level). These outcomes are typically condition-specific, meaning comparison within conditions is possible, but difficulty arises in comparing between conditions.

**Cost-utility analysis:** a method of evaluation that measures health benefits in preference-based, non-monetary units such as Quality Adjusted Life Years (QALYs) or Disability Adjusted Life Years (DALYs). These units are helpful for economic evaluation as they are generic and can be applied across conditions, allowing for comparison between conditions.

**Full economic evaluation:** is an economic evaluation that compares both the costs and effects of alternate healthcare interventions. The examples of cost-effectiveness and cost-utility above reflect full economic evaluations as they analyse both costs and outcomes of interventions.

**Partial economic evaluation:** is an economic evaluation that does not compare both the costs and effects of alternate healthcare interventions. A common partial economic evaluation is a cost-analysis that presents the costs of interventions only, with no consideration of effects. Partial economic evaluations are sometimes conducted within or alongside randomised controlled trials. However, the conduct of partial economic evaluations are not limited to randomised controlled trial study designs, with alternatives including economic evaluations alongside natural experiments or economic modelling studies which often utilise sources of data from previous literature in addition to or instead of collecting primary data.

**Randomised controlled trial:** a study in which a number of similar people are randomly assigned to 2 (or more) groups to test a specific drug, treatment or other intervention. One group (the intervention group) has the intervention being tested, the other (the comparison or control group) has an alternative intervention, a dummy intervention (placebo) or no intervention at all. The groups are followed up at set time periods to see how effective the experimental intervention was. Outcomes are measured at specific times and any difference in response between the groups is assessed statistically. Randomised controlled trials are the highest standard of research trials as their design helps to reduce biases that may impact the findings.

**Randomised controlled trials with partial economic evaluations:** are a study type that follows the randomised controlled trial methodology. Randomised controlled trials with cost comparisons collect information on the costs associated with the interventions studied to allow for comparison across alternate interventions not just based on clinical effect but also costs.

**Within-trial economic evaluation:** is a full economic evaluation of the costs and effects of healthcare interventions that are being studied as part of a clinical trial. The primary aim of clinical trials is typically a measurement of clinical effect of administering an intervention (e.g., change in blood pressure). These clinical effects of the intervention are then assessed together with the costs of the intervention in the economic evaluation.

**Cost-consequence analysis:** Is a form of partial economic evaluation that presents the disaggregated costs and outcomes of an intervention.

**Statistical significance:** a statistically significant result is one that is deemed to be down to a true effect rather than random chance. It is a way to determine if a relationship between variables is caused by something other than chance.

## 1. BACKGROUND

### 1.1 Who is this review for?

This Rapid Review was conducted as part of the Health and Care Research Wales Evidence Centre Work Programme. The research question was suggested by representatives of the Stroke Association UK in Wales.

The initial aim was to inform decision making by highlighting the cost and potential broader benefits of investing in life after stroke services. However, the findings of this review suggest that further research is needed before firm policy recommendations can be made. This review provides a foundation for future work and may still support early discussions around the economic value of post-stroke support services.

### 1.2 Background and purpose of this review

In the UK, more than 100,000 strokes occur each year, and this is expected to rise to over 2.1 million by 2035 (Stroke Association, 2023). In addition, there are approximately 70,000 stroke survivors living in Wales (Welsh Government, 2024b).

Strokes can vary significantly in their impact on individuals. Some strokes may result in mild symptoms, while others can lead to severe disability. The effects of a stroke depend on factors such as the location of the brain affected, the type of stroke (ischemic or haemorrhagic), and the promptness of medical intervention. Some common consequences of strokes are physical impairments (such as difficulty swallowing or speaking) and cognitive challenges (such as memory loss, concentration difficulties, and language impairments) (Donkor, 2018). Strokes can affect the activities of daily living of stroke survivors, including performing basic self-care tasks such as bathing, dressing, and eating (Lee et al, 2021).

Appropriate management and care are essential to prevent further strokes. Life after stroke services go beyond clinical interventions. Services may involve a multidisciplinary approach with social care and local authority provision. Additionally, third sector organisations offer a wider range of support, including initiatives such as care navigation, financial support, and social prescribing. Life after stroke services are non-medical and are complementary to rehabilitation with an emphasis on living well after a stroke. Life after stroke support aims to empower people to take an active part in their own recovery. Therefore, it addresses the holistic needs of an individual post-stroke through care navigation, advocacy, information and coaching that has a residual impact on people’s physical and emotional state. Life after stroke services provide targeted assistance in communication and emotional support, including tools, reassurance and peer support with others who have experienced stroke (Stroke Association, 2023). There are also broader services for carers of stroke survivors. Carers can find themselves in care roles overnight, which can have a detrimental effect on the whole family (Magwood et al, 2020).

The successful implementation of such services necessitates a focus on ensuring equitable access alongside robust evidence regarding their effectiveness and cost-effectiveness. Investing in services that improve the health and well-being of stroke survivors and their families will also have the potential to reduce the need for more costly and intense interventions further down the line.

This review will focus on the cost-effectiveness of life after stroke services, which support stroke survivors as they transition back to their daily lives after leaving the hospital. Post stroke services include both rehabilitation and life after stroke services. The remit of the review was on life after stroke services and not rehabilitation. Social care and community settings are included in this review, and non-medical interventions delivered in hospital and community-based settings and interventions delivered by health care professionals that do not include rehabilitation have also been considered. This review reports on evidence on the cost-effectiveness of life after stroke services from economic evaluations. The review also reports on cost comparisons and health or social care resource use from randomised controlled trials. This report did not focus on clinical effectiveness evidence of interventions. This report did not include evidence relating to views and preferences from qualitative data.

### 1.3 NICE guidance on life after stroke services

This section summarises guidance on life after stroke services that were included in the National Institute for Health and Care Excellence (NICE) 2023 guidance (NG236) on stroke rehabilitation in adults (NICE, 2023). The NICE guidance presented in this section will be revisited and considered in relation to the main findings of this rapid review later in the discussion section of this report (Section 3).

The NICE 2023 guidance (NG236) focuses on rehabilitation after a stroke for persons over the age of sixteen. It aims to ensure people are assessed for prevalent issues and conditions associated with stroke to allow them to get the care and therapy they need. It includes recommendations regarding the organisation and implementation of rehabilitation in hospital and community settings. Although this guidance predominantly focuses on clinically-led rehabilitation, some aspects consider the importance of life after stroke services that enhance rehabilitation. For example, one recommendation was the implementation of community participation programmes that encourage involvement in social activities. Such programmes focus on providing education, support or practice in areas such as participation in peer support groups, political or civic roles, leisure activities including exercise, art or music, participation in faith-based groups or organisations, walking or using other means of transport, such as buses, mobility scooters or taxis, employment or voluntary opportunities.

The nature of each community engagement programme and the extent of the involvement of health care professionals can vary. The findings of the NICE evidence review identified non-clinically led programmes such as group-based physical exercise and art and music activities that align with life after stroke services. Participants of these programmes typically reported that their quality of life improved, although the benefits they encountered differed among the various programmes used in the studies. Members of the NICE committee also had positive experiences of participating in these programmes and agreed they were of value to people post stroke. Therefore, the committee recommended that people could be referred to a community programme if there was one available which met their needs. They also agreed that family members and carers could find the programmes beneficial in preventing feelings of social isolation, improving quality of life and reducing caregiver strain.

NICE recommends that those requiring rehabilitation post-stroke should receive it from a specialised stroke service either within a stroke unit or in the community following early supported discharge. The types of support that NICE recommends are providing support and information to individuals post-stroke, along with their family members and carers. Reviewing a person’s information needs at their 6-month and annual stroke reviews, as well as at the start and end of any therapy, is recommended. Assessing care and support needs should encompass identifying the ongoing needs of the individual affected by stroke and their families and carers. NICE recommends training for family members and carers who are willing and able to be involved in supporting the individual after stroke. The training and support requirements of family members and caregivers should be assessed regularly, recognising that these needs may change over time.

Long-term health and social care support should encourage people to focus on life after stroke and help them achieve their goals, which may include daily activities and leisure pursuits, as well as reinforce their social roles, such as in employment, education, volunteering, and community participation programmes. A study in South Africa that focused on a return-to-work programme led by a physiotherapist and occupational therapist found important benefits to returning to work. Based on this evidence, NICE recommends that issues relating to return to work should be promptly identified following stroke and reviewed regularly by identifying the physical, cognitive, communicative and psychological requirements of the job. Any issues that impact work performance should be identified by conducting workplace visits in collaboration with employers to implement reasonable adjustments (NICE, 2023).

This rapid review aims to complement the NICE 2023 guidance (NG236) on adult stroke rehabilitation. This rapid review focuses on non-clinical life after stroke services and does not include studies of rehabilitation interventions delivered by health care professionals. We have also considered non-medical interventions delivered in hospital settings and interventions delivered by health care professionals. The rapid review findings reported in Section 2 will be considered in relation to the NICE guidance in the discussion section of this report (Section 3).

**Table 1:**
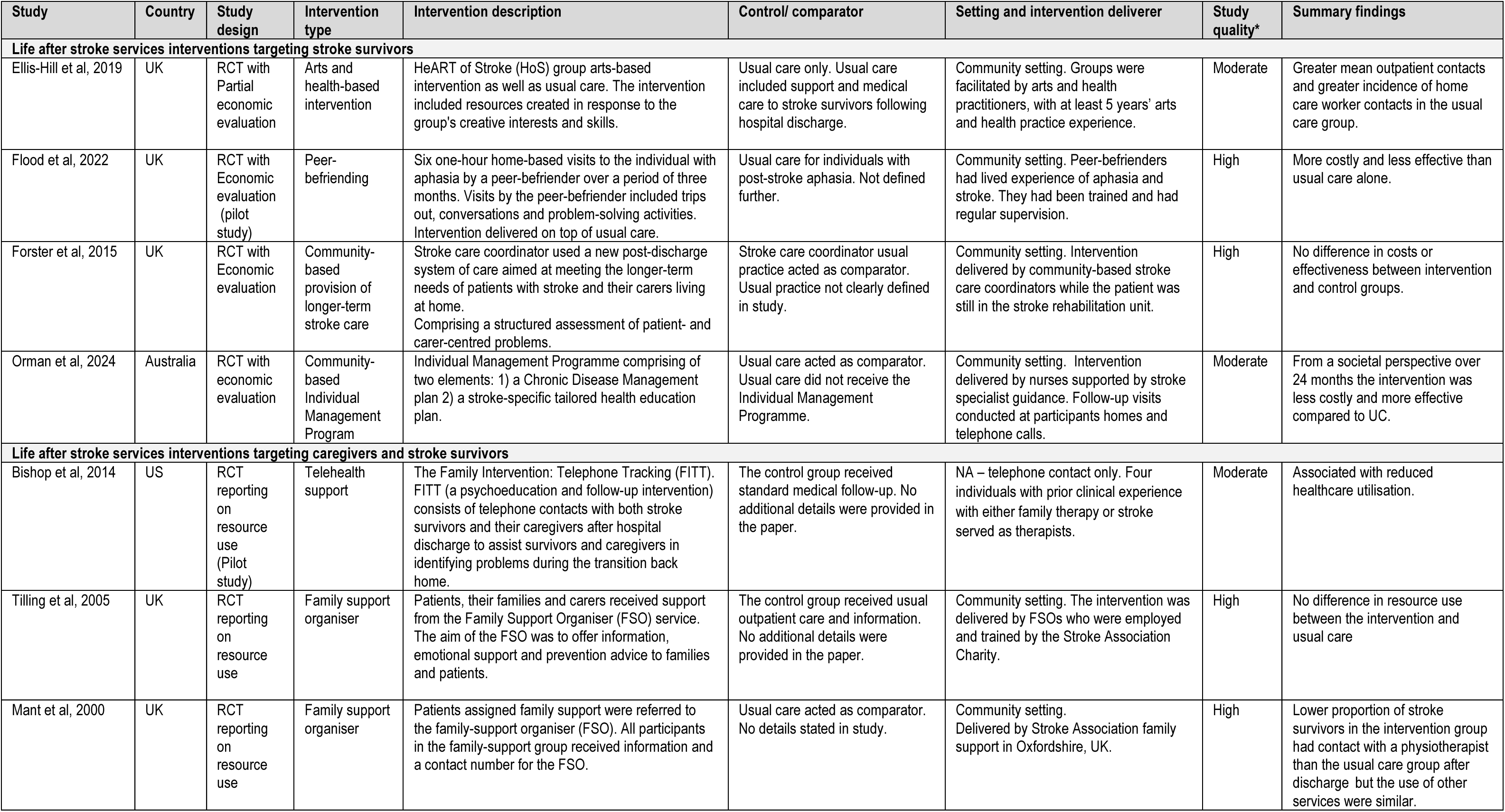
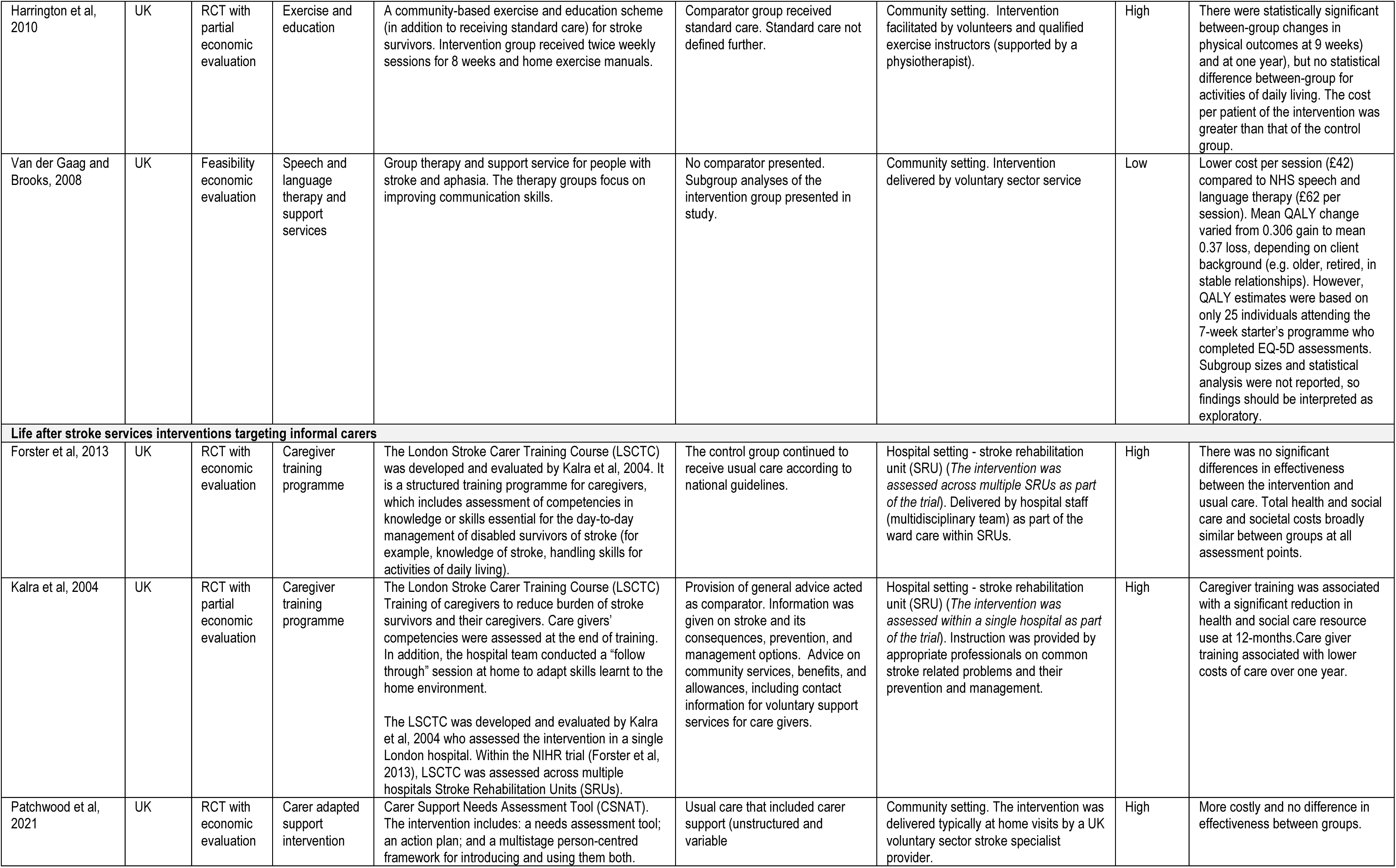
Study Characteristics.

| Study | Country | Study design | Intervention type | Intervention description | Control/ comparator | Setting and intervention deliverer | Study quality* | Summary findings |
| --- | --- | --- | --- | --- | --- | --- | --- | --- |
| **Life after stroke services interventions targeting stroke survivors** |  |  |  |  |  |  |  |  |
| Ellis-Hill et al, 2019 | UK | RCT with Partial economic evaluation | Arts and health-based intervention | HeART of Stroke (HoS) group arts-based intervention as well as usual care. The intervention included resources created in response to the group's creative interests and skills. | Usual care only. Usual care included support and medical care to stroke survivors following hospital discharge. | Community setting. Groups were facilitated by arts and health practitioners, with at least 5 years' arts and health practice experience. | Moderate | Greater mean outpatient contacts and greater incidence of home care worker contacts in the usual care group. |
| Flood et al, 2022 | UK | RCT with Economic evaluation (pilot study) | Peer-befriending | Six one-hour home-based visits to the individual with aphasia by a peer-befriender over a period of three months. Visits by the peer-befriender included trips out, conversations and problem-solving activities. Intervention delivered on top of usual care. | Usual care for individuals with post-stroke aphasia. Not defined further. | Community setting. Peer-befrienders had lived experience of aphasia and stroke. They had been trained and had regular supervision. | High | More costly and less effective than usual care alone. |
| Forster et al, 2015 | UK | RCT with Economic evaluation | Community-based provision of longer-term stroke care | Stroke care coordinator used a new post-discharge system of care aimed at meeting the longer-term needs of patients with stroke and their carers living at home. Comprising a structured assessment of patient- and carer-centred problems. | Stroke care coordinator usual practice acted as comparator. Usual practice not clearly defined in study. | Community setting. Intervention delivered by community-based stroke care coordinators while the patient was still in the stroke rehabilitation unit. | High | No difference in costs or effectiveness between intervention and control groups. |
| Orman et al, 2024 | Australia | RCT with economic evaluation | Community-based Individual Management Program | Individual Management Programme comprising of two elements: 1) a Chronic Disease Management plan 2) a stroke-specific tailored health education plan. | Usual care acted as comparator. Usual care did not receive the Individual Management Programme. | Community setting. Intervention delivered by nurses supported by stroke specialist guidance. Follow-up visits conducted at participants homes and telephone calls. | Moderate | From a societal perspective over 24 months the intervention was less costly and more effective compared to UC. |
| **Life after stroke services interventions targeting caregivers and stroke survivors** |  |  |  |  |  |  |  |  |
| Bishop et al, 2014 | US | RCT reporting on resource use (Pilot study) | Telehealth support | The Family Intervention: Telephone Tracking (FITT). FITT (a psychoeducation and follow-up intervention) consists of telephone contacts with both stroke survivors and their caregivers after hospital discharge to assist survivors and caregivers in identifying problems during the transition back home. | The control group received standard medical follow-up. No additional details were provided in the paper. | NA – telephone contact only. Four individuals with prior clinical experience with either family therapy or stroke served as therapists. | Moderate | Associated with reduced healthcare utilisation. |
| Tilling et al, 2005 | UK | RCT reporting on resource use | Family support organiser | Patients, their families and carers received support from the Family Support Organiser (FSO) service. The aim of the FSO was to offer information, emotional support and prevention advice to families and patients. | The control group received usual outpatient care and information. No additional details were provided in the paper. | Community setting. The intervention was delivered by FSOs who were employed and trained by the Stroke Association Charity. | High | No difference in resource use between the intervention and usual care |
| Mant et al, 2000 | UK | RCT reporting on resource use | Family support organiser | Patients assigned family support were referred to the family-support organiser (FSO). All participants in the family-support group received information and a contact number for the FSO. | Usual care acted as comparator. No details stated in study. | Community setting. Delivered by Stroke Association family support in Oxfordshire, UK. | High | Lower proportion of stroke survivors in the intervention group had contact with a physiotherapist than the usual care group after discharge but the use of other services were similar. |
| Harrington et al, 2010 | UK | RCT with partial economic evaluation | Exercise and education | A community-based exercise and education scheme (in addition to receiving standard care) for stroke survivors. Intervention group received twice weekly sessions for 8 weeks and home exercise manuals. | Comparator group received standard care. Standard care not defined further. | Community setting. Intervention facilitated by volunteers and qualified exercise instructors (supported by a physiotherapist). | High | There were statistically significant between-group changes in physical outcomes at 9 weeks) and at one year), but no statistical difference between-group for activities of daily living. The cost per patient of the intervention was greater than that of the control group. |
| Van der Gaag and Brooks, 2008 | UK | Feasibility economic evaluation | Speech and language therapy and support services | Group therapy and support service for people with stroke and aphasia. The therapy groups focus on improving communication skills. | No comparator presented. Subgroup analyses of the intervention group presented in study. | Community setting. Intervention delivered by voluntary sector service | Low | Lower cost per session (£42) compared to NHS speech and language therapy (£62 per session). Mean QALY change varied from 0.306 gain to mean 0.37 loss, depending on client background (e.g. older, retired, in stable relationships). However, QALY estimates were based on only 25 individuals attending the 7-week starter's programme who completed EQ-5D assessments. Subgroup sizes and statistical analysis were not reported, so findings should be interpreted as exploratory. |
| **Life after stroke services interventions targeting informal carers** |  |  |  |  |  |  |  |  |
| Forster et al, 2013 | UK | RCT with economic evaluation | Caregiver training programme | The London Stroke Carer Training Course (LSCTC) was developed and evaluated by Kalra et al, 2004. It is a structured training programme for caregivers, which includes assessment of competencies in knowledge or skills essential for the day-to-day management of disabled survivors of stroke (for example, knowledge of stroke, handling skills for activities of daily living). | The control group continued to receive usual care according to national guidelines. | Hospital setting - stroke rehabilitation unit (SRU) (*The intervention was assessed across multiple SRUs as part of the trial*). Delivered by hospital staff (multidisciplinary team) as part of the ward care within SRUs. | High | There was no significant differences in effectiveness between the intervention and usual care. Total health and social care and societal costs broadly similar between groups at all assessment points. |
| Kalra et al, 2004 | UK | RCT with partial economic evaluation | Caregiver training programme | The London Stroke Carer Training Course (LSCTC) Training of caregivers to reduce burden of stroke survivors and their caregivers. Care givers' competencies were assessed at the end of training. In addition, the hospital team conducted a "follow through" session at home to adapt skills learnt to the home environment.<br>The LSCTC was developed and evaluated by Kalra et al, 2004 who assessed the intervention in a single London hospital. Within the NIHR trial (Forster et al, 2013), LSCTC was assessed across multiple hospitals Stroke Rehabilitation Units (SRUs). | Provision of general advice acted as comparator. Information was given on stroke and its consequences, prevention, and management options. Advice on community services, benefits, and allowances, including contact information for voluntary support services for care givers. | Hospital setting - stroke rehabilitation unit (SRU) (*The intervention was assessed within a single hospital as part of the trial*). Instruction was provided by appropriate professionals on common stroke related problems and their prevention and management. | High | Caregiver training was associated with a significant reduction in health and social care resource use at 12-months. Care giver training associated with lower costs of care over one year. |
| Patchwood et al, 2021 | UK | RCT with economic evaluation | Carer adapted support intervention | Carer Support Needs Assessment Tool (CSNAT). The intervention includes: a needs assessment tool; an action plan; and a multistage person-centred framework for introducing and using them both. | Usual care that included carer support (unstructured and variable | Community setting. The intervention was delivered typically at home visits by a UK voluntary sector stroke specialist provider. | High | More costly and no difference in effectiveness between groups. |

## 2. RESULTS

### 2.1 Overview of the evidence base

Evidence of life after stroke service interventions aimed at stroke survivors only is presented in Section 2.2, interventions aimed at informal carers are presented in Section 2.3, followed by evidence of life after stroke services that support both informal carers and stroke survivors in Section 2.4. The evidence includes studies investigating the cost-effectiveness of life after-stroke services or studies that assess their impact on health or social care budgets directly or via their resource use.

A detailed summary of the eligibility criteria and the methods used for conducting the review are presented in Section 5. The rapid review search strategy is presented in Appendix 1. After the removal of duplicates, the database searches identified 7,364 references (see Figure 1 in Section 6.1 for the PRISMA diagram).

**Figure 1.**
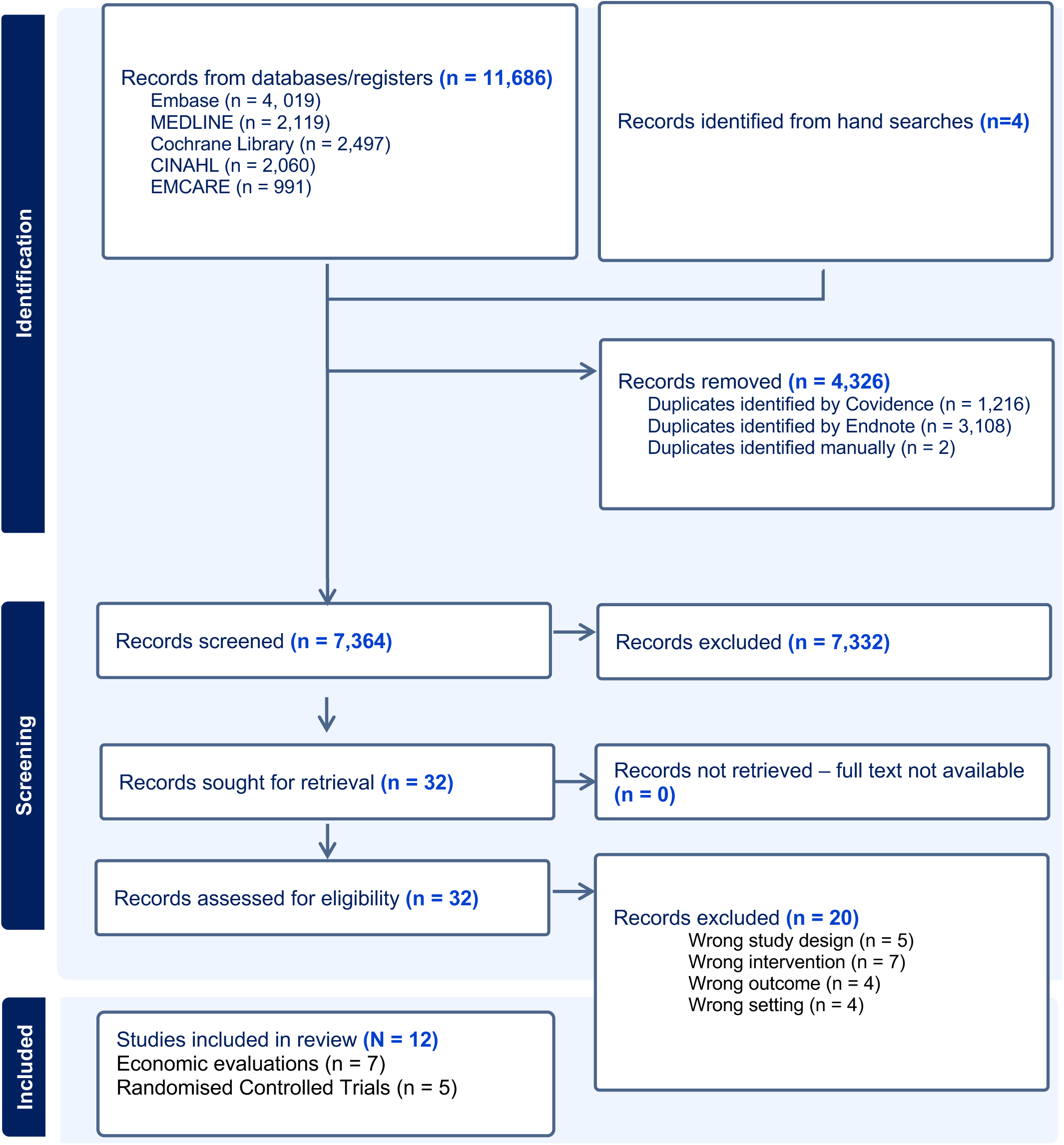
PRISMA 2020 flow diagram of included records (Page et al, 2021)

Following title and abstract screening, 32 papers were retrieved for full text screening. Twelve studies were included in this rapid review: seven economic evaluations and five randomised controlled trials that reported outcome data on health care or social care utilisation. Of these two randomised controlled trials reported comparative cost data. Four studies reported on interventions to support stroke survivors, which included peer-befriending (Flood et al, 2022), community-based provision of longer-term stroke care (Forster et al, 2015), an arts and health-based intervention (Ellis-Hill et al, 2019) and a community-based Individual Management Program (Orman et al, 2024). Three studies reported on interventions for informal carers. Of these, two studies evaluated carer education interventions, and one evaluated an intervention aimed as identifying the carer support needs upfront informal carers of stroke survivors (Kalra et al, 2004; Forster et al, 2013; Patchwood et al, 2021;). Five studies reported on interventions aimed at supporting stroke survivors and carers, which included speech and language therapy and support services (vaan der Gaag and Brooks, 2008), a telehealth support intervention (Bishop et al, 2014), a combined exercise and education intervention (Harrington et al, 2010), and two studies reported on family support interventions (Mant et al, 2000; Tilling et al, 2005).

Of the 12 studies included in this review, seven studies were economic evaluations which reported on:

- A needs assessment tool for caregivers (Patchwood et al, 2021);
- Training for caregivers (Forster et al, 2013);
- An exercise and education reintegration programme for stroke survivors and their Families (Harrington et al, 2010), and
- A community-based Individual Management Program for post-stroke survivors (Orman et al, 2024).
- therapy for stroke survivors and their families (van der Gaag and Brooks, 2008);
- peer-befriending for stroke survivors (Flood et al, 2022), and
- a post-discharge structured assessment (Forster et al, 2015).

Five studies included in this review were randomised controlled trials reporting on the resource use of life after-stroke services.

Two of these randomised controlled trials reported on comparative cost data specifically relating to:

- Training caregivers (Kalra et al, 2004); An arts and health-based intervention (Ellis-Hill et al, 2019), and Three of these randomised controlled trials did not report on comparative cost data, which specifically related to:
- Family support interventions (Mant et al, 2000; Tilling et al, 2005).
- A telehealth intervention designed to assist stroke survivors and their carers (Bishop et al, 2014).

### 2.2 Review of interventions to support stroke survivors only

Two high quality full economic evaluations, one high quality feasibility economic evaluation and one moderate quality randomised controlled trial are included in this rapid review focusing on interventions for stroke survivors only.

Flood and colleagues (2022) aimed to determine the feasibility of carrying out an economic evaluation of a peer-befriending intervention alongside standard care, as compared to standard care alone for individuals with post-stroke aphasia. The intervention involved six one-hour sessions with a peer-befriender who had experienced aphasia or stroke. These sessions consisted of outings, joint activities, conversations, and problem-solving activities. The analysis was conducted over a period of ten months and was undertaken from both NHS health and social care and societal perspectives. The costs were reported in British pound sterling for the cost year 2018/19 (Table 3 Section 6.2). Cost-effectiveness measured by the EQ-5D-5L VAS generated an Incremental Cost Effectiveness Ratio (ICER) of -£4,175. This indicates the intervention was more costly and less effective than usual care alone. Mapping of EQ-5D-5L data to EQ-5D-3L value sets generated an ICER of -£49,488. The ICER for utility, based on an improved change in mood (measured by the GHQ-12), was £373. Peer-befriending combined with usual care was found to be less effective than usual care alone, as measured by the EQ-5D-5L VAS. The mean health outcome gain was 5.19 for the intervention arm and 5.76 for usual care. There were no statistically significant differences in health and social care costs between the control and intervention groups after ten months, except for outpatient appointments, which were higher in the usual care group (p = 0.04). By calculating the average training and supervision costs, in addition to the cost of each befriender visit, the average cost was £57.24 per befriender visit (Flood et al, 2022).

In a prospective cost-effectiveness analysis undertaken by Forster and colleagues (2015), the objective was to evaluate the clinical and cost-effectiveness of a post-discharge stroke care system provided by community-based SCCs compared to the usual practice of SCCs. The post-discharge system of care comprised a structured assessment of long-term stroke problems (through sixteen structured assessment questions) and a care plan that included a goal and action planner. Following the initial post-discharge review, subsequent follow-up contacts were conducted to review assessment questions and goals. The number of contacts was determined by the SCCs’ usual practice and patient needs. The lack of clinical effectiveness restricted the cost-effectiveness analysis. At six months, the intervention had low probabilities of cost-effectiveness from both health and societal perspectives and for both outcomes (psychological well-being and functional outcomes for patient and carer) (Forster et al, 2015).

A randomised controlled trial by Ellis-Hill and colleagues (2019) aimed to evaluate the acceptability of ‘HeART of Stroke’, a community-based arts and health group intervention, to increase psychological well-being in stroke survivors. HeART of Stroke was facilitated by an experienced arts and health practitioner who prepared creative resources in response to group interests such as paints, drawing materials, clay, textiles and mixed media. ‘Stimulus’ pieces were offered, such as books, poems, images, music and films. Ten (two-hour) sessions were held over a 14-week period. The intervention was combined with usual care and compared to usual care only. Usual care included multidisciplinary medical care. After the ten sessions, individuals in the intervention group evidenced small QALY gains compared to usual care only (0.18 vs 0.17). The intervention would cost the health care payer, on average, £327 per participant in Bournemouth and £657 in Cambridgeshire (cost difference driven by increased venue hire costs in Cambridgeshire). The cost could be as low as £245 per participant at a full capacity of 8 people (in Bournemouth) (Ellis-Hill et al, 2019). Mean resource use contacts and associated costs were comparable across both groups. Differences between groups included greater mean outpatient physiotherapy contacts in the usual care group (0.3 vs 1; £6 vs £20); greater mean ‘other outpatient appointments’ in the usual care group (1.2 vs 2; £140 vs £196) and greater incidence of home care worker contacts in the usual care group (0 vs 0.9; £0 vs £4) (Ellis-Hill et al, 2019).

A within-trial cost-effectiveness analysis conducted by Orman and colleagues (2024) evaluated a comprehensive community-based Individual Management Program for post-stroke survivors. The programme consisted of a nurse-led chronic disease management plan and a tailored health education plan delivered at home by a nurse. Individuals in the intervention group received home visits at baseline, three months and 12 months, with telephone chronic disease management plan reviews at six and 18 months, in addition to usual care. Control group participants received usual care only. QALYs were estimated from utility scores obtained from the Assessment of Quality of Life 4-Dimension (AQoL-4D) questionnaire. These QALYs informed an ICER for intervention versus control, calculated at 12- and 24-months follow-up. There was no statistically significant difference in per-person QALYs between the groups within 12 months (β = 0.006, 95% CI: −0.051; 0.063) or 24 months (β = 0.031, 95% CI: −0.070; 0.133) after adjustment. ICERs at 12 months indicated the intervention was unlikely to be cost-effective from a health system perspective (AUD 136,363/QALY) or a wider societal perspective (AUD 87,027/QALY) (Orman et al, 2024). When re-calculated at 24 months follow-up, the ICER was AUD 53,175/QALY from a health system perspective, indicating a greater probability of being cost-effective. From a wider societal perspective at 24 months, the intervention was less costly and more effective than usual care with a mean cost of AUD 49,045 and 1.352 QALYs compared to AUD 51,394 and 1.324 QALYs in the usual care group. At 24 months follow-up, there was a 60.5% probability that the intervention was cost-effective from a societal perspective (Orman et al, 2024). At 24 months follow-up, average per-person costs from a societal perspective were greater for the usual care group (AUD 51,394, 95% CI: AUD 43,167; AUD 59,621) compared to the intervention (AUD 49,045, 95% CI: 39,127; AUD 58,962). From a health system perspective, the reverse was true. Average per-person costs were greater in the intervention group (AUD 21,707, 95% CI: AUD 16,929; AUD 26,485 for the intervention) than the usual care group (AUD 20,232, 95% CI: AUD 16,808; AUD 23,655) (Orman et al, 2024).

#### 2.2.1 Bottom line results of interventions to support stroke survivors only

Neither of the two economic evaluations focusing on interventions to support stroke survivors suggested the intervention could be cost-effective (Forster et al, 2015: Orman et al, 2024). In the peer-befriending intervention assessed by Flood and colleagues, the intervention was found to be more costly and less effective than usual care alone.

A post-discharge stroke care system assessed by Forster and colleagues (2015) had low probabilities of cost-effectiveness from both health and societal perspectives and for both outcomes (psychological well-being and functional outcomes for patient and carer).

The cost-effectiveness analysis conducted by Orman and colleagues (2024) suggested that a comprehensive community-based Individual Management Program for post-stroke survivors could be cost-effective from a societal perspective at 24-month follow-up. Outcomes measured in QALYs for individuals in the intervention group improved between 12 and 24-month follow-up, increasing the likelihood of being cost-effective compared to usual care.

The randomised controlled trial of a community-based arts and health group intervention by Ellis-Hill and colleagues (2019) found comparable health care resource use between the intervention and usual care groups; however, some differences in resource use costs between groups included greater mean outpatient contacts and greater incidence of home care worker contacts in the usual care group (Ellis-Hill et al, 2019).

### 2.3 Review of interventions for informal carers

Three UK based studies included in this review reported on interventions aimed at informal carers of stroke survivors. This included two high quality within-trial economic evaluations (Patchwood et al, 2021; Forster et al, 2013) and one high quality randomised controlled trial included health and social care resource use (Kalra et al, 2004).

The randomised controlled trial conducted by Kalra and colleagues assessed the impact of a caregiver training intervention on health and social care resource use at 12-months (Kalra et al, 2004). The London Stroke Carer Training Course (LSCTC) was developed and evaluated by Kalra et al, 200 who assessed the intervention in a single London hospital. Although the intervention was delivered within a hospital-based stroke unit, this was a non-medical intervention and may be comparable to life after stroke services that aim to enhance carer knowledge. The intervention involves carer support and training, which would eventually transition to a community setting. Carers in the intervention group received training from appropriate professionals from the stroke rehabilitation unit on prevalent stroke-related issues and methods for their prevention. The control group received carer support but no training. The provision of caregiver training was found to significantly reduce health and social care resource use costs at 12 months (£10,133 (SD: £8676) v £13,794 (SD: £10 510); P = 0.001), which was primarily due to lower hospital resource use costs (£1145 (SD £2553) v £1411 (SD £2742). Non-hospital costs, including community-based resource use costs, were similar for the caregiver training intervention and ‘no training’ groups; however, the use of personal, domestic and respite care services was less in the caregiver training group (Kalra et al., 2004). In addition, this training resulted in improved psychosocial outcomes for both carers and stroke survivors after one year, with carers reporting less anxiety (3 vs 4; p = 0.0001) and depression (2 vs 3; p = 0.0001) as well as better quality of life (80 vs 70; p = 0.001) (Kalra et al, 2004). The participants and those delivering the intervention in this trial were not blinded, which means that the group that they were allocated to (intervention or control) was not concealed and may have impacted their behaviour or interpretation of the outcome. The authors of this study acknowledged this as a study limitation, saying that there is a possibility that the measurement of the outcomes between groups and results of the study may be distorted (Kalra et al, 2004).

A within-trial cost-effectiveness analysis and cost-utility analysis conducted by Forster and colleagues (2013) aimed to evaluate the cost-effectiveness of a structured, competency-based training programme for caregivers. The LSCTC was the same intervention developed and evaluated by Kalra and colleagues, who assessed the intervention in a single London hospital. However, in this current economic evaluation, Forster and colleagues went on to assess the LSCTC across multiple hospitals. The intervention was delivered by community-based stroke care coordinators (SCC) while the patient was still in the stroke rehabilitation unit. The economic evaluation was undertaken from NHS health and social care and societal perspectives over a one-year time horizon. The costs that were assessed included health and social care services, as well as the cost associated with delivering the intervention, which included the development of the programme and staff training. The LSCTC was not determined to be cost-effective when compared to usual care due to a lack of clinical effectiveness and marginal differences in costs (Forster et al, 2013). Total health and social care and societal costs were broadly similar between groups at all assessment points.

However, caregivers in the intervention group had higher health and social care costs after six months, with an increase of £207 (95% CI: £5 to £408). The disparity ceased to exist after twelve months and was not evident when the costs from both evaluation periods were combined as one-year costs. The primary outcomes of the trial were functional independence for stroke survivors and caregiver burden. When compared to usual care, the LSCTC programme was found to not be as effective in improving stroke survivors’ long-term recovery or psychological well-being. The LSCTC was also not as effective as usual care in alleviating caregiver burden or improving their psychological well-being (Forster et al, 2013).

A trial-based cost-utility analysis conducted by Patchwood and colleagues (2021) aimed to evaluate the cost-effectiveness of the Carer Support Needs Assessment Tool (CSNAT) for stroke survivors. Although the intervention was practitioner-facilitated, it took a person-centred approach that enabled carers to identify and prioritise their unmet needs and participate in tailoring personalised support. The cost-utility analysis was undertaken from an NHS perspective over a one-year time horizon. The costs that were assessed included health and social care services, as well as the cost associated with delivering the intervention. These costs were reported in British pound sterling for the cost year 2017/18 (Table 3, Section 6.2). The analysis determined that the intervention is not likely to be cost-effective when compared to usual care. The net expenses amounted to £39.05, with a 95% confidence interval (CI) ranging from -£69.61 to £147.71. The difference in Quality-Adjusted Life Years (QALYs) was estimated to be −0.004, suggesting a marginal decrease in the quality of life for carers receiving the intervention. The 95% confidence interval for this QALY estimate ranged from −0.020 to 0.012 (Patchwood et al, 2021). There was no difference in resource utilisation between the intervention and usual care groups, apart from practice nurse contacts, which were used by a greater proportion of the intervention group, 43%, compared with 28% in the usual care group. Overall, mean costs for NHS resource use were marginally higher in the intervention group primarily due to staff training in the intervention arm and additional carer support, which amounted to 4.7 hours compared to 4.2 hours (Patchwood et al, 2021).

#### 2.3.1 Bottom line results for interventions to support informal caregivers

Two studies evaluated the same carer training intervention, which was a non-clinical intervention delivered by healthcare professionals in a hospital setting while the stroke survivor was an in-patient. In one study, the intervention was delivered in a single unit (Kalra et al, 2004) and then went on to be delivered across multiple stroke units in an economic evaluation (Forster et al, 2013).

Carer training is associated with a significant reduction in health and social care resource use at 12 months; this is primarily due to lower hospital resource use costs. Non-hospital costs, including community-based resource use costs, were similar for the caregiver training intervention and ‘no training’ groups; however, the use of personal, domestic and respite care services was less in the caregiver training group (Kalra et al., 2004).

Caregiver training immediately following a stroke was not cost-effective in the economic evaluation assessed by Forster and Colleagues (2013). The total health and social care and societal costs in this study were broadly similar between groups at all assessment points.

The Carer Support Needs Assessment Tool (CSNAT) for stroke assessed by Patchwood and colleagues (2021) was not cost-effective compared to usual care. The mean costs for NHS resource use were marginally higher in the intervention group in this study.

### 2.4 Review of interventions to support both informal caregivers and stroke survivors

Four studies reported on interventions aimed at both stroke survivors and carers. One feasibility economic evaluation (van der Gaag and Brooks 2008) and three high quality randomised controlled trials interventions providing family support for stroke survivors and informal carers were evaluated compared to usual care (Mant et al, 2000; Tilling et al, 2005; Bishop et al, 2014).

A within-trial cost-consequence analysis conducted by Harrington et al. (2010) aimed to improve the integration and well-being of stroke survivors and their families through a community-based intervention that combined exercise and education. The intervention was facilitated by volunteers and qualified exercise instructors and was supported by a physiotherapist. The primary outcome of this study was to evaluate physical improvement and the individual’s ability to reintegrate into a “normal” lifestyle. The economic analysis was undertaken from an NHS health and social care perspective and a societal perspective over a one-year time horizon. The costs assessed included health and social care services, the costs associated with delivering the intervention, and self-reported personal costs. The only significant difference between the intervention and those receiving usual care was greater physical improvement at nine weeks (p = 0.022) and one year (p = 0.024) reported in the intervention group. The intervention group also showed greater improvement at six months for the psychological domain of the WHOQol-bref questionnaire. The cost per participant in the intervention group was £746 (CI: -£432 to £1,924) greater than the usual care group.

The cost difference, not including patient care, was £296 (95% CI: -£321 to £913) (Harrington et al, 2010). This indicates that patient care was the most significant driver of cost differences between both groups.

van der Gaag and Brooks (2008) investigated the feasibility of undertaking a full economic appraisal of Connect, a voluntary sector service providing speech and language therapy and support services for individuals with aphasia and their families. Connect offers a 7-week community-based initiation programme called Starter’s, followed by a range of activities, including a women’s group, a group focused on improving communication skills, and a conversation group. The therapy groups aim to enhance communication skills and train individuals to teach volunteer service providers. Trained counsellors who personally experience aphasia also provided counselling services. The analysis was undertaken from a health care and societal perspective and was conducted over 18 months. These costs were reported in British pound sterling for the cost year 2002 (Table 3, Section 6.2). The Connect 7-week Starter’s Programme had a lower cost per session (£42) compared to NHS speech and language therapy (£62 per session). When considering the transportation costs for both programmes, the Connect programme was still less expensive at £55 per session compared to £74 (van der Gaag and Brooks, 2008). The average QALY gain for individuals receiving the intervention who were older, retired before experiencing a stroke, in stable relationships, and had good social support was 0.306. Individuals who remained working at the time of the stroke and expressed inadequate social support experienced a QALY loss of −0.37 (van der Gaag and Brooks, 2008). However, the QALY data from this study were generated from a small sample of 25 individuals on one of the programmes and lacked a control group, limiting the generalisability and robustness of the QALY estimates. Moreover, although QALYs were derived using EQ-5D, the reporting of subgroup sizes and statistical analysis was limited. As such, these results are best viewed as preliminary rather than conclusive evidence of the programme’s effectiveness across different demographic groups.

A pilot randomised controlled trial conducted by Bishop and colleagues (2014) aimed to test the efficacy of the Family Intervention: Telephone Tracking (FITT) intervention designed to assist stroke survivors and their caregivers during the first six months after stroke. FITT was delivered by individuals with prior clinical experience in family therapy or stroke (including a stroke rehabilitation nurse and therapists). During the six-month evaluation, an average of 19 telephone contacts were made with caregiver and survivor dyads. Participants in the intervention group received treatment as usual plus the FITT telephone contacts, while the control group received usual care only. Resource use was captured at three- and six-month follow-up for both groups. Resource use outcomes at six months indicated a trend of reduced therapy hours for the intervention group, suggesting the potential of FITT to reduce therapy time. 27% of survivors in the intervention group and 45% of survivors in the usual care group required rehospitalisation in the six months following stroke, which was suggestive but not statistically significant (χ2 (1, n = 49) = 1.57, p = 0.21). When considering rehospitalisation episodes, a large effect size for days re-hospitalised favoured FITT when analyses were undertaken using only those participants who experienced rehospitalisation, suggesting that when hospitalisation was necessary, hospital stays tended to be briefer for participants receiving the intervention (Bishop et al, 2014). Functional independence, depression and family functioning outcomes were assessed at three- and six-month follow-ups. At three months, caregivers in the FITT group had significant improvement in functional independence relative to caregivers in the usual care group, and this effect continued as a trend at six months. On average, caregivers in the FITT group reported improved activities of daily living scores, while those in the usual care groups worsened. Better family functioning was evidenced within caregiver/survivor dyads in the intervention group compared to usual care only. A non-significant positive change in depression outcome was also observed in the intervention group compared to usual care (Bishop et al, 2014).

A randomised controlled trial conducted by Mant and colleagues (2000) aimed to evaluate the impact of family support provided by the Stroke Association in Oxfordshire on stroke survivors and their caregivers. A family support organiser (FSO) was assigned to stroke survivors who were allocated to family support. The FSO used their discretion to determine the nature and frequency of interaction depending on the challenges and needs expressed by the families. Regarding health care resource use, the intervention led to a lower percentage of stroke survivors in the intervention group compared to the usual care group seeing a physiotherapist after being discharged (44% vs 56%, p = 0.04). In addition, stroke survivors in the intervention group used the Stroke Association stroke clubs more often and relied less on speech and language therapy compared to those in the usual care group (Mant et al, 2000). Carers in the intervention group demonstrated better outcomes compared to those in the usual care group in energy (p = 0.02), mental health (p = 0.004), pain (p = 0.03), physical function (p = 0.025), and general health perception (p = 0.02). Quality of life was significantly higher (p = 0.01), and they reported greater satisfaction with their understanding of stroke compared to the usual care group (83% vs 71%; p = 0.04).

However, this intervention did not provide any statistically significant impact on patient outcomes. There were no differences between the groups in terms of stroke survivors’ understanding of stroke, disability, handicap, quality of life, satisfaction with services, and knowledge about stroke. It is worth noting that the interviewer was not blinded to the allocation of participants in this study. Efforts were made in this study to explore the extent to which bias may have occurred, and it was found that there was a greater difference in the responses that were self-completed than those that were completed by the interviewer, which means that if bias had occurred, it did not favour the intervention.

A randomised controlled trial conducted by Tilling and colleagues (2005) aimed to evaluate the effectiveness of an FSO service for stroke survivors and their caregivers. The service provided FSO telephone consultations or in-person home visits with the patient, their caregiver, or both. FSOs received training from the Stroke Association, with the primary objective of providing information, emotional support, and preventive guidance to stroke survivors and their caregivers for post-stroke management. This support was designed to aid the transition from hospital to home and involved facilitating access to local statutory and voluntary services. Resource use data was captured after one year. A greater number of stroke survivors in the intervention group consulted their GP (p=0.08, 95% CI: –1, 20), but fewer visited the hospital due to stroke related issues (p=0.009, 95% CI: –30, –4). However, there was no significant difference in GP or hospital visits between the groups. The trial indicated that FSOs were not effective in improving the everyday impact in stroke survivors when compared to usual care and had no difference on resource use (Tilling et al, 2005). At three-month follow-up, stroke survivors in the intervention group self-reported poorer impact of their stroke on daily life compared to those receiving usual care (6 [intervention] v 7 [usual care]; 95% CI: –1.7, +0.01; p = 0.05). A greater proportion of stroke survivors in the intervention group reported that stroke continued to adversely affect their lives compared to those receiving usual care. However, stroke survivors in the intervention group reported higher satisfaction with the information they received regarding their stroke recovery [76 (71%) intervention, 53 (49%) usual care; p = 0.001] and preventative advice [58 (54%) intervention, 46 (42%) usual care; p = 0.09] compared to the usual care group (Tilling et al, 2005). Within the intervention conducted by Tilling and colleagues (2005), the participants were aware of whether they were allocated to the intervention or usual outpatient care (not blinded). The authors of this study did not explore the extent to which this may have resulted in bias.

#### 2.4.1 Bottom line results for interventions to support both informal carers and stroke survivors

There is a lack of evidence on the cost-effectiveness of interventions to support informal carers and stroke survivors. However, two partial economic evaluations reported on comparative costs (Harrington et al, 2010; van der Gaag and Brookes, 2008) and three studies reported on resource use but did not provide cost comparisons (Mant et al, 2000; Tilling et al, 2005; Bishop et al, 2014).

The community-based intervention that combined exercise and education, evaluated by Harrington and colleagues (2010), reported that the cost per patient of the intervention was greater than that of the control group (the mean intervention cost was £746 higher than the mean control group).

The feasibility economic analysis of speech and language therapy and support services assessed by van der Gaag and Brooks (2008). The Connect 7-week Starter’s Programme had a lower cost per session (£42) compared to NHS speech and language therapy (£62 per session). Three randomised controlled trials to support both informal carers and stroke survivors did not include comparative cost data.

A randomised controlled trial of a telehealth intervention to support families, assessed by Bishop and colleagues (2014), was associated with decreased health care utilisation.

In the randomised controlled trial conducted by Mant and colleagues (2000) assessing family support, a lower proportion of stroke survivors in the intervention group had contact with a physiotherapist than the usual care group after discharge. In another randomised controlled trial assessing family support organisers conducted by Tilling and colleagues (2005), the findings reported no difference in resource use between the intervention and usual care.

## 3. DISCUSSION

### 3.1 Summary of the findings

The primary focus of this rapid review was to identify evidence on the cost-effectiveness of life after stroke services that provide community-based and non-medical holistic support. This review also reported on the health and social care resource use and costs of life after stroke services. This review did not focus on rehabilitation. However, it is acknowledged that there is often an overlap in the boundary between life after stroke services and rehabilitation. This review did not include clinical interventions delivered in health care settings. However, non-clinical interventions delivered by health care professionals in clinical settings which may be relevant to life after stroke services were considered.

This review has highlighted a need for more research evidence on the cost-effectiveness of comprehensive life-after-stroke services, particularly when they are intended to be delivered in a holistic multi-component format. However, this review has identified evidence on the cost-effectiveness and resource utilisation of specific interventions within these services that support both stroke survivors and carers.

We identified four studies assessing interventions targeting stroke survivors, but did not identify any evidence supporting their cost-effectiveness. Two trial-based economic evaluations found that community-based provision of longer-term stroke care (Forster et al, 2015) and a community-based individual management programme (Orman et al, 2024) were not cost-effective relative to usual care. However, as the follow-up period increased the probably of the programme being cost-effectiveness increased. Moreover, a trial-based economic evaluation of a peer-befriending intervention for stroke survivors was found to be more costly and less effective than usual care alone (Flood et al, 2022). The fourth study, a randomised controlled trial of an arts and health-based intervention for stroke survivors reported comparable health care resource use between the intervention and usual care groups; however, some differences in resource use costs between groups included greater mean outpatient contacts and greater incidence of home care worker contacts in the usual care group (Ellis-Hill et al, 2019).

Three studies that assessed training interventions for carers were identified in this rapid review. Two studies evaluated the same carer training intervention, which was a non-clinical intervention delivered by healthcare professionals in a hospital setting, while the stroke survivor was an in-patient. In one study, the intervention was delivered in a single unit (Kalra et al, 2004) and then went on to be delivered across multiple stroke units in an economic evaluation (Forster et al, 2013). Carer training was associated with a significant reduction in health and social care resource use at 12-months (Kalra et al, 2004), but was not found to be cost-effective in the separate study assessing the same intervention across multiple sites (Forster et al, 2013). In the third study of carer training, a carer adapted support intervention, which included a Carer Support Need Assessment Tool (CSNAT), was not cost-effective compared to usual care (Patchwood et al, 2021). Moreover, the mean costs for NHS resource use were marginally higher in the intervention group in this study.

This review identified five studies assessing interventions targeting both stroke survivors and their informal carers, but none of these studies provided evidence relating to their cost-effectiveness. Two partial economic evaluations presented comparative costs; the first was a combined exercise and education intervention and reported higher mean delivery costs for the intervention group compared to the usual care group (Harrington et al., 2010). The second partial economic evaluation assessed speech and language therapy and a support service intervention delivered by a third-sector voluntary organisation, and reported a lower cost per session for the intervention compared to an NHS-delivered speech and language therapy session (van der Gaag et al., 2008). The reported subgroup QALY differences in this small feasibility study lacked detailed subgroup data or statistical analysis meaning these findings should be treated as exploratory rather than conclusive.

The remaining three studies of interventions to support stroke survivors and their informal carers reported on resource utilisation, but did not provide any cost data as part of their analysis. A randomised controlled trial of a telehealth intervention to support families, assessed by Bishop and colleagues (2014), was associated with decreased health care utilisation. A randomised controlled trial of a family support intervention reported that a lower proportion of stroke survivors in the intervention group had physiotherapist contacts when compared to the usual care group following hospital discharge (Mant et al, 2000). In a separate randomised controlled trial assessing a family support organiser intervention, the results indicated no difference in resource use between the intervention and usual care groups (Tilling and colleagues, 2005).

#### Rapid review findings in relation to NICE guidance

This current rapid review aims to complement the NICE 2023 guidance (NG236) on adult stroke rehabilitation. However, this rapid review focuses on non-clinical life after stroke services and does not include studies of rehabilitation interventions delivered by health care professionals. We have also considered non-medical interventions delivered in hospital settings and interventions delivered by health care professionals. The rapid review findings are considered in relation to the NICE guidance in the following sections, which highlight community participation programmes, assessment of care and support needs, carer training, telerehabilitation, community-based communication and support groups, and care in the community and early hospital discharge.

#### Community participation programmes

NICE guidance indicates that community participation programmes that include peer support and leisure activities have been evidenced to be of value to people post-stroke. NICE also advocates that the involvement of family members in these programmes can be beneficial in improving quality of life and caregiver strain (NICE, 2023). This rapid review identified two economic evaluation studies and one randomised controlled trial assessing community participation programmes. The peer-befriending intervention assessed by Flood and colleagues was found to be more costly and less effective than usual care alone (Flood et al, 2022). There were some physical improvement and psychological outcomes within the community exercise and education scheme, assessed by Harrington and colleagues. However, the cost per patient of the intervention was greater than that of the usual care group (Harrington et al, 2010). The community-based arts and health group intervention assessed by Ellis-Hill and colleagues led to minimal QALY gain when compared to usual care, and there were no significant changes in health care resource use (Ellis-Hill et al, 2019).

#### Assessment of care and support needs, and carer training

NICE recommends appropriate assessment of care and support needs, which includes training in care to family members and carers who are willing and able to be involved in supporting the person after stroke. Family and carer training and support needs should be reviewed at a minimum during the person’s six-month and annual reviews (NICE, 2023). This rapid review identified three studies focussing on carer support needs and training. One study providing training to caregivers during stroke survivors’ rehabilitation was found to significantly reduce health care costs and caregiver burden as well as improve psychosocial outcomes for both caregivers and stroke survivors (Kalra et al, 2004). However, the economic evaluation assessing the same intervention across multiple sites (Forster et al, 2013) contradicted the results of Kalra and colleagues, as carer training in this study did not reduce health care utilisation and was also not found to be cost-effective. Forster and colleagues conclude that caregiver training delivered in the immediate post-stroke period may not be as effective as being delivered after discharge by community-based teams. The Carer Support Needs Assessment Tool, which was assessed by Patchwood and colleagues, was not cost-effective and did not evidence any improvement in carer burden. Additionally, the mean costs for NHS resource use were marginally higher in the intervention group within this study (Patchwood et al, 2021).

NICE (2023) recommends that long-term health and social support should include the provision of information so that people after a stroke, as well as their families and carers, can recognise the complications of the condition. This review identified two randomised controlled trials evaluating the impact of Family Support Officers on stroke survivors and their carers. The study conducted by Mant and colleagues (2000), demonstrated better outcomes overall for carers with significantly higher quality of life and greater satisfaction with their understanding of stroke compared to the usual care group. In contrast, in the randomised controlled trial by Tilling and colleagues (2005), family support organisers were not effective in improving the everyday impact on stroke survivors when compared to usual care.

Regarding resource use, a lower proportion of stroke survivors in the family support group had contact with a physiotherapist than the usual care group after discharge in the study conducted by Mant and colleagues (2000). However, family support organisers in the study conducted by Tilling and colleagues (2005), demonstrated no meaningful effect on GP visits and stroke related hospital visits.

#### Telerehabilitation

NICE (2023) recommends that post-stroke telerehabilitation be delivered as an alternative to face-to-face interaction with a health care professional. Components can include interventions, supervision, education, consultations and counselling. This review identified a randomised controlled trial evaluating the Telephone Tracking (FITT) intervention designed to assist stroke survivors and their caregivers during the first six months after stroke (Bishop et al, 2014). This intervention evidenced the potential to decrease health care utilisation and improve the quality of life for stroke survivors and their caregivers.

#### Community-based communication and support groups

NICE (2023) recommends encouraging people with communication difficulties to participate in community-based communication and support groups (such as those provided by voluntary organisations). This review identified one study evaluating a community-based speech and language therapy programme (van der Gaag & Brookes, 2008). Although the intervention was less costly per session compared to its NHS equivalent of speech and language therapy, the intervention evidenced mixed results on its effectiveness. Those who were retired prior to experiencing stroke evidenced greater quality of life than working-age individuals post-intervention.

#### Care in the community and early hospital discharge

Based on a NICE (2023) evidence review, the committee agreed that transfer of care from hospital to community, including early supported discharge, had clinically important benefits of reducing physical dependency, improved quality of life or had no negative impact on it, and reduced psychological distress. Qualitative evidence also found that people after stroke, their families, carers, and health care professionals saw early supported discharge as beneficial. This review found one intervention that evaluated a community-based individual management programme. The cost-effectiveness analysis suggested that the programme could be cost-effective from a societal perspective at 24-month follow-up. Outcomes measured in QALYs for individuals in the intervention group improved between 12 and 24-month follow-up, increasing the likelihood of being cost-effective compared to usual care. (Orman et al, 2024).

#### Assessments of psychological well-being and functional outcomes

NICE (2023) guidance indicates that the assessment of psychological functioning should be included within the six-month annual reviews and the referral of stroke survivors to appropriate services for assessment and treatment when changes in psychological functioning are identified. This review identified one study assessing psychological well-being and functional outcomes for stroke survivors and carers (Forster et al, 2015). This study had low probabilities of cost-effectiveness from both health and societal perspectives and for both outcomes.

### 3.2 Strengths and limitations of the available evidence

This review identified seven economic evaluations of life after stroke services, of which five were deemed to be of high quality. In addition, five randomised controlled trials were identified evaluating the impact of life after stroke services on health and social care resource use, three of which were deemed to be of high quality. Two of the identified randomised controlled trials reported comparative cost data.

Many of these studies were feasibility studies that did not carry out a full evaluation or evaluated across small sample sizes, which limits confidence in the results.

Very few of the identified studies evidenced the clinical effectiveness of life after-stroke interventions. The lack of clinical effectiveness in some studies hindered the ability to interpret cost-effectiveness analysis.

Clinical effectiveness was typically assessed over short-term time horizons in most of the identified studies, with a follow-up period of less than a year. The NICE manual for conducting health technology evaluations states that a time horizon long enough to reflect all important differences in costs or outcomes should be used (NICE, 2022). Given the potential for long-term impacts of life after stroke services, short-time horizons may not provide us with a full understanding of health outcomes or resource utilisation and costs. This is evidenced in the economic evaluation conducted by Orman and colleagues (2024) in which significant improvements to QALY were found between 12 and 24-month follow-up. A shorter time-horizon would not have identified these gains. NICE recommends return to work support for stroke survivors. This includes identifying any issues that may impact work performance and conducting workplace visits in collaboration with employers to implement reasonable adjustments (NICE, 2023). However, this review did not identify any evidence on the cost-effectiveness of return-to-work support for stroke survivors.

### 3.3 Strengths and limitations of this rapid review

This rapid review undertook thorough literature searches of evidence from January 2000 to August 2024, using a well-developed search strategy and robust methodology. We aimed to capture studies from comparative countries to allow the generalisability of findings to the UK health and care system. The searches aimed to identify evidence on the cost-effectiveness of life after stroke services, which support stroke survivors as they transition back to their daily lives after leaving the hospital, and the impact of these services on health and social care resource use.

As this review focused on the cost-effectiveness of life after stroke services and the impact on health and social care resource use, there may be a considerable amount of evidence on non-medical interventions that are effective but have not been economically evaluated that were not considered for inclusion in this review.

This review included interventions targeting not only stroke survivors but also carer/stroke survivor dyads and carers themselves, which gives a comprehensive overview of the available literature on life after stroke services.

The search strategy of this review covered a wide time period; however, half of the included studies were published in the last 10 years (2014 onwards). The relevancy and generalisability of such dated findings mean that studies were conducted in significantly different settings than the present day in which policy and practice have changed.

### 3.4 Implications for policy and practice

- While this review focused on evidence of cost-effectiveness and resource utilisation, which are helpful in the context of resource allocation for future roll-out of services, decisions relating to policy and practice should also consider the wider evidence base on clinical effectiveness and patient preferences going forward.
- Community-based individual management programmes for post-stroke survivors may have the potential to be more cost-effective than usual care.
- Life after stroke services may prevent the need for more costly and invasive treatments in the future.
- The reduction in resource use for family support for stroke survivors and their carers may be associated with cost savings and reduced burden on the NHS. However, an increase in health care and social care use may also be appropriate due to better signposting or identification of peoples’ needs.
- There is a need for adequate assessment of family and caregiver support needs to identify and address unmet needs.

### 3.5 Implications for future research

- As stroke survivors have a diverse range of unmet needs, targeted, bespoke interventions may be the best way to provide holistic, non-medical life after stroke care. Research into the effectiveness and cost-effectiveness of bespoke life after stroke services should be conducted to evidence this claim.
- Many of the samples in the included studies lacked ethnic diversity, which does not reflect the diversity of the UK population. Stroke trials need strategies to achieve equity of access, given that a large portion of UK stroke admissions are from Black, Asian and minority ethnic communities (Office for National Statistics, 2021; Raleigh, 2023; Fluck et al, 2023).
- To fully understand the cost-effectiveness of life after stroke interventions, future research should adopt longer study time horizons to allow for the assessment of potential long-term impacts. Exploration of outcomes over longer time horizons will enable researchers to gain valuable insights into any sustained benefits of interventions. The use of model-based economic analyses can facilitate longer time horizons, beyond that typically used in clinical trials, through the extrapolation of cost and outcomes data.
- While the gold standard model of traditional cost-effectiveness analysis provides a valuable framework for assessing the cost-effectiveness of health care interventions, it may not capture the broader, softer benefits of life after stroke services, including interventions recommended by NICE. Qualitative interviews and focus groups can shed light on these softer outcomes, such as reduced social isolation, independence, and increased participation in daily activities. While this rapid review focused on quantitative data, future research may benefit from incorporating qualitative evidence to provide a more comprehensive understanding of the value of these interventions.

#### Economic considerations

- The average cost to society per stroke survivor in the first-year post-stroke in the UK is £45,409. The key drivers of this cost are informal care and lost productivity costs (Stroke Association, 2020). Appropriate and timely post-stroke support for stroke survivors, their families and or caregivers could help alleviate some of this economic impact.
- Stroke costs NHS Wales £220 million per year. When considering a wider societal economic cost, this figure rises to £1.6 billion per year. If current trends persist with no action taken, this figure is forecast to increase to £2.8 billion per annum by 2035 (Welsh Government, 2024a).

## Abbreviations

Acronym: Full Description
ADL: Activities of Daily Living
AQoL-4D: Assessment of Quality of Life-Four Dimensions
AUD: Australian Dollars
CCA: Cost-Consequence Analysis
CDM: Chronic Disease Management
CI: Confidence Interval
CSNAT: Carer Support Needs Assessment
CSRI: Client Service Receipt Inventory
CUA: Cost Utility Analysis
DISCS: Depression Intensity Scale Circles
EQ-5D-5L: EuroQol-5 Dimensions-5 Levels
FAD: Family Assessment Device
FACQ: Family Appraisal of Caregiving Questionnaire
FAI: Frenchay Activities Index
FITT: Family Intervention: Telephone Tracking
FSO: Family Support Organiser
GDS: Geriatric Depression Scale
GHQ-12: General Health Questionnaire-12
HADS: Hospital Anxiety and Depression Scale
HISDS-III: Head Injury Semantic Differential Scale
HRQoL: Health Related Quality of Life
ICER: Incremental Cost Effectiveness Ratio
IMP: Individual Management Program
JBI: Joanna Briggs Institute
LAS: Life After Stroke
LOS: Length of Stay
LSCTC: London Stroke Carer Training Course
NEADL: Nottingham Extended Activities of Daily Living
NICE: National Institute for Health and Care Excellence
OECD: Organisation for Economic Co-operation and Development
PCS: Perceived Criticism Scale
QALY: Quality Adjusted Life Year
RCT: Randomised Controlled Trial
RNLI: Reintegration to Normal Living Index
RSES: Rosenberg Self-Esteem Scale
SCC: Stroke Care Coordinator
SD: Standard Deviation
SE: Standard Error
SIPSO: Subjective Index of Physical and Social Outcome
SIS: Stroke Impact Scale
SLT: Speech and Language Therapy
SMF: Standard Medical Follow-up
SRU: Stroke Rehabilitation Unit
TIA: Transient Ischaemic Attack
UC: Usual Care
VAS: Visual Analogue Scale
WEMWBS: Warwick-Edinburgh Mental Well-being Scale
WTP: Willingness to Pay

## Data Availability

All data produced in the present study are available upon reasonable request to the authors

## RAPID REVIEW METHODS

### 5.1

**Table 2:** Eligibility criteria.

|  | Inclusion criteria | Exclusion criteria |
| --- | --- | --- |
| **Participants** | Stroke survivors and their families/carers | People who have other conditions (e.g., cardio-vascular disease) that mimic stroke. The use of rehabilitation service. |
| **Settings** | Social care and community settings.<br>Non-medical interventions delivered in hospital and community-based settings may be considered. | Hospital and community-based stroke rehabilitation and medical treatment. |
| **Intervention / exposure** | Life after stroke (LAS) services include community based, holistic and non-medical support for as long as needed after stroke. Provision is a mix of home visits and telephone support. The core principles are:<br>- Personalised support according to need – e.g., one off advice vs ongoing 1:1 keyworker style support.<br>- LAS addresses holistic needs through care navigation, advocacy, information and coaching.<br>All services that were identified from the Integrated Life After Stroke Support (ILASS) report will also be included such as:<br>- Telephone services<br>- Support services<br>- Befriending<br>- Wider social care or local authority funded services<br>- Linking up services<br>- Financial support services<br>- Return to work initiatives<br>- Carer/family support services<br>- Post-stroke reviews<br>- Information provision<br>- Active listening and emotional wellbeing support<br>- Lifestyle support including educational approaches<br>- Secondary prevention support<br>- Communication support for people with ongoing communication difficulties<br>- Peer support<br>N.B. Relevant non-clinical services may be clinically led or provided/funded by clinical organisations (e.g., health boards in Wales), local authorities or third sector organisations, multidisciplinary teams, third sector/voluntary organisations, care navigators or coordinators. |  |
| **Comparison** | Any comparator e.g., alternative services, charities, usual care or no intervention comparator. |  |
| **Outcomes** | Cost-effectiveness outcomes (e.g., cost per QALY).<br>Costs: cost savings for NHS/social care, return on investment and social return on investment, societal costs (e.g., informal care costs, productivity losses/gains), third sector costs.<br>Resource utilisation: health care or social care resource use. |  |
| **Study design** | Full economic evaluations: (cost-effectiveness analyses, cost-utility analyses, cost-benefit analyses, cost-minimisation analyses).<br>Partial economic evaluations: Return on Investment (ROI) analysis, Social Return on Investment (SROI) analysis.<br>RCTs reporting outcome data on health care or social care resource use. |  |
| **Countries** | A focus on UK and European literature with a view of broadening to OECD countries. | Non-OECD countries |
| **Language of publication** | English | Full text publications not available in the English language |
| **Publication date** | 2000 | Studies published prior to 2000 |
| **Publication type** | Published and preprint |  |

### 5.2 Literature search

A two-step approach was adopted to search the literature. Each search focused on terms to describe services following a stroke combined with different validated filters to limit the search numbers and to increase sensitivity. Search 1: limited the search by geographical area using a validated UK filter (Ayiku et al, 2021). Search 2: limited the search by study type using a combination of economic filters (CADTH, 2024a; CADTH, 2024b) and geographical area using an OECD filter (Ayiku et al, 2021). Searches were conducted within the period 2000 to August 2024 in the following databases: Medline (Ovid), EMBASE (Ovid), CINAHL (EBSCO), EMCARE (Ovid) and Cochrane Library. Search strategies 1 and 2 conducted in Medline via Ovid are presented in Appendix 1.

### 5.3 Study selection process

Three reviewers independently screened 50% each of the titles and abstracts using the Covidence review management software. Two reviewers independently screened 100% of the full text articles. Following the independent full text screening stage, discrepancies were resolved through discussion with the review team to come to an agreement on the final inclusions.

### 5.4 Data extraction

Data extraction was based on the outlined eligibility criteria. For the economic evaluation studies, the review team extracted data on study country, type of intervention, study design, sample size, length of follow-up, type of economic evaluation, perspective of analysis, currency and cost year, details of discounting and sensitivity analysis, main costs and outcomes measures, and main health economics findings. For randomised control trials, the review team extracted data of study country, type of intervention, study design, sample size, participants and settings, intervention and comparator/control, outcomes that are relevant to the review question and main outcomes of the trial.

### 5.5 Quality appraisal

Economic evaluation studies were assessed using the JBI critical appraisal checklist for economic evaluations, and the randomised controlled trials were assessed using the JBI Checklist for randomised controlled trials (Joanna Briggs Institute, 2017b).

For the economic evaluation studies and the randomised controlled trials, the scoring algorithm employed by the authors awarded a single point to any element marked Y or NA while awarding no point to any element marked U or N. These points were totalled out of 11, and quality cut-offs were created to categorise the evidence into quality levels. The following cut-off scores for the economic evaluations are as follows: 9 to 11 – High Quality, 5 to 8 – Moderate, and 0 to 4 – Low Quality. The cut-off scores for randomised controlled trials were as follows: 11 to 13 – high quality, 5 to 10 – moderate, and 1 to 4 – low quality.

### 5.6 Synthesis

Due to the heterogeneity of the included studies, a narrative synthesis was reported.

## EVIDENCE

### 6.1 Search results and study selection

After the removal of duplicates, the search identified 7,053 studies. Full texts (n=32) were reviewed, and twelve studies were included in this rapid review: economic evaluations (n = 7) and randomised controlled trials (n = 5).

### 6.2 Data extraction

Data extraction for the economic evaluations and randomised controlled trials are presented in Table 2 and Table 3, respectively.

**Table 3:**
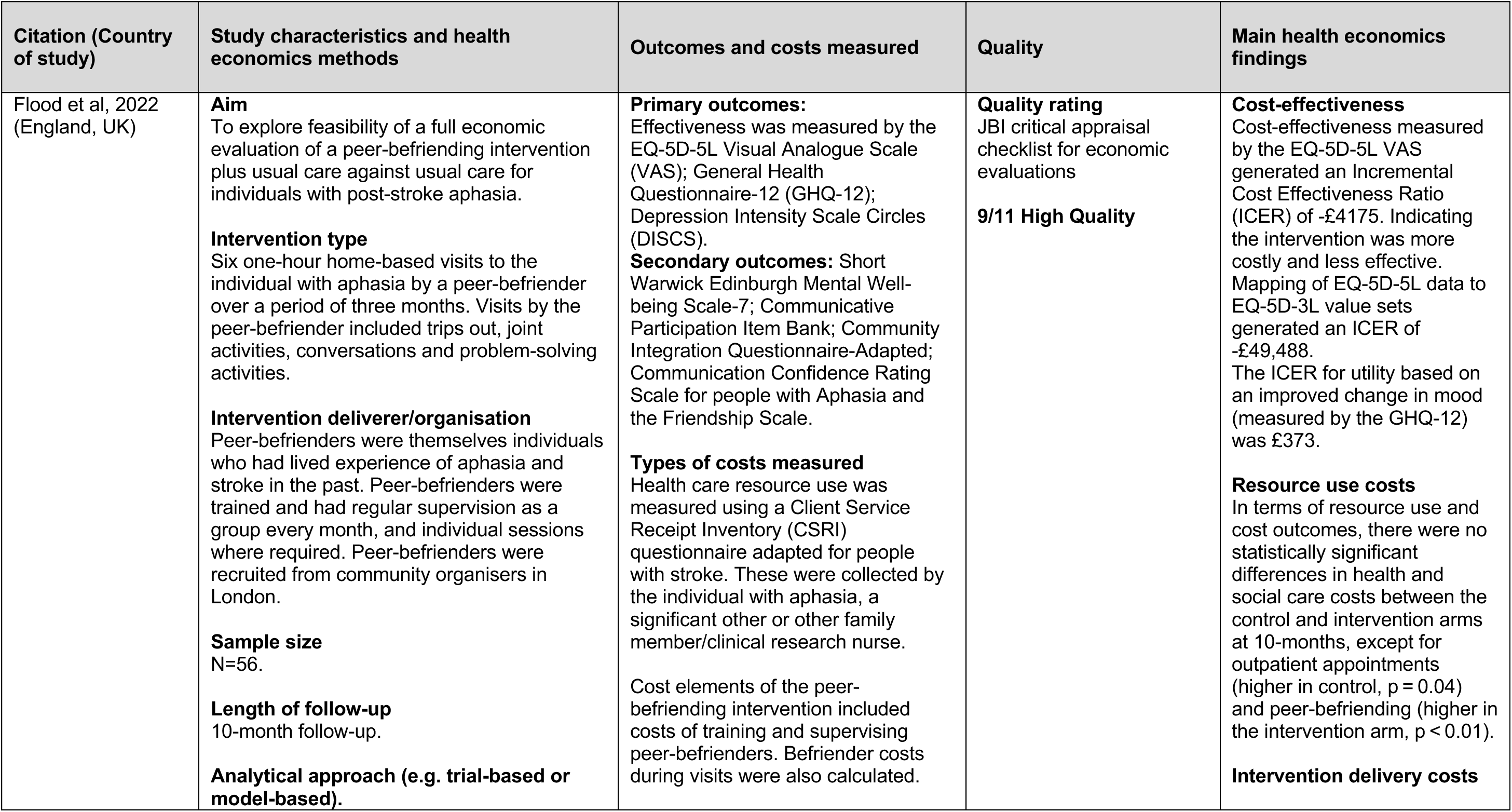
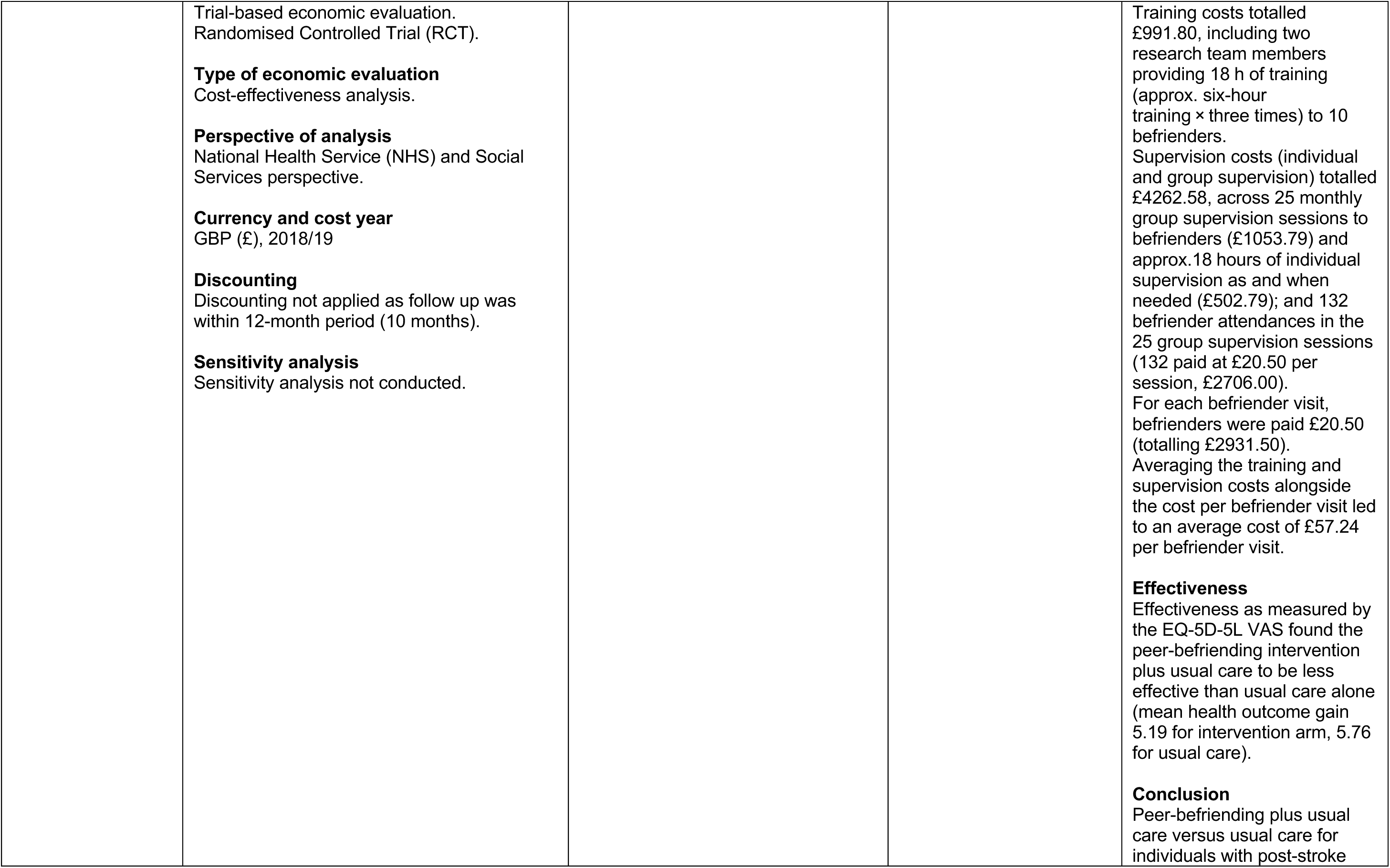
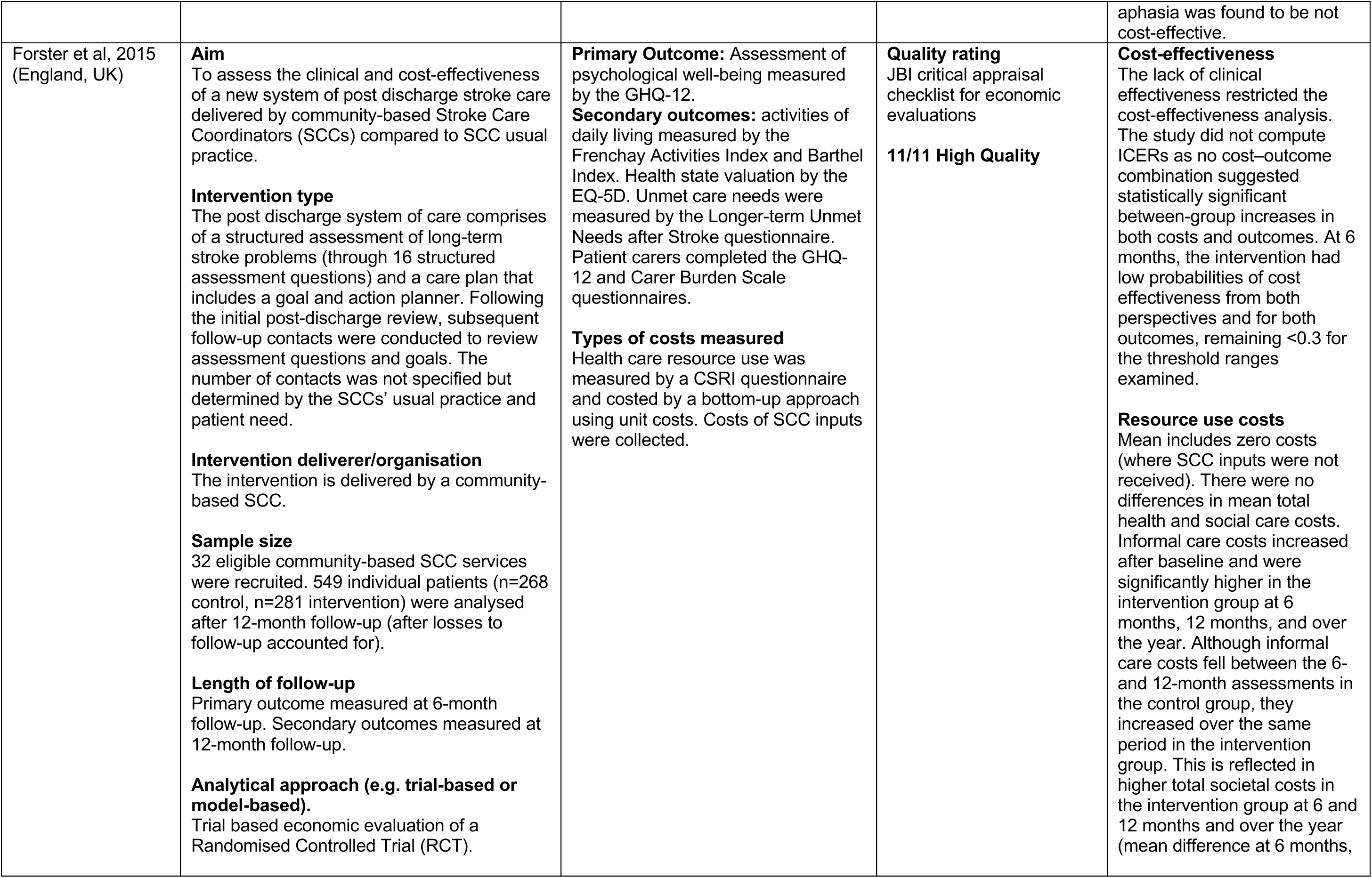
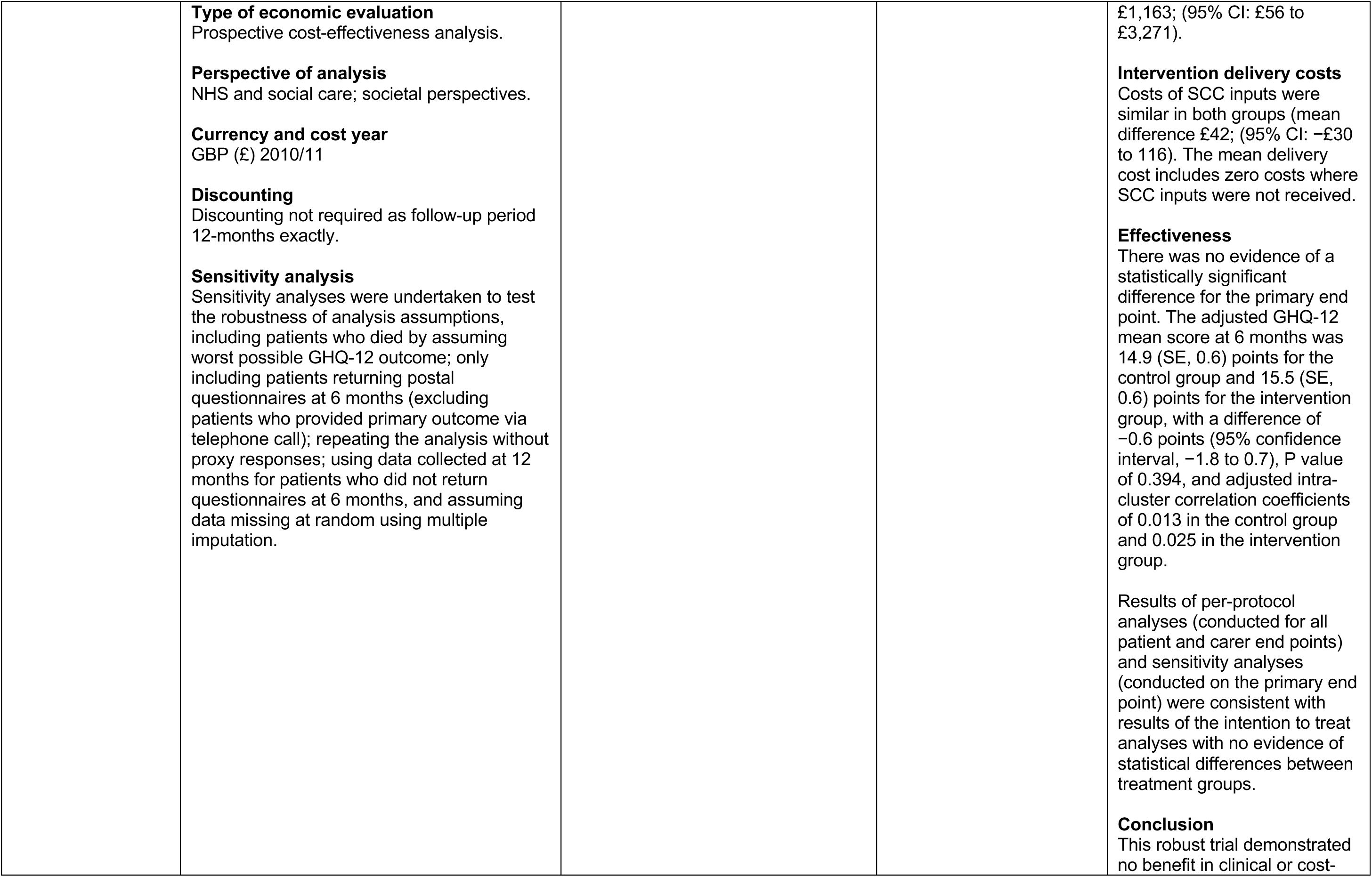
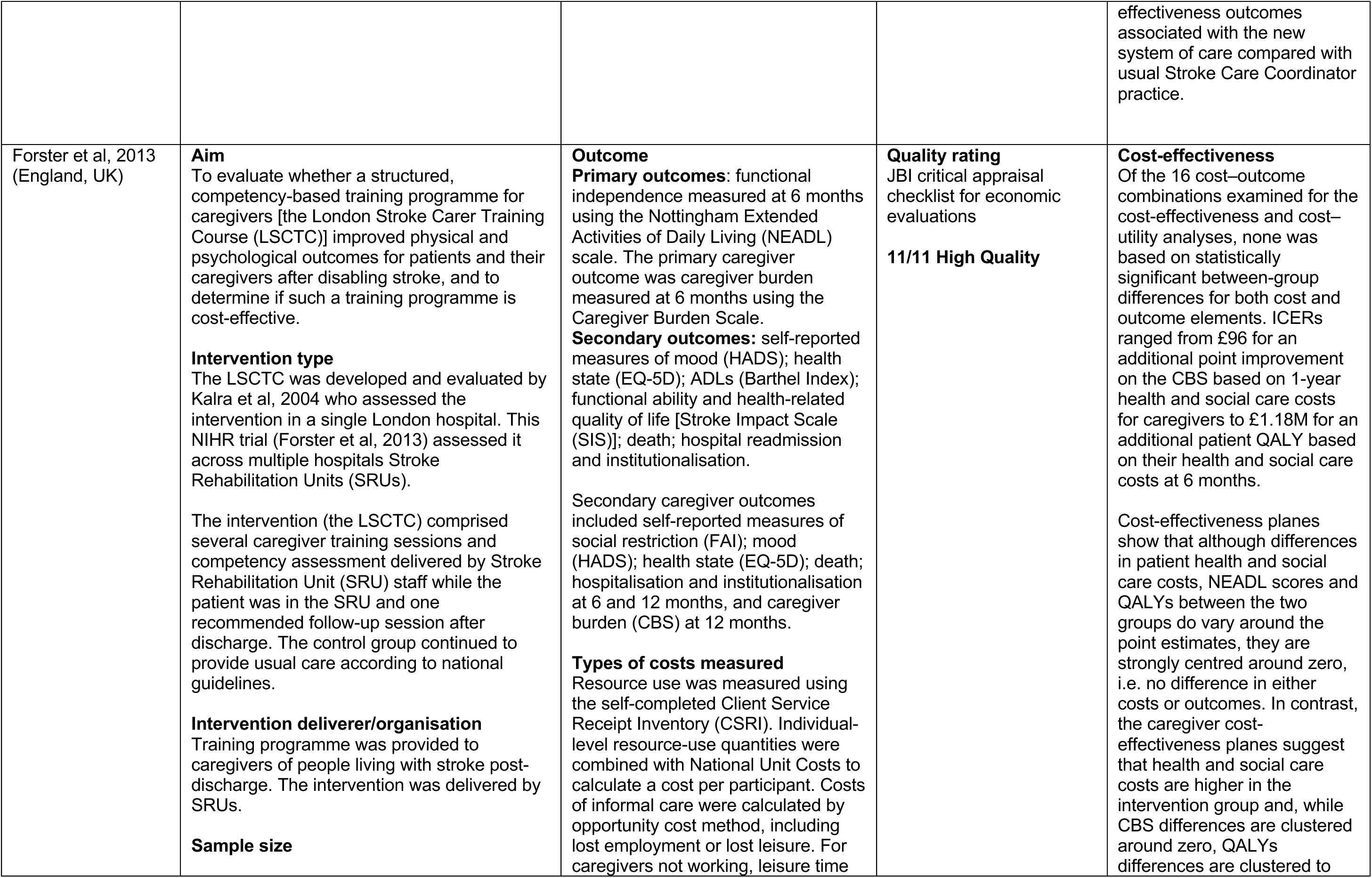
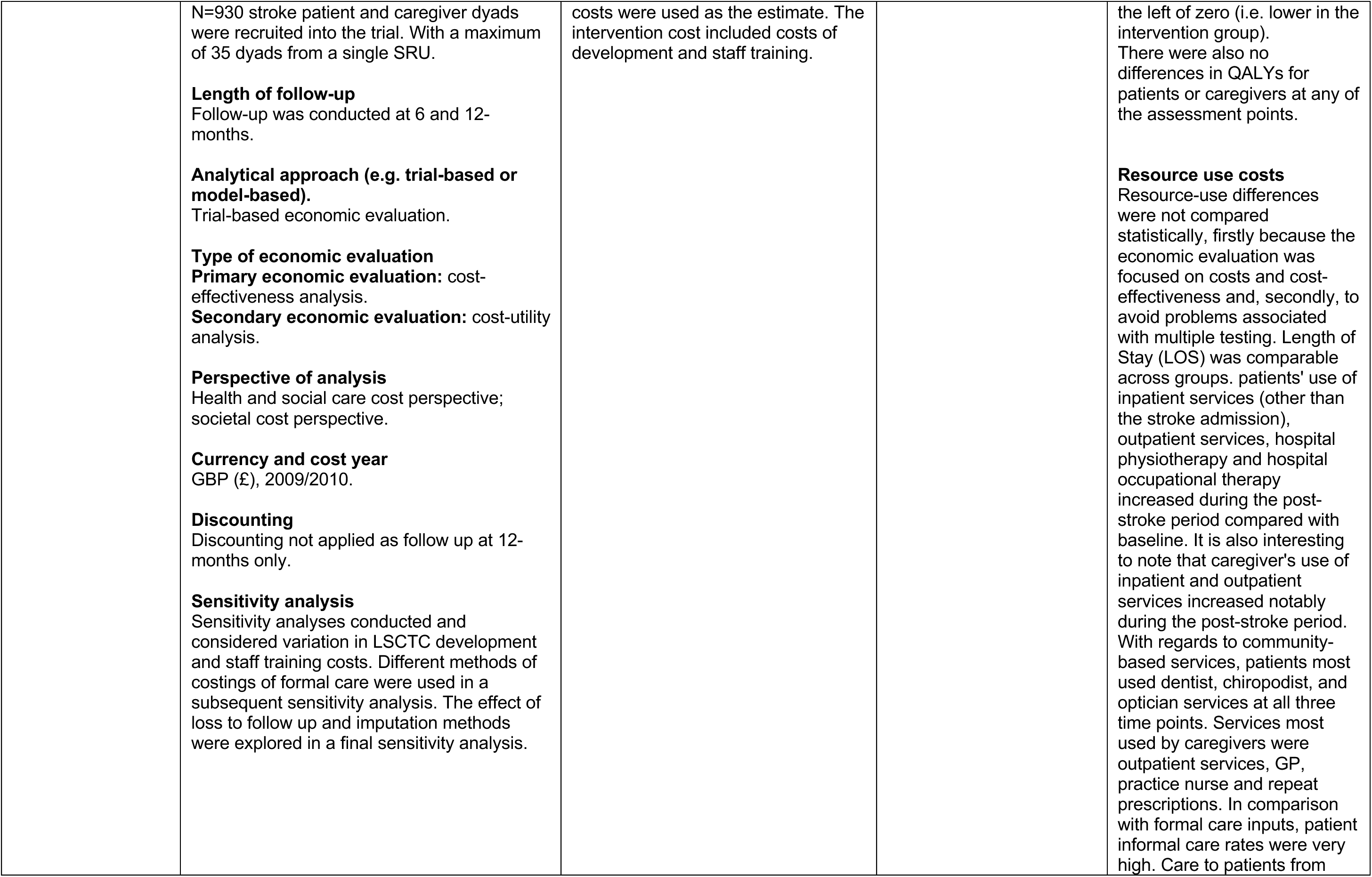
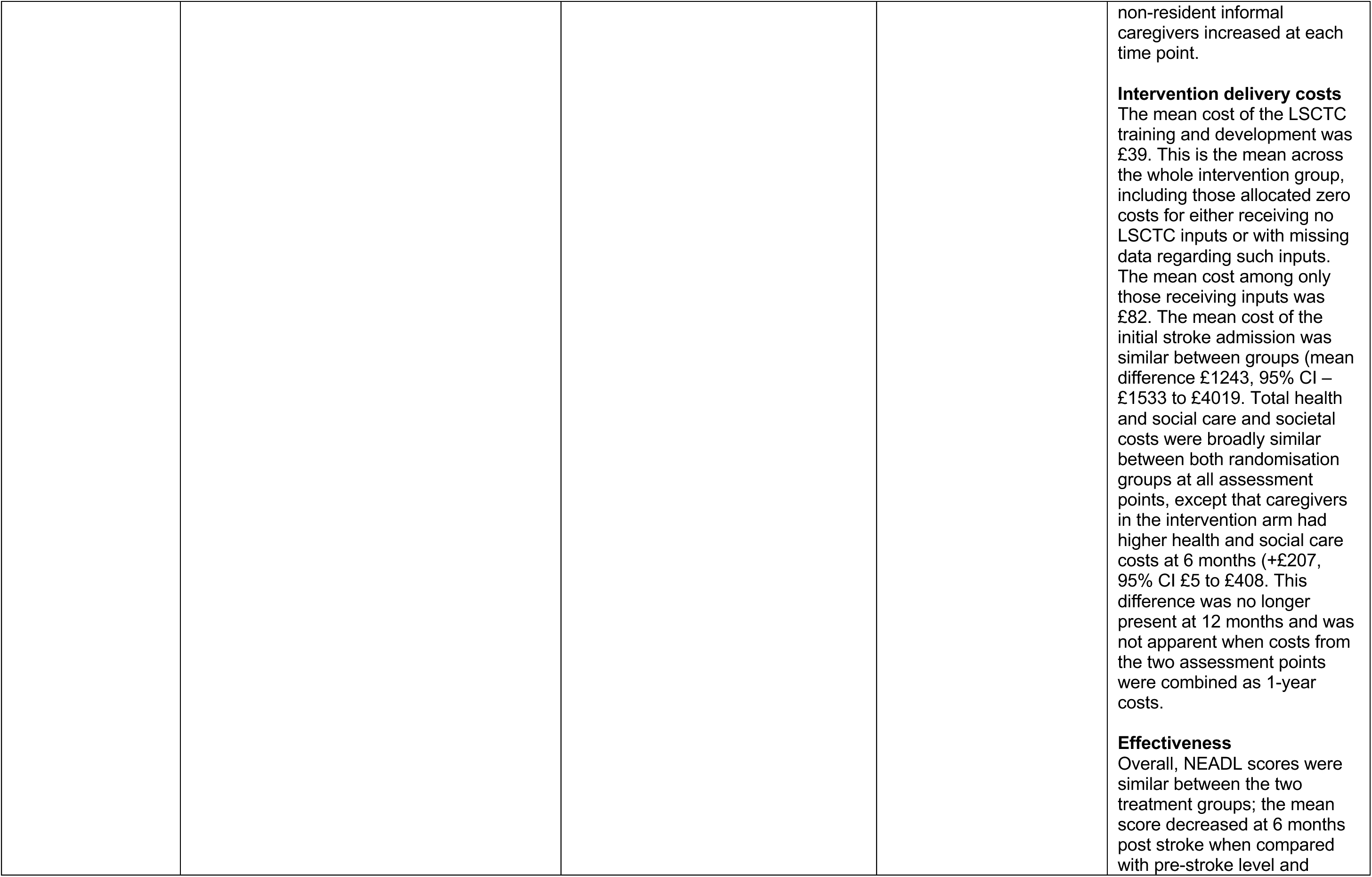
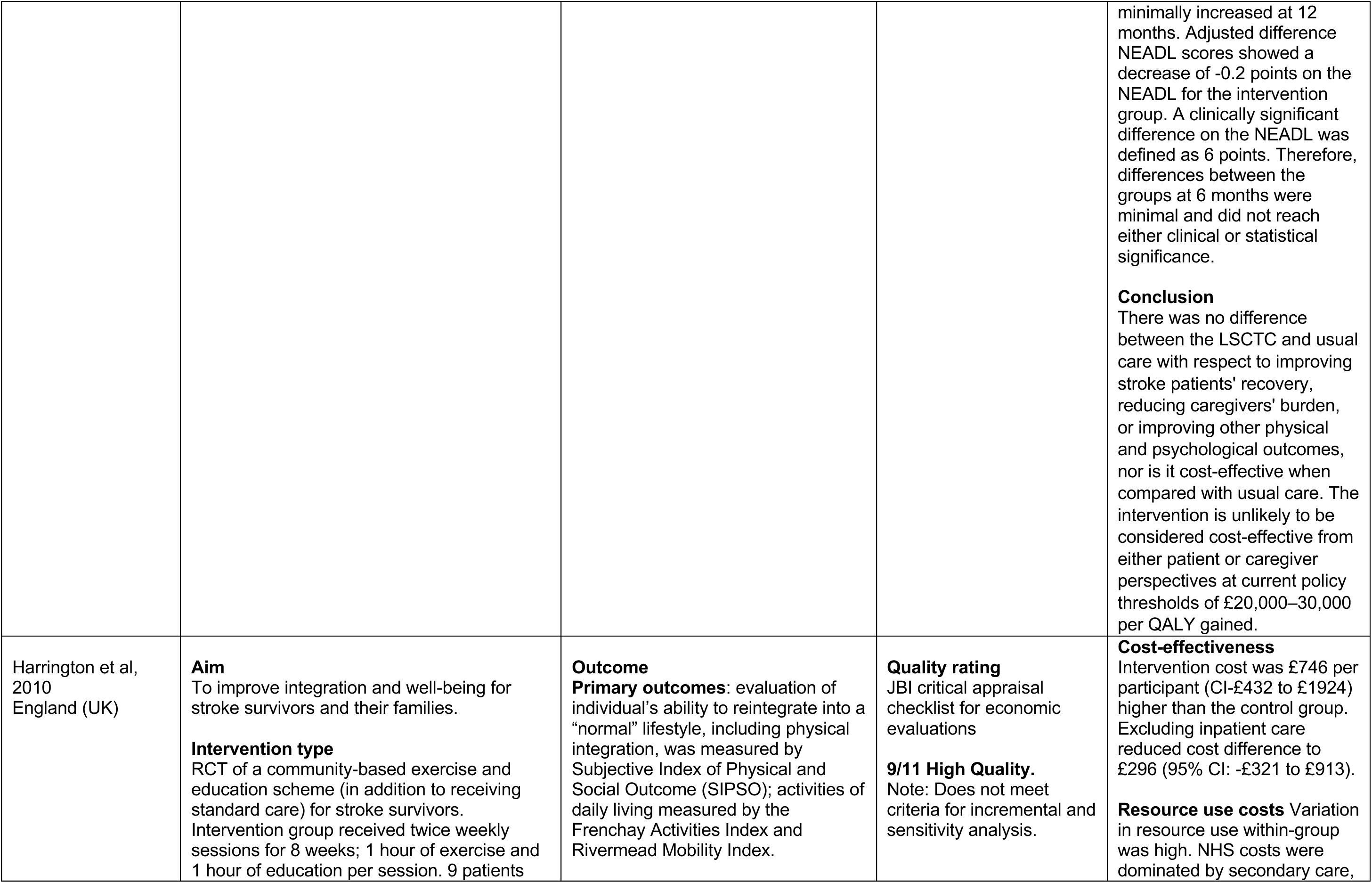
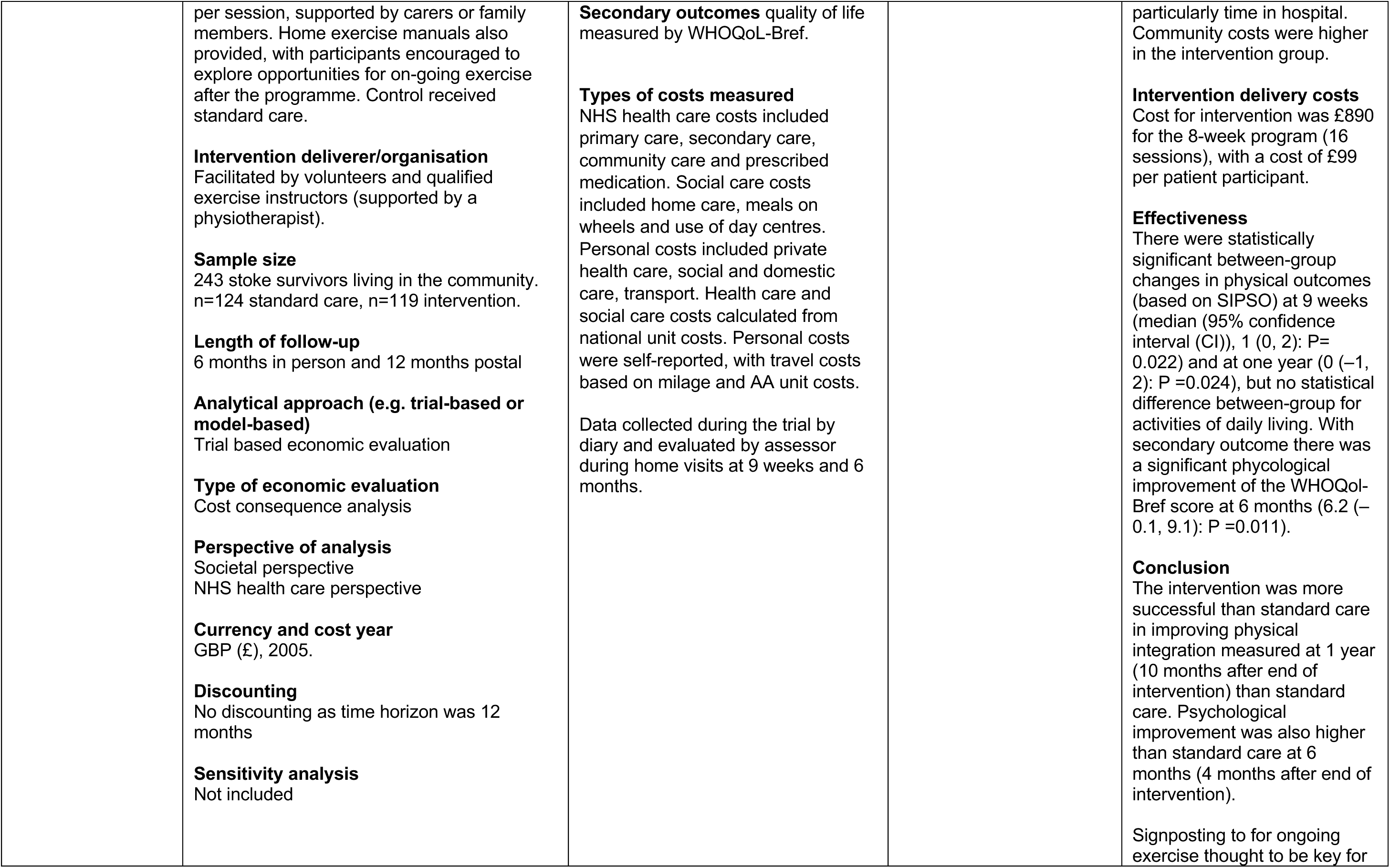
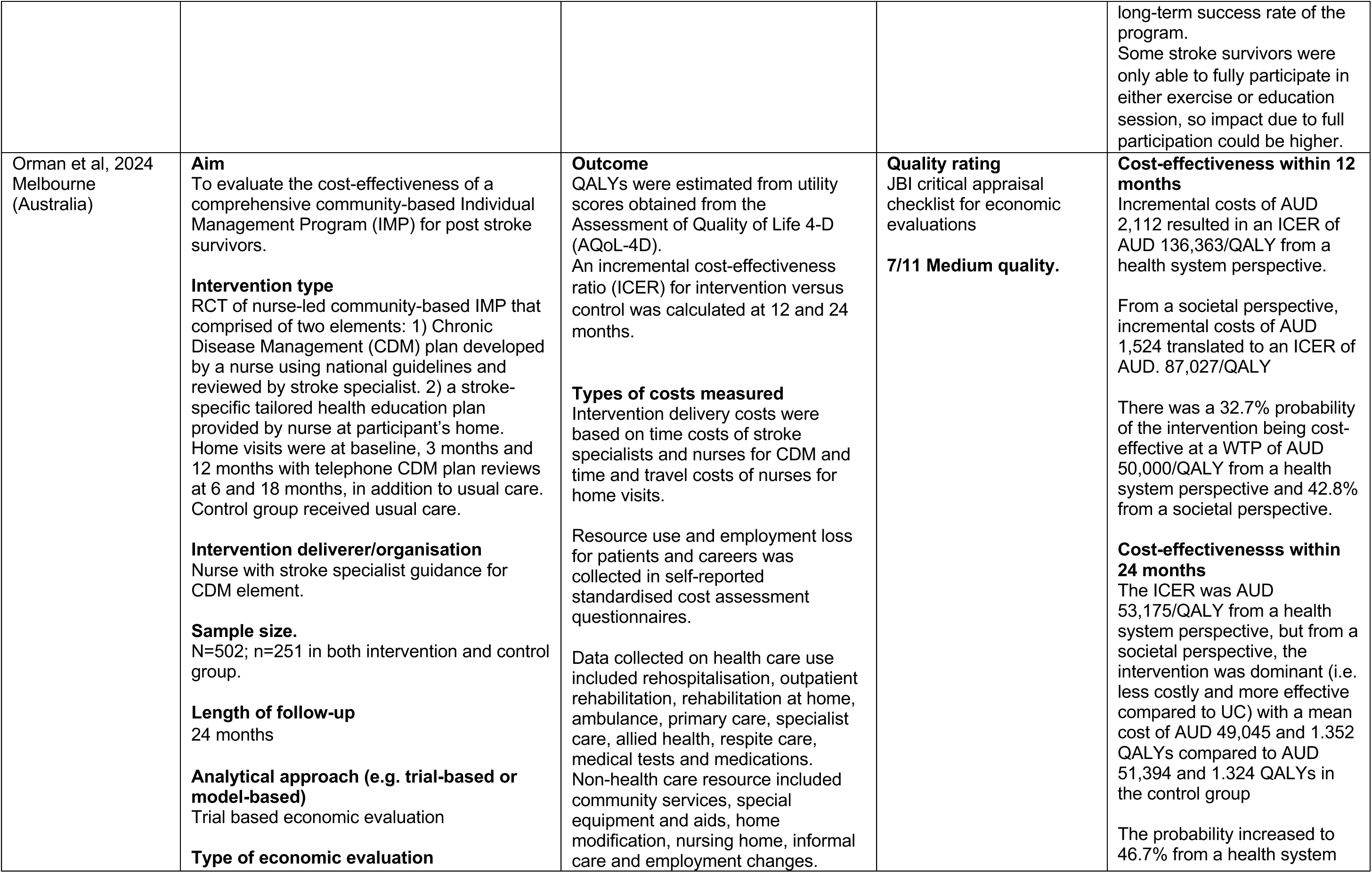
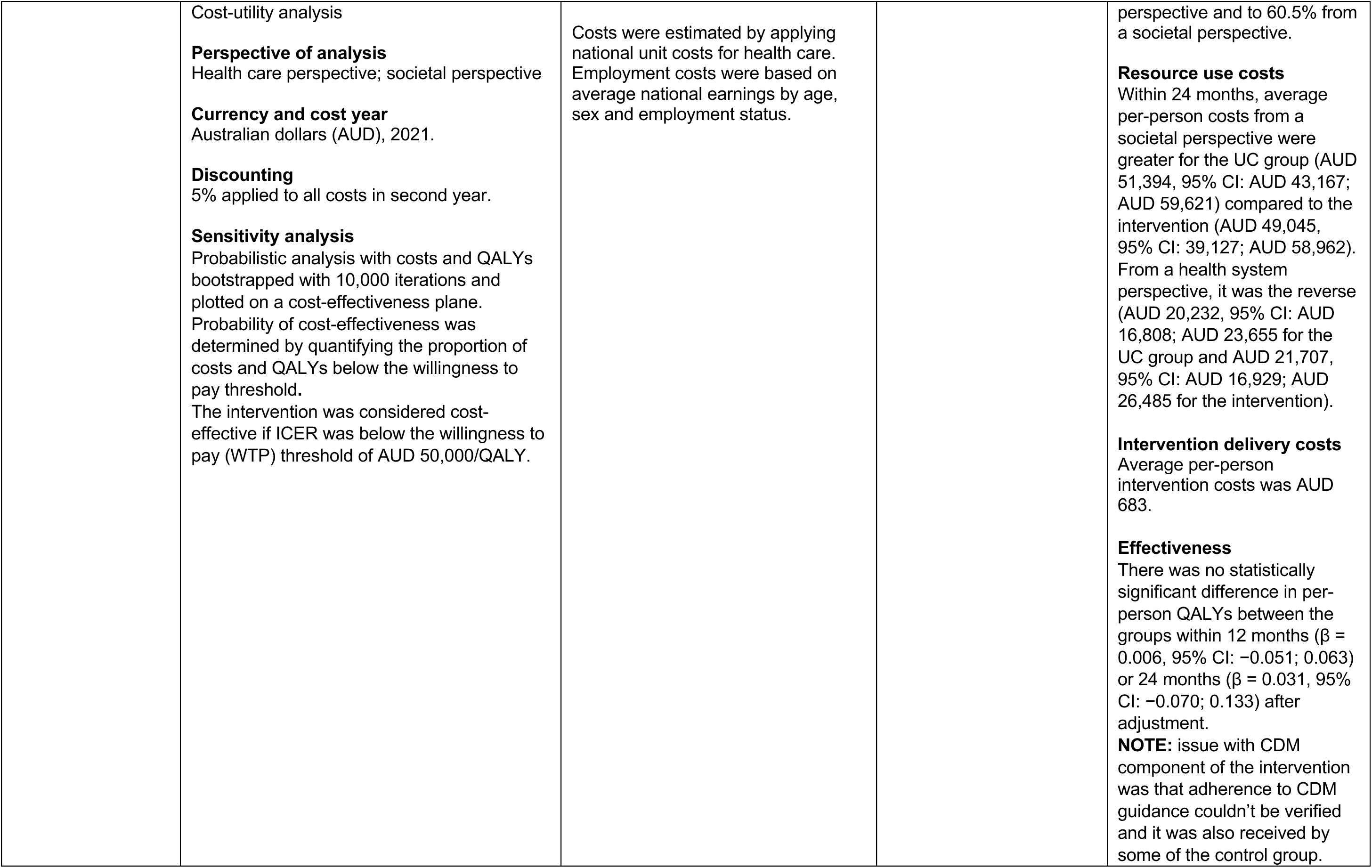
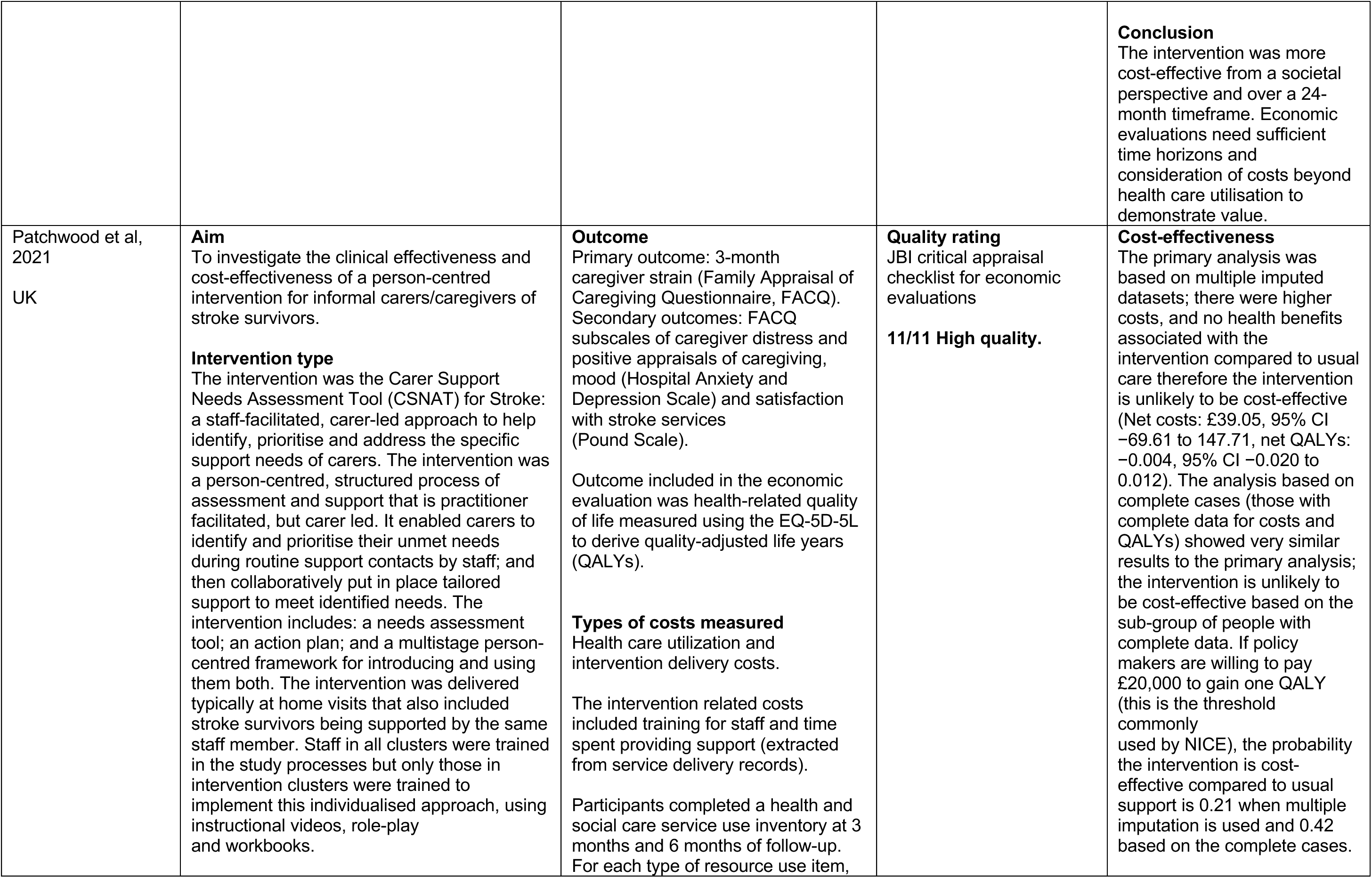
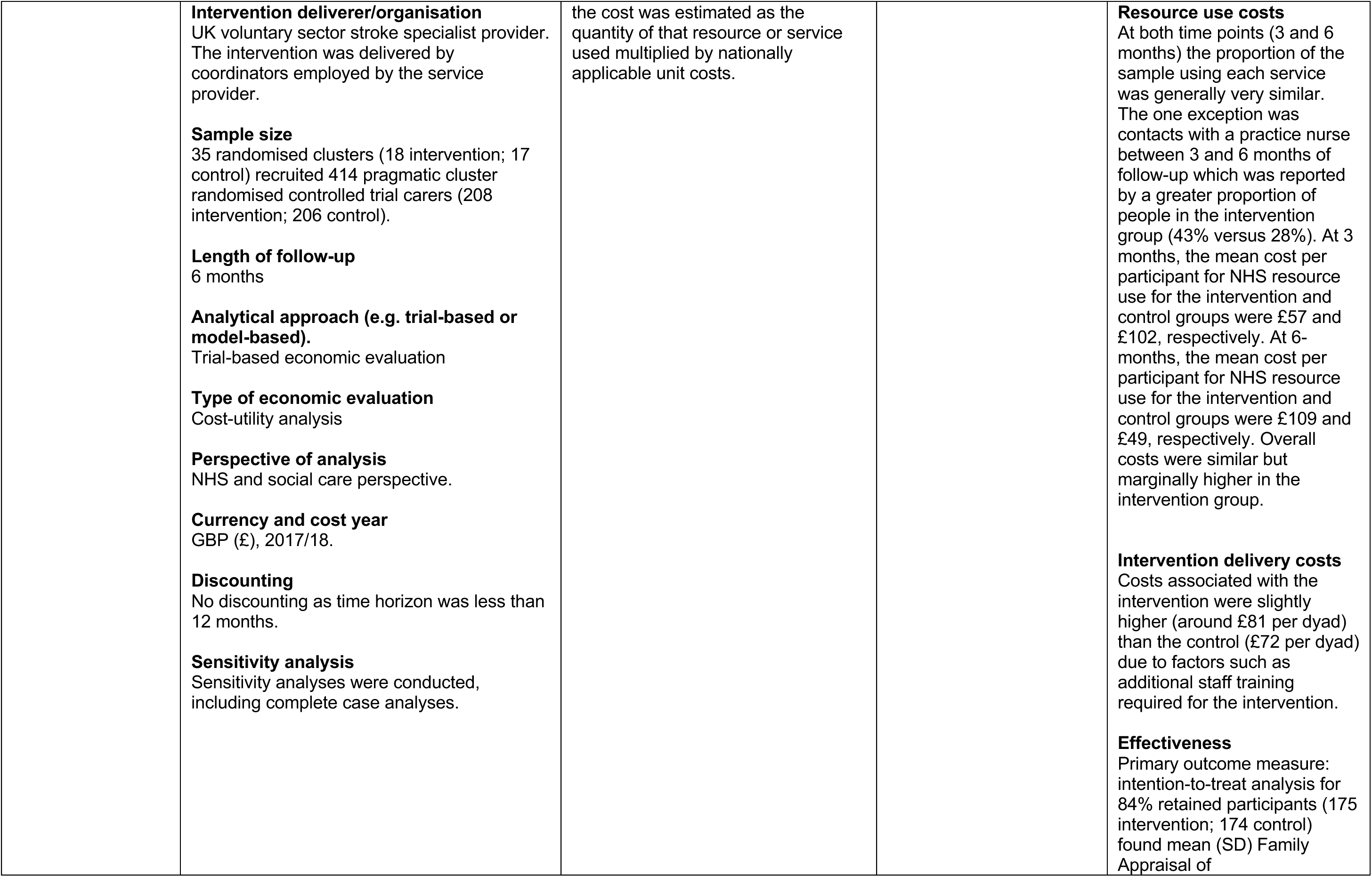
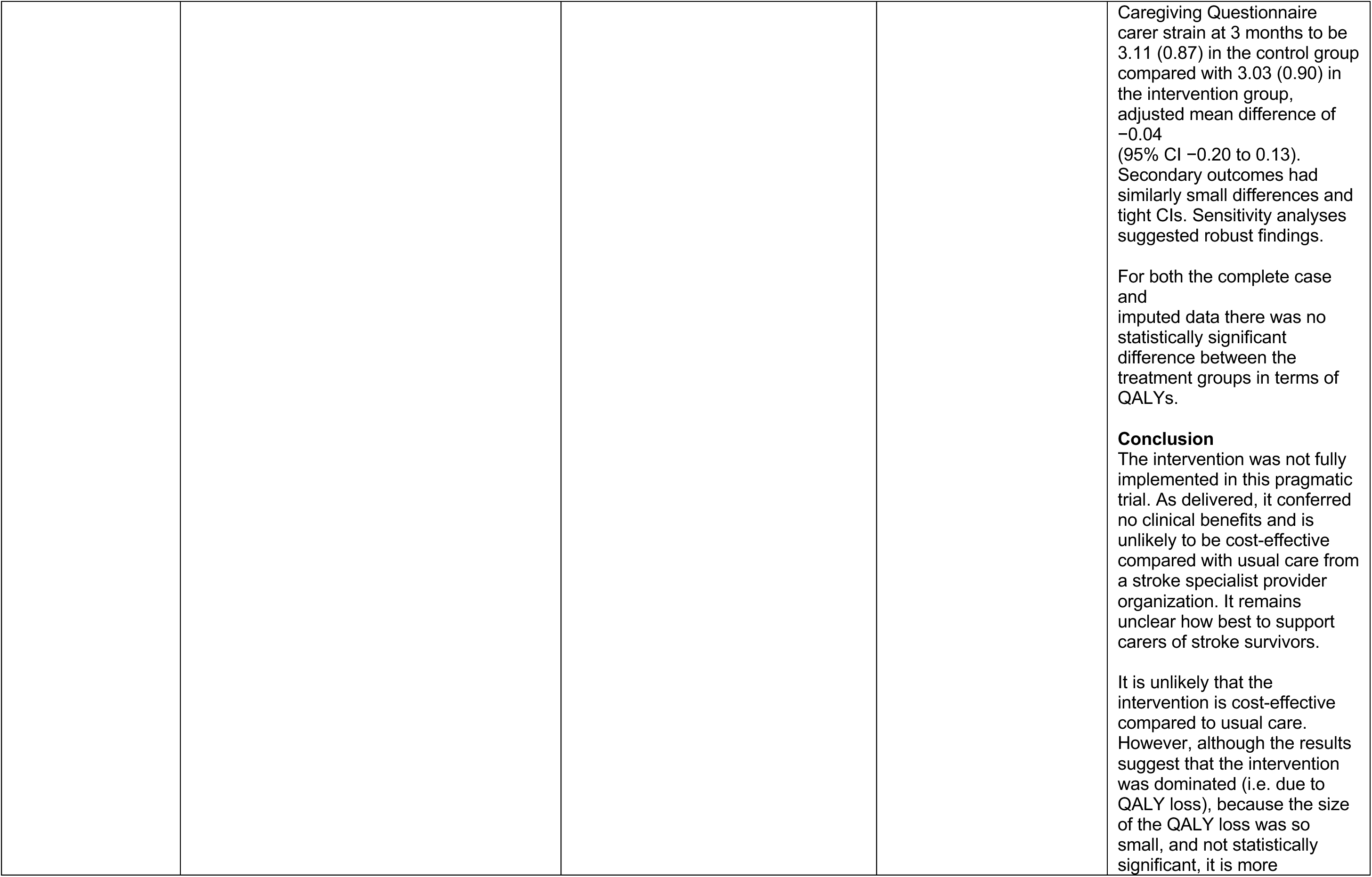
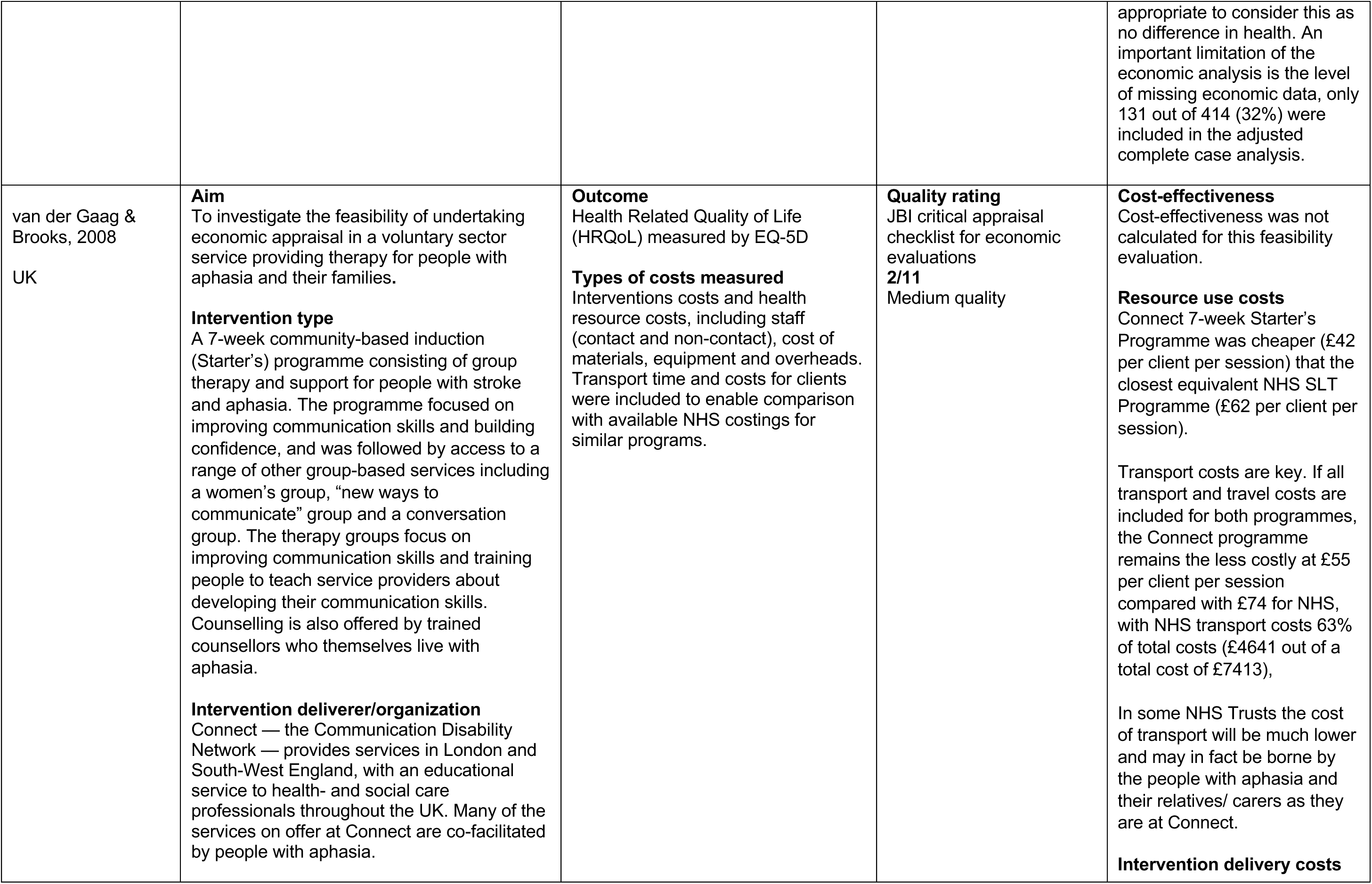
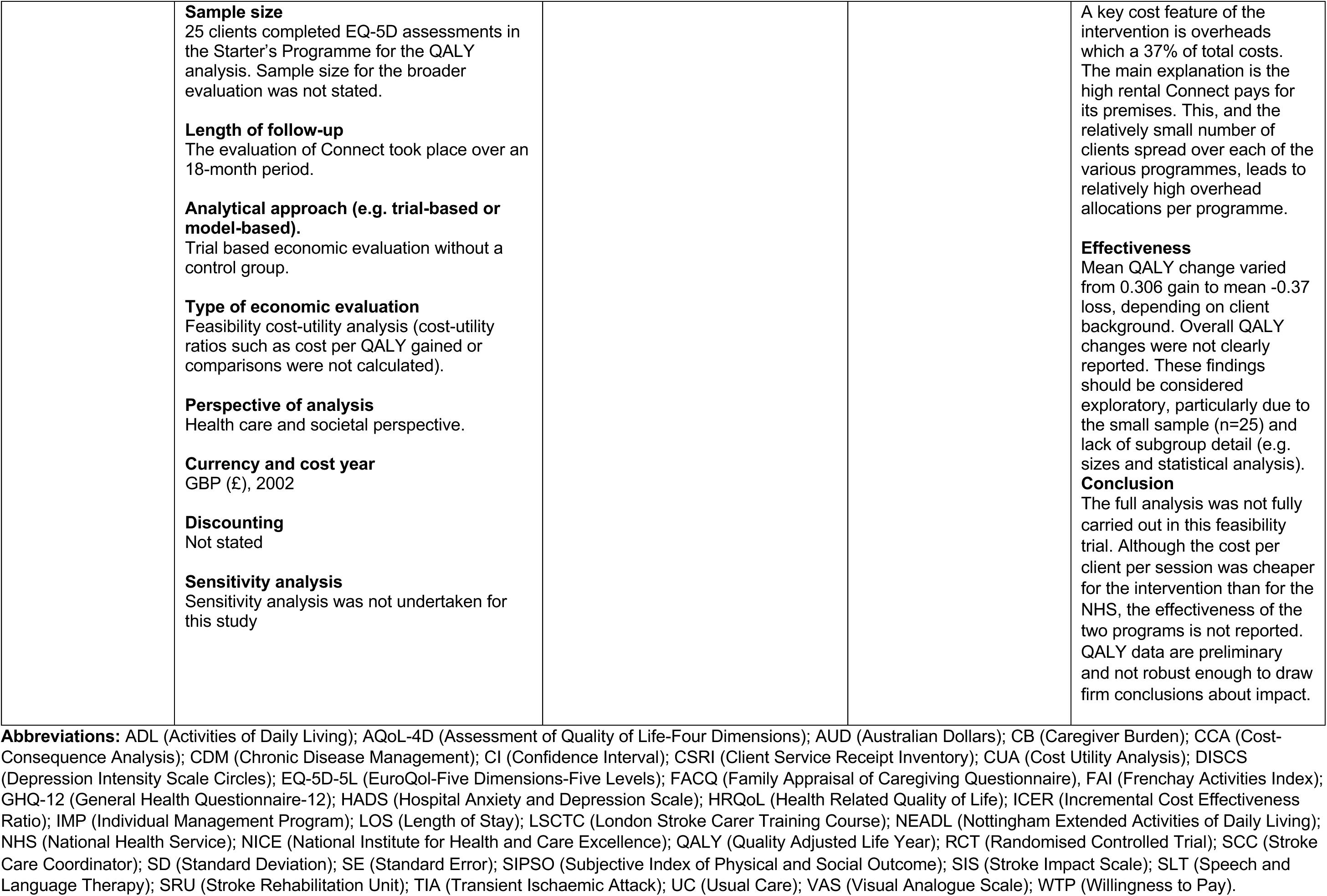
Evidence from included economic evaluations of life after stroke services (n = 7).

| Citation (Country of study) | Study characteristics and health economics methods | Outcomes and costs measured | Quality | Main health economics findings |
| --- | --- | --- | --- | --- |
| Flood et al, 2022 (England, UK) | **Aim**<br>To explore feasibility of a full economic evaluation of a peer-befriending intervention plus usual care against usual care for individuals with post-stroke aphasia.<br>**Intervention type**<br>Six one-hour home-based visits to the individual with aphasia by a peer-befriender over a period of three months. Visits by the peer-befriender included trips out, joint activities, conversations and problem-solving activities.<br>**Intervention deliverer/organisation**<br>Peer-befrienders were themselves individuals who had lived experience of aphasia and stroke in the past. Peer-befrienders were trained and had regular supervision as a group every month, and individual sessions where required. Peer-befrienders were recruited from community organisers in London.<br>**Sample size**<br>N=56.<br>**Length of follow-up**<br>10-month follow-up.<br>**Analytical approach (e.g. trial-based or model-based).** | **Primary outcomes:**<br>Effectiveness was measured by the EQ-5D-5L Visual Analogue Scale (VAS); General Health Questionnaire-12 (GHQ-12); Depression Intensity Scale Circles (DISCS).<br>**Secondary outcomes:** Short Warwick Edinburgh Mental Well-being Scale-7; Communicative Participation Item Bank; Community Integration Questionnaire-Adapted; Communication Confidence Rating Scale for people with Aphasia and the Friendship Scale.<br>**Types of costs measured**<br>Health care resource use was measured using a Client Service Receipt Inventory (CSRI) questionnaire adapted for people with stroke. These were collected by the individual with aphasia, a significant other or other family member/clinical research nurse.<br>Cost elements of the peer-befriending intervention included costs of training and supervising peer-befrienders. Befriender costs during visits were also calculated. | **Quality rating**<br>JBI critical appraisal checklist for economic evaluations<br>**9/11 High Quality** | **Cost-effectiveness**<br>Cost-effectiveness measured by the EQ-5D-5L VAS generated an Incremental Cost Effectiveness Ratio (ICER) of -£4175. Indicating the intervention was more costly and less effective. Mapping of EQ-5D-5L data to EQ-5D-3L value sets generated an ICER of -£49,488. The ICER for utility based on an improved change in mood (measured by the GHQ-12) was £373.<br>**Resource use costs**<br>In terms of resource use and cost outcomes, there were no statistically significant differences in health and social care costs between the control and intervention arms at 10-months, except for outpatient appointments (higher in control, $p = 0.04$) and peer-befriending (higher in the intervention arm, $p < 0.01$).<br>**Intervention delivery costs** |
|  | Trial-based economic evaluation.<br>Randomised Controlled Trial (RCT).<br>**Type of economic evaluation**<br>Cost-effectiveness analysis.<br>**Perspective of analysis**<br>National Health Service (NHS) and Social Services perspective.<br>**Currency and cost year**<br>GBP (£), 2018/19<br>**Discounting**<br>Discounting not applied as follow up was within 12-month period (10 months).<br>**Sensitivity analysis**<br>Sensitivity analysis not conducted. |  |  | Training costs totalled £991.80, including two research team members providing 18 h of training (approx. six-hour training × three times) to 10 befrienders.<br>Supervision costs (individual and group supervision) totalled £4262.58, across 25 monthly group supervision sessions to befrienders (£1053.79) and approx. 18 hours of individual supervision as and when needed (£502.79); and 132 befriender attendances in the 25 group supervision sessions (132 paid at £20.50 per session, £2706.00).<br>For each befriender visit, befrienders were paid £20.50 (totalling £2931.50).<br>Averaging the training and supervision costs alongside the cost per befriender visit led to an average cost of £57.24 per befriender visit.<br>**Effectiveness**<br>Effectiveness as measured by the EQ-5D-5L VAS found the peer-befriending intervention plus usual care to be less effective than usual care alone (mean health outcome gain 5.19 for intervention arm, 5.76 for usual care).<br>**Conclusion**<br>Peer-befriending plus usual care versus usual care for individuals with post-stroke<br>aphasia was found to be not cost-effective. |
| Forster et al, 2015<br>(England, UK) | **Aim**<br>To assess the clinical and cost-effectiveness of a new system of post discharge stroke care delivered by community-based Stroke Care Coordinators (SCCs) compared to SCC usual practice.<br>**Intervention type**<br>The post discharge system of care comprises of a structured assessment of long-term stroke problems (through 16 structured assessment questions) and a care plan that includes a goal and action planner. Following the initial post-discharge review, subsequent follow-up contacts were conducted to review assessment questions and goals. The number of contacts was not specified but determined by the SCCs' usual practice and patient need.<br>**Intervention deliverer/organisation**<br>The intervention is delivered by a community-based SCC.<br>**Sample size**<br>32 eligible community-based SCC services were recruited. 549 individual patients (n=268 control, n=281 intervention) were analysed after 12-month follow-up (after losses to follow-up accounted for).<br>**Length of follow-up**<br>Primary outcome measured at 6-month follow-up. Secondary outcomes measured at 12-month follow-up.<br>**Analytical approach (e.g. trial-based or model-based).**<br>Trial based economic evaluation of a Randomised Controlled Trial (RCT). | **Primary Outcome:** Assessment of psychological well-being measured by the GHQ-12.<br>**Secondary outcomes:** activities of daily living measured by the Frenchay Activities Index and Barthel Index. Health state valuation by the EQ-5D. Unmet care needs were measured by the Longer-term Unmet Needs after Stroke questionnaire. Patient carers completed the GHQ-12 and Carer Burden Scale questionnaires.<br>**Types of costs measured**<br>Health care resource use was measured by a CSRI questionnaire and costed by a bottom-up approach using unit costs. Costs of SCC inputs were collected. | **Quality rating**<br>JBI critical appraisal checklist for economic evaluations<br>**11/11 High Quality** | **Cost-effectiveness**<br>The lack of clinical effectiveness restricted the cost-effectiveness analysis. The study did not compute ICERs as no cost–outcome combination suggested statistically significant between-group increases in both costs and outcomes. At 6 months, the intervention had low probabilities of cost effectiveness from both perspectives and for both outcomes, remaining <0.3 for the threshold ranges examined.<br>**Resource use costs**<br>Mean includes zero costs (where SCC inputs were not received). There were no differences in mean total health and social care costs. Informal care costs increased after baseline and were significantly higher in the intervention group at 6 months, 12 months, and over the year. Although informal care costs fell between the 6- and 12-month assessments in the control group, they increased over the same period in the intervention group. This is reflected in higher total societal costs in the intervention group at 6 and 12 months and over the year (mean difference at 6 months, |
|  | **Type of economic evaluation**<br>Prospective cost-effectiveness analysis.<br>**Perspective of analysis**<br>NHS and social care; societal perspectives.<br>**Currency and cost year**<br>GBP (£) 2010/11<br>**Discounting**<br>Discounting not required as follow-up period 12-months exactly.<br>**Sensitivity analysis**<br>Sensitivity analyses were undertaken to test the robustness of analysis assumptions, including patients who died by assuming worst possible GHQ-12 outcome; only including patients returning postal questionnaires at 6 months (excluding patients who provided primary outcome via telephone call); repeating the analysis without proxy responses; using data collected at 12 months for patients who did not return questionnaires at 6 months, and assuming data missing at random using multiple imputation. |  |  | £1,163; (95% CI: £56 to £3,271).<br>**Intervention delivery costs**<br>Costs of SCC inputs were similar in both groups (mean difference £42; (95% CI: -£30 to 116). The mean delivery cost includes zero costs where SCC inputs were not received.<br>**Effectiveness**<br>There was no evidence of a statistically significant difference for the primary end point. The adjusted GHQ-12 mean score at 6 months was 14.9 (SE, 0.6) points for the control group and 15.5 (SE, 0.6) points for the intervention group, with a difference of -0.6 points (95% confidence interval, -1.8 to 0.7), P value of 0.394, and adjusted intra-cluster correlation coefficients of 0.013 in the control group and 0.025 in the intervention group.<br>Results of per-protocol analyses (conducted for all patient and carer end points) and sensitivity analyses (conducted on the primary end point) were consistent with results of the intention to treat analyses with no evidence of statistical differences between treatment groups.<br>**Conclusion**<br>This robust trial demonstrated no benefit in clinical or cost-effectiveness outcomes associated with the new system of care compared with usual Stroke Care Coordinator practice. |
| Forster et al, 2013<br>(England, UK) | **Aim**<br>To evaluate whether a structured, competency-based training programme for caregivers [the London Stroke Carer Training Course (LSCTC)] improved physical and psychological outcomes for patients and their caregivers after disabling stroke, and to determine if such a training programme is cost-effective.<br>**Intervention type**<br>The LSCTC was developed and evaluated by Kalra et al, 2004 who assessed the intervention in a single London hospital. This NIHR trial (Forster et al, 2013) assessed it across multiple hospitals Stroke Rehabilitation Units (SRUs).<br>The intervention (the LSCTC) comprised several caregiver training sessions and competency assessment delivered by Stroke Rehabilitation Unit (SRU) staff while the patient was in the SRU and one recommended follow-up session after discharge. The control group continued to provide usual care according to national guidelines.<br>**Intervention deliverer/organisation**<br>Training programme was provided to caregivers of people living with stroke post-discharge. The intervention was delivered by SRUs.<br>**Sample size** | **Outcome**<br>**Primary outcomes:** functional independence measured at 6 months using the Nottingham Extended Activities of Daily Living (NEADL) scale. The primary caregiver outcome was caregiver burden measured at 6 months using the Caregiver Burden Scale.<br>**Secondary outcomes:** self-reported measures of mood (HADS); health state (EQ-5D); ADLs (Barthel Index); functional ability and health-related quality of life [Stroke Impact Scale (SIS)]; death; hospital readmission and institutionalisation.<br>Secondary caregiver outcomes included self-reported measures of social restriction (FAI); mood (HADS); health state (EQ-5D); death; hospitalisation and institutionalisation at 6 and 12 months, and caregiver burden (CBS) at 12 months.<br>**Types of costs measured**<br>Resource use was measured using the self-completed Client Service Receipt Inventory (CSRI). Individual-level resource-use quantities were combined with National Unit Costs to calculate a cost per participant. Costs of informal care were calculated by opportunity cost method, including lost employment or lost leisure. For caregivers not working, leisure time | **Quality rating**<br>JBI critical appraisal checklist for economic evaluations<br>**11/11 High Quality** | **Cost-effectiveness**<br>Of the 16 cost–outcome combinations examined for the cost-effectiveness and cost–utility analyses, none was based on statistically significant between-group differences for both cost and outcome elements. ICERs ranged from £96 for an additional point improvement on the CBS based on 1-year health and social care costs for caregivers to £1.18M for an additional patient QALY based on their health and social care costs at 6 months.<br>Cost-effectiveness planes show that although differences in patient health and social care costs, NEADL scores and QALYs between the two groups do vary around the point estimates, they are strongly centred around zero, i.e. no difference in either costs or outcomes. In contrast, the caregiver cost-effectiveness planes suggest that health and social care costs are higher in the intervention group and, while CBS differences are clustered around zero, QALYs differences are clustered to |
|  | N=930 stroke patient and caregiver dyads were recruited into the trial. With a maximum of 35 dyads from a single SRU.<br>**Length of follow-up**<br>Follow-up was conducted at 6 and 12-months.<br>**Analytical approach (e.g. trial-based or model-based).**<br>Trial-based economic evaluation.<br>**Type of economic evaluation**<br>**Primary economic evaluation:** cost-effectiveness analysis.<br>**Secondary economic evaluation:** cost-utility analysis.<br>**Perspective of analysis**<br>Health and social care cost perspective; societal cost perspective.<br>**Currency and cost year**<br>GBP (£), 2009/2010.<br>**Discounting**<br>Discounting not applied as follow up at 12-months only.<br>**Sensitivity analysis**<br>Sensitivity analyses conducted and considered variation in LSCTC development and staff training costs. Different methods of costings of formal care were used in a subsequent sensitivity analysis. The effect of loss to follow up and imputation methods were explored in a final sensitivity analysis. | costs were used as the estimate. The intervention cost included costs of development and staff training. |  | the left of zero (i.e. lower in the intervention group). There were also no differences in QALYs for patients or caregivers at any of the assessment points.<br>**Resource use costs**<br>Resource-use differences were not compared statistically, firstly because the economic evaluation was focused on costs and cost-effectiveness and, secondly, to avoid problems associated with multiple testing. Length of Stay (LOS) was comparable across groups. patients' use of inpatient services (other than the stroke admission), outpatient services, hospital physiotherapy and hospital occupational therapy increased during the post-stroke period compared with baseline. It is also interesting to note that caregiver's use of inpatient and outpatient services increased notably during the post-stroke period. With regards to community-based services, patients most used dentist, chiropodist, and optician services at all three time points. Services most used by caregivers were outpatient services, GP, practice nurse and repeat prescriptions. In comparison with formal care inputs, patient informal care rates were very high. Care to patients from<br>non-resident informal caregivers increased at each time point.<br>**Intervention delivery costs**<br>The mean cost of the LSCTC training and development was £39. This is the mean across the whole intervention group, including those allocated zero costs for either receiving no LSCTC inputs or with missing data regarding such inputs. The mean cost among only those receiving inputs was £82. The mean cost of the initial stroke admission was similar between groups (mean difference £1243, 95% CI – £1533 to £4019. Total health and social care and societal costs were broadly similar between both randomisation groups at all assessment points, except that caregivers in the intervention arm had higher health and social care costs at 6 months (+£207, 95% CI £5 to £408. This difference was no longer present at 12 months and was not apparent when costs from the two assessment points were combined as 1-year costs.<br>**Effectiveness**<br>Overall, NEADL scores were similar between the two treatment groups; the mean score decreased at 6 months post stroke when compared with pre-stroke level and<br>minimally increased at 12 months. Adjusted difference NEADL scores showed a decrease of -0.2 points on the NEADL for the intervention group. A clinically significant difference on the NEADL was defined as 6 points. Therefore, differences between the groups at 6 months were minimal and did not reach either clinical or statistical significance.<br>**Conclusion**<br>There was no difference between the LSCTC and usual care with respect to improving stroke patients' recovery, reducing caregivers' burden, or improving other physical and psychological outcomes, nor is it cost-effective when compared with usual care. The intervention is unlikely to be considered cost-effective from either patient or caregiver perspectives at current policy thresholds of £20,000–30,000 per QALY gained. |
| Harrington et al, 2010<br>England (UK) | **Aim**<br>To improve integration and well-being for stroke survivors and their families.<br>**Intervention type**<br>RCT of a community-based exercise and education scheme (in addition to receiving standard care) for stroke survivors. Intervention group received twice weekly sessions for 8 weeks; 1 hour of exercise and 1 hour of education per session. 9 patients | **Outcome**<br>**Primary outcomes:** evaluation of individual's ability to reintegrate into a "normal" lifestyle, including physical integration, was measured by Subjective Index of Physical and Social Outcome (SIPSO); activities of daily living measured by the Frenchay Activities Index and Rivermead Mobility Index. | **Quality rating**<br>JBI critical appraisal checklist for economic evaluations<br>**9/11 High Quality.**<br>Note: Does not meet criteria for incremental and sensitivity analysis. | **Cost-effectiveness**<br>Intervention cost was £746 per participant (CI-£432 to £1924) higher than the control group. Excluding inpatient care reduced cost difference to £296 (95% CI: -£321 to £913).<br>**Resource use costs** Variation in resource use within-group was high. NHS costs were dominated by secondary care, |

|  |  |  |  |
| --- | --- | --- | --- |
|  | per session, supported by carers or family members. Home exercise manuals also provided, with participants encouraged to explore opportunities for on-going exercise after the programme. Control received standard care.<br>**Intervention deliverer/organisation**<br>Facilitated by volunteers and qualified exercise instructors (supported by a physiotherapist).<br>**Sample size**<br>243 stroke survivors living in the community. n=124 standard care, n=119 intervention.<br>**Length of follow-up**<br>6 months in person and 12 months postal<br>**Analytical approach (e.g. trial-based or model-based)**<br>Trial based economic evaluation<br>**Type of economic evaluation**<br>Cost consequence analysis<br>**Perspective of analysis**<br>Societal perspective<br>NHS health care perspective<br>**Currency and cost year**<br>GBP (£), 2005.<br>**Discounting**<br>No discounting as time horizon was 12 months<br>**Sensitivity analysis**<br>Not included | **Secondary outcomes** quality of life measured by WHOQoL-Bref.<br>**Types of costs measured**<br>NHS health care costs included primary care, secondary care, community care and prescribed medication. Social care costs included home care, meals on wheels and use of day centres. Personal costs included private health care, social and domestic care, transport. Health care and social care costs calculated from national unit costs. Personal costs were self-reported, with travel costs based on mileage and AA unit costs.<br>Data collected during the trial by diary and evaluated by assessor during home visits at 9 weeks and 6 months. | particularly time in hospital. Community costs were higher in the intervention group.<br>**Intervention delivery costs**<br>Cost for intervention was £890 for the 8-week program (16 sessions), with a cost of £99 per patient participant.<br>**Effectiveness**<br>There were statistically significant between-group changes in physical outcomes (based on SIPSO) at 9 weeks (median (95% confidence interval (CI)), 1 (0, 2); P= 0.022) and at one year (0 (-1, 2); P =0.024), but no statistical difference between-group for activities of daily living. With secondary outcome there was a significant psychological improvement of the WHOQoL-Bref score at 6 months (6.2 (-0.1, 9.1); P =0.011).<br>**Conclusion**<br>The intervention was more successful than standard care in improving physical integration measured at 1 year (10 months after end of intervention) than standard care. Psychological improvement was also higher than standard care at 6 months (4 months after end of intervention).<br>Signposting to for ongoing exercise thought to be key for |
|  |  |  |  | long-term success rate of the program.<br>Some stroke survivors were only able to fully participate in either exercise or education session, so impact due to full participation could be higher. |
| Orman et al, 2024<br>Melbourne<br>(Australia) | **Aim**<br>To evaluate the cost-effectiveness of a comprehensive community-based Individual Management Program (IMP) for post stroke survivors.<br>**Intervention type**<br>RCT of nurse-led community-based IMP that comprised of two elements: 1) Chronic Disease Management (CDM) plan developed by a nurse using national guidelines and reviewed by stroke specialist. 2) a stroke-specific tailored health education plan provided by nurse at participant's home. Home visits were at baseline, 3 months and 12 months with telephone CDM plan reviews at 6 and 18 months, in addition to usual care. Control group received usual care.<br>**Intervention deliverer/organisation**<br>Nurse with stroke specialist guidance for CDM element.<br>**Sample size.**<br>N=502; n=251 in both intervention and control group.<br>**Length of follow-up**<br>24 months<br>**Analytical approach (e.g. trial-based or model-based)**<br>Trial based economic evaluation<br>**Type of economic evaluation** | **Outcome**<br>QALYs were estimated from utility scores obtained from the Assessment of Quality of Life 4-D (AQoL-4D).<br>An incremental cost-effectiveness ratio (ICER) for intervention versus control was calculated at 12 and 24 months.<br>**Types of costs measured**<br>Intervention delivery costs were based on time costs of stroke specialists and nurses for CDM and time and travel costs of nurses for home visits.<br>Resource use and employment loss for patients and careers was collected in self-reported standardised cost assessment questionnaires.<br>Data collected on health care use included rehospitalisation, outpatient rehabilitation, rehabilitation at home, ambulance, primary care, specialist care, allied health, respite care, medical tests and medications. Non-health care resource included community services, special equipment and aids, home modification, nursing home, informal care and employment changes. | **Quality rating**<br>JBI critical appraisal checklist for economic evaluations<br>**7/11 Medium quality.** | **Cost-effectiveness within 12 months**<br>Incremental costs of AUD 2,112 resulted in an ICER of AUD 136,363/QALY from a health system perspective.<br>From a societal perspective, incremental costs of AUD 1,524 translated to an ICER of AUD. 87,027/QALY<br>There was a 32.7% probability of the intervention being cost-effective at a WTP of AUD 50,000/QALY from a health system perspective and 42.8% from a societal perspective.<br>**Cost-effectiveness within 24 months**<br>The ICER was AUD 53,175/QALY from a health system perspective, but from a societal perspective, the intervention was dominant (i.e. less costly and more effective compared to UC) with a mean cost of AUD 49,045 and 1.352 QALYs compared to AUD 51,394 and 1.324 QALYs in the control group<br>The probability increased to 46.7% from a health system |

|  |  |  |  |
| --- | --- | --- | --- |
|  | Cost-utility analysis<br>**Perspective of analysis**<br>Health care perspective; societal perspective<br>**Currency and cost year**<br>Australian dollars (AUD), 2021.<br>**Discounting**<br>5% applied to all costs in second year.<br>**Sensitivity analysis**<br>Probabilistic analysis with costs and QALYs bootstrapped with 10,000 iterations and plotted on a cost-effectiveness plane. Probability of cost-effectiveness was determined by quantifying the proportion of costs and QALYs below the willingness to pay threshold. The intervention was considered cost-effective if ICER was below the willingness to pay (WTP) threshold of AUD 50,000/QALY. | Costs were estimated by applying national unit costs for health care. Employment costs were based on average national earnings by age, sex and employment status. | perspective and to 60.5% from a societal perspective.<br>**Resource use costs**<br>Within 24 months, average per-person costs from a societal perspective were greater for the UC group (AUD 51,394, 95% CI: AUD 43,167; AUD 59,621) compared to the intervention (AUD 49,045, 95% CI: 39,127; AUD 58,962). From a health system perspective, it was the reverse (AUD 20,232, 95% CI: AUD 16,808; AUD 23,655 for the UC group and AUD 21,707, 95% CI: AUD 16,929; AUD 26,485 for the intervention).<br>**Intervention delivery costs**<br>Average per-person intervention costs was AUD 683.<br>**Effectiveness**<br>There was no statistically significant difference in per-person QALYs between the groups within 12 months ($\beta = 0.006$, 95% CI: $-0.051$; $0.063$) or 24 months ($\beta = 0.031$, 95% CI: $-0.070$; $0.133$) after adjustment.<br>**NOTE:** issue with CDM component of the intervention was that adherence to CDM guidance couldn't be verified and it was also received by some of the control group. |
|  |  |  |  | **Conclusion**<br>The intervention was more cost-effective from a societal perspective and over a 24-month timeframe. Economic evaluations need sufficient time horizons and consideration of costs beyond health care utilisation to demonstrate value. |
| Patchwood et al, 2021<br>UK | **Aim**<br>To investigate the clinical effectiveness and cost-effectiveness of a person-centred intervention for informal carers/caregivers of stroke survivors.<br>**Intervention type**<br>The intervention was the Carer Support Needs Assessment Tool (CSNAT) for Stroke: a staff-facilitated, carer-led approach to help identify, prioritise and address the specific support needs of carers. The intervention was a person-centred, structured process of assessment and support that is practitioner facilitated, but carer led. It enabled carers to identify and prioritise their unmet needs during routine support contacts by staff; and then collaboratively put in place tailored support to meet identified needs. The intervention includes: a needs assessment tool; an action plan; and a multistage person-centred framework for introducing and using them both. The intervention was delivered typically at home visits that also included stroke survivors being supported by the same staff member. Staff in all clusters were trained in the study processes but only those in intervention clusters were trained to implement this individualised approach, using instructional videos, role-play and workbooks. | **Outcome**<br>Primary outcome: 3-month caregiver strain (Family Appraisal of Caregiving Questionnaire, FACQ). Secondary outcomes: FACQ subscales of caregiver distress and positive appraisals of caregiving, mood (Hospital Anxiety and Depression Scale) and satisfaction with stroke services (Pound Scale).<br>Outcome included in the economic evaluation was health-related quality of life measured using the EQ-5D-5L to derive quality-adjusted life years (QALYs).<br>**Types of costs measured**<br>Health care utilization and intervention delivery costs.<br>The intervention related costs included training for staff and time spent providing support (extracted from service delivery records).<br>Participants completed a health and social care service use inventory at 3 months and 6 months of follow-up. For each type of resource use item, | **Quality rating**<br>JBI critical appraisal checklist for economic evaluations<br>**11/11 High quality.** | **Cost-effectiveness**<br>The primary analysis was based on multiple imputed datasets; there were higher costs, and no health benefits associated with the intervention compared to usual care therefore the intervention is unlikely to be cost-effective (Net costs: £39.05, 95% CI -69.61 to 147.71, net QALYs: -0.004, 95% CI -0.020 to 0.012). The analysis based on complete cases (those with complete data for costs and QALYs) showed very similar results to the primary analysis; the intervention is unlikely to be cost-effective based on the sub-group of people with complete data. If policy makers are willing to pay £20,000 to gain one QALY (this is the threshold commonly used by NICE), the probability the intervention is cost-effective compared to usual support is 0.21 when multiple imputation is used and 0.42 based on the complete cases. |

|  |  |  |  |
| --- | --- | --- | --- |
|  | **Intervention deliverer/organisation**<br>UK voluntary sector stroke specialist provider. The intervention was delivered by coordinators employed by the service provider.<br>**Sample size**<br>35 randomised clusters (18 intervention; 17 control) recruited 414 pragmatic cluster randomised controlled trial carers (208 intervention; 206 control).<br>**Length of follow-up**<br>6 months<br>**Analytical approach (e.g. trial-based or model-based).**<br>Trial-based economic evaluation<br>**Type of economic evaluation**<br>Cost-utility analysis<br>**Perspective of analysis**<br>NHS and social care perspective.<br>**Currency and cost year**<br>GBP (£), 2017/18.<br>**Discounting**<br>No discounting as time horizon was less than 12 months.<br>**Sensitivity analysis**<br>Sensitivity analyses were conducted, including complete case analyses. | the cost was estimated as the quantity of that resource or service used multiplied by nationally applicable unit costs. | **Resource use costs**<br>At both time points (3 and 6 months) the proportion of the sample using each service was generally very similar. The one exception was contacts with a practice nurse between 3 and 6 months of follow-up which was reported by a greater proportion of people in the intervention group (43% versus 28%). At 3 months, the mean cost per participant for NHS resource use for the intervention and control groups were £57 and £102, respectively. At 6-months, the mean cost per participant for NHS resource use for the intervention and control groups were £109 and £49, respectively. Overall costs were similar but marginally higher in the intervention group.<br>**Intervention delivery costs**<br>Costs associated with the intervention were slightly higher (around £81 per dyad) than the control (£72 per dyad) due to factors such as additional staff training required for the intervention.<br>**Effectiveness**<br>Primary outcome measure: intention-to-treat analysis for 84% retained participants (175 intervention; 174 control) found mean (SD) Family Appraisal of |
|  |  |  |  | Caregiving Questionnaire carer strain at 3 months to be 3.11 (0.87) in the control group compared with 3.03 (0.90) in the intervention group, adjusted mean difference of -0.04 (95% CI -0.20 to 0.13). Secondary outcomes had similarly small differences and tight CIs. Sensitivity analyses suggested robust findings.<br>For both the complete case and imputed data there was no statistically significant difference between the treatment groups in terms of QALYs.<br>**Conclusion**<br>The intervention was not fully implemented in this pragmatic trial. As delivered, it conferred no clinical benefits and is unlikely to be cost-effective compared with usual care from a stroke specialist provider organization. It remains unclear how best to support carers of stroke survivors.<br>It is unlikely that the intervention is cost-effective compared to usual care. However, although the results suggest that the intervention was dominated (i.e. due to QALY loss), because the size of the QALY loss was so small, and not statistically significant, it is more<br>appropriate to consider this as no difference in health. An important limitation of the economic analysis is the level of missing economic data, only 131 out of 414 (32%) were included in the adjusted complete case analysis. |
| van der Gaag & Brooks, 2008<br>UK | **Aim**<br>To investigate the feasibility of undertaking economic appraisal in a voluntary sector service providing therapy for people with aphasia and their families.<br>**Intervention type**<br>A 7-week community-based induction (Starter's) programme consisting of group therapy and support for people with stroke and aphasia. The programme focused on improving communication skills and building confidence, and was followed by access to a range of other group-based services including a women's group, "new ways to communicate" group and a conversation group. The therapy groups focus on improving communication skills and training people to teach service providers about developing their communication skills. Counselling is also offered by trained counsellors who themselves live with aphasia.<br>**Intervention deliverer/organization**<br>Connect — the Communication Disability Network — provides services in London and South-West England, with an educational service to health- and social care professionals throughout the UK. Many of the services on offer at Connect are co-facilitated by people with aphasia. | **Outcome**<br>Health Related Quality of Life (HRQoL) measured by EQ-5D<br>**Types of costs measured**<br>Interventions costs and health resource costs, including staff (contact and non-contact), cost of materials, equipment and overheads. Transport time and costs for clients were included to enable comparison with available NHS costings for similar programs. | **Quality rating**<br>JBI critical appraisal checklist for economic evaluations<br>**2/11**<br>Medium quality | **Cost-effectiveness**<br>Cost-effectiveness was not calculated for this feasibility evaluation.<br>**Resource use costs**<br>Connect 7-week Starter's Programme was cheaper (£42 per client per session) than the closest equivalent NHS SLT Programme (£62 per client per session).<br>Transport costs are key. If all transport and travel costs are included for both programmes, the Connect programme remains the less costly at £55 per client per session compared with £74 for NHS, with NHS transport costs 63% of total costs (£4641 out of a total cost of £7413),<br>In some NHS Trusts the cost of transport will be much lower and may in fact be borne by the people with aphasia and their relatives/ carers as they are at Connect.<br>**Intervention delivery costs** |
|  | **Sample size**<br>25 clients completed EQ-5D assessments in the Starter's Programme for the QALY analysis. Sample size for the broader evaluation was not stated.<br>**Length of follow-up**<br>The evaluation of Connect took place over an 18-month period.<br>**Analytical approach (e.g. trial-based or model-based).**<br>Trial based economic evaluation without a control group.<br>**Type of economic evaluation**<br>Feasibility cost-utility analysis (cost-utility ratios such as cost per QALY gained or comparisons were not calculated).<br>**Perspective of analysis**<br>Health care and societal perspective.<br>**Currency and cost year**<br>GBP (£), 2002<br>**Discounting**<br>Not stated<br>**Sensitivity analysis**<br>Sensitivity analysis was not undertaken for this study |  |  | A key cost feature of the intervention is overheads which a 37% of total costs. The main explanation is the high rental Connect pays for its premises. This, and the relatively small number of clients spread over each of the various programmes, leads to relatively high overhead allocations per programme.<br>**Effectiveness**<br>Mean QALY change varied from 0.306 gain to mean -0.37 loss, depending on client background. Overall QALY changes were not clearly reported. These findings should be considered exploratory, particularly due to the small sample (n=25) and lack of subgroup detail (e.g. sizes and statistical analysis).<br>**Conclusion**<br>The full analysis was not fully carried out in this feasibility trial. Although the cost per client per session was cheaper for the intervention than for the NHS, the effectiveness of the two programs is not reported. QALY data are preliminary and not robust enough to draw firm conclusions about impact. |

**Abbreviations:** ADL (Activities of Daily Living); AQoL-4D (Assessment of Quality of Life-Four Dimensions); AUD (Australian Dollars); CB (Caregiver Burden); CCA (Cost-Consequence Analysis); CDM (Chronic Disease Management); CI (Confidence Interval); CSRI (Client Service Receipt Inventory); CUA (Cost Utility Analysis); DISCS (Depression Intensity Scale Circles); EQ-5D-5L (EuroQoL-Five Dimensions-Five Levels); FACQ (Family Appraisal of Caregiving Questionnaire), FAI (Frenchay Activities Index); GHQ-12 (General Health Questionnaire-12); HADS (Hospital Anxiety and Depression Scale); HRQoL (Health Related Quality of Life); ICER (Incremental Cost Effectiveness Ratio); IMP (Individual Management Program); LOS (Length of Stay); LSCTC (London Stroke Carer Training Course); NEADL (Nottingham Extended Activities of Daily Living); NHS (National Health Service); NICE (National Institute for Health and Care Excellence); QALY (Quality Adjusted Life Year); RCT (Randomised Controlled Trial); SCC (Stroke

**Table 4:**
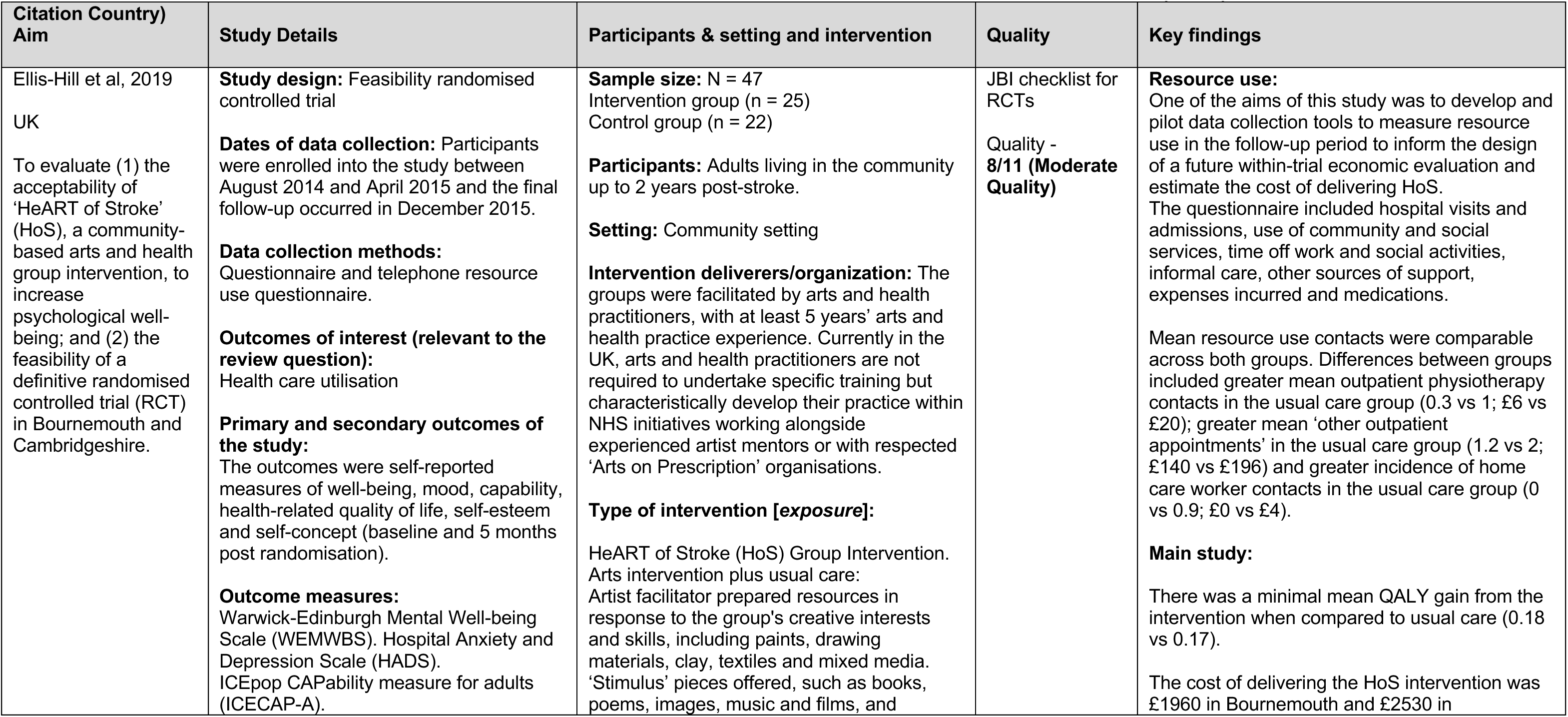
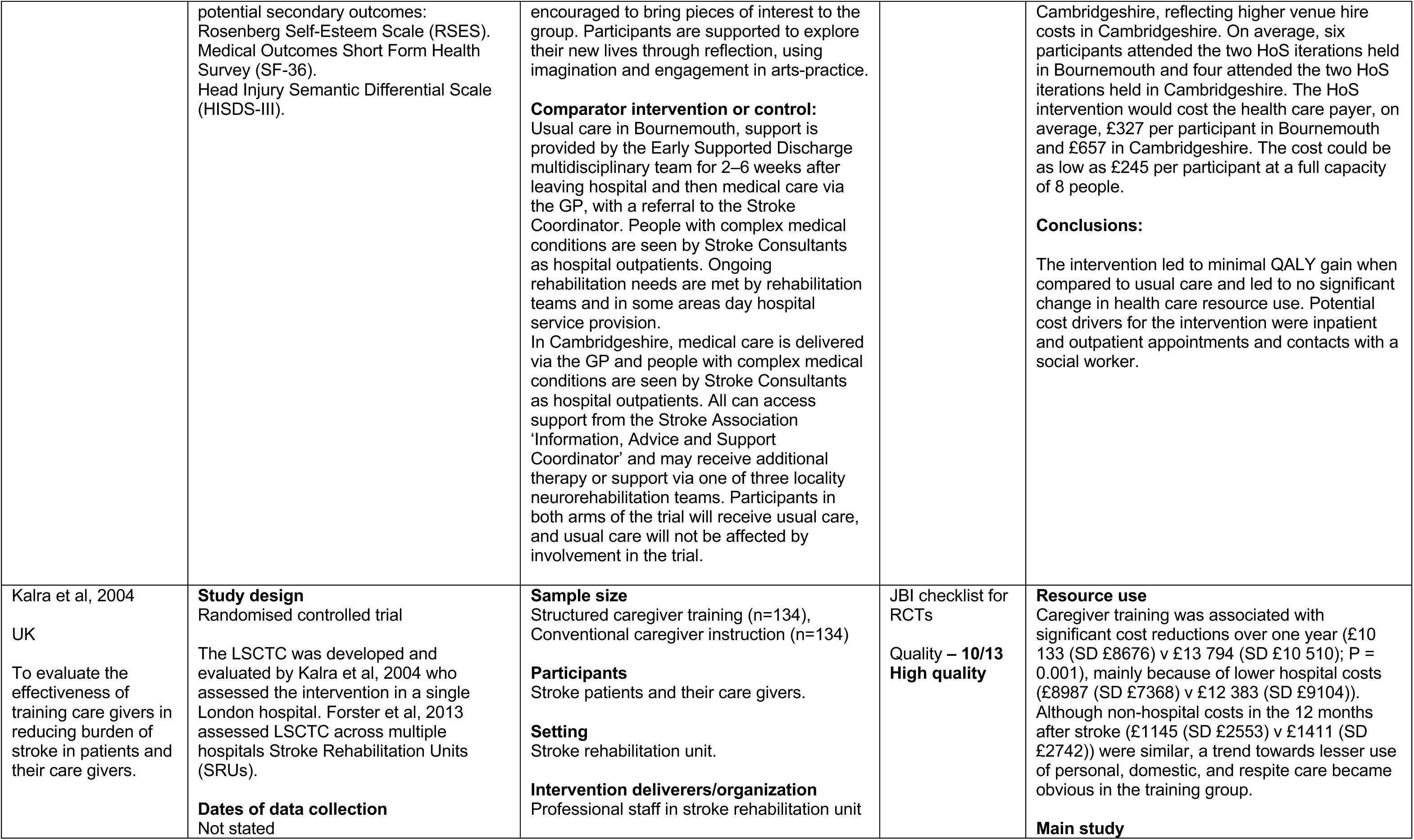
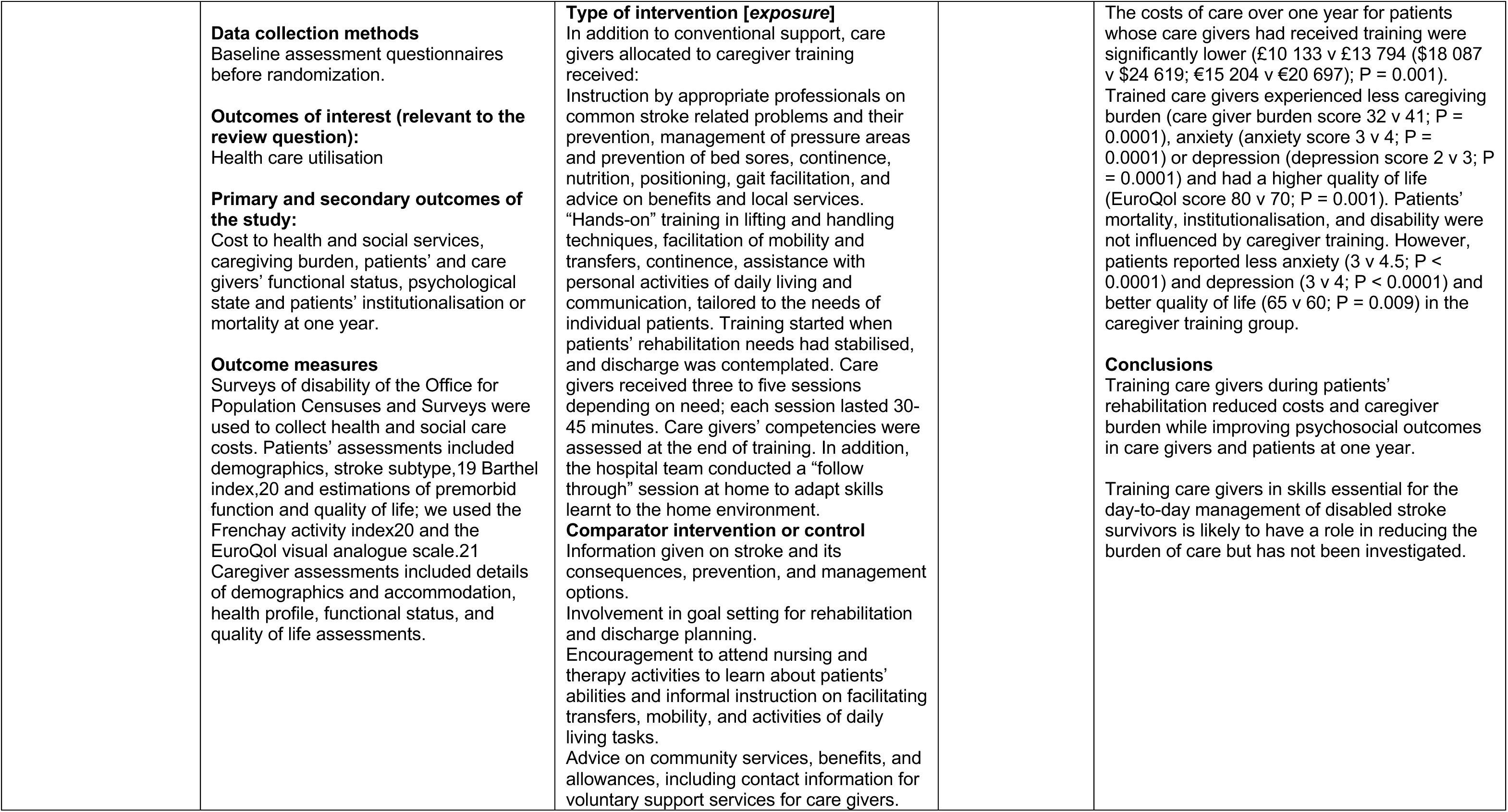
Evidence from randomised controlled trials of life after stroke services with cost comparison (n = 2)

| Citation Country)<br>Aim | Study Details | Participants & setting and intervention | Quality | Key findings |
| --- | --- | --- | --- | --- |
| Ellis-Hill et al, 2019<br>UK<br>To evaluate (1) the acceptability of 'HeART of Stroke' (HoS), a community-based arts and health group intervention, to increase psychological well-being; and (2) the feasibility of a definitive randomised controlled trial (RCT) in Bournemouth and Cambridgeshire. | **Study design:** Feasibility randomised controlled trial<br>**Dates of data collection:** Participants were enrolled into the study between August 2014 and April 2015 and the final follow-up occurred in December 2015.<br>**Data collection methods:** Questionnaire and telephone resource use questionnaire.<br>**Outcomes of interest (relevant to the review question):** Health care utilisation<br>**Primary and secondary outcomes of the study:** The outcomes were self-reported measures of well-being, mood, capability, health-related quality of life, self-esteem and self-concept (baseline and 5 months post randomisation).<br>**Outcome measures:** Warwick-Edinburgh Mental Well-being Scale (WEMWBS). Hospital Anxiety and Depression Scale (HADS). ICEpop CAPability measure for adults (ICECAP-A). | **Sample size:** N = 47<br>Intervention group (n = 25)<br>Control group (n = 22)<br>**Participants:** Adults living in the community up to 2 years post-stroke.<br>**Setting:** Community setting<br>**Intervention deliverers/organization:** The groups were facilitated by arts and health practitioners, with at least 5 years' arts and health practice experience. Currently in the UK, arts and health practitioners are not required to undertake specific training but characteristically develop their practice within NHS initiatives working alongside experienced artist mentors or with respected 'Arts on Prescription' organisations.<br>**Type of intervention [exposure]:**<br>HeART of Stroke (HoS) Group Intervention. Arts intervention plus usual care: Artist facilitator prepared resources in response to the group's creative interests and skills, including paints, drawing materials, clay, textiles and mixed media. 'Stimulus' pieces offered, such as books, poems, images, music and films, and | JB1 checklist for RCTs<br>Quality - **8/11 (Moderate Quality)** | **Resource use:**<br>One of the aims of this study was to develop and pilot data collection tools to measure resource use in the follow-up period to inform the design of a future within-trial economic evaluation and estimate the cost of delivering HoS. The questionnaire included hospital visits and admissions, use of community and social services, time off work and social activities, informal care, other sources of support, expenses incurred and medications.<br>Mean resource use contacts were comparable across both groups. Differences between groups included greater mean outpatient physiotherapy contacts in the usual care group (0.3 vs 1; £6 vs £20); greater mean 'other outpatient appointments' in the usual care group (1.2 vs 2; £140 vs £196) and greater incidence of home care worker contacts in the usual care group (0 vs 0.9; £0 vs £4).<br>**Main study:**<br>There was a minimal mean QALY gain from the intervention when compared to usual care (0.18 vs 0.17).<br>The cost of delivering the HoS intervention was £1960 in Bournemouth and £2530 in |
|  | potential secondary outcomes: Rosenberg Self-Esteem Scale (RSES). Medical Outcomes Short Form Health Survey (SF-36). Head Injury Semantic Differential Scale (HISDS-III). | encouraged to bring pieces of interest to the group. Participants are supported to explore their new lives through reflection, using imagination and engagement in arts-practice.<br>**Comparator intervention or control:**<br>Usual care in Bournemouth, support is provided by the Early Supported Discharge multidisciplinary team for 2–6 weeks after leaving hospital and then medical care via the GP, with a referral to the Stroke Coordinator. People with complex medical conditions are seen by Stroke Consultants as hospital outpatients. Ongoing rehabilitation needs are met by rehabilitation teams and in some areas day hospital service provision.<br>In Cambridgeshire, medical care is delivered via the GP and people with complex medical conditions are seen by Stroke Consultants as hospital outpatients. All can access support from the Stroke Association 'Information, Advice and Support Coordinator' and may receive additional therapy or support via one of three locality neurorehabilitation teams. Participants in both arms of the trial will receive usual care, and usual care will not be affected by involvement in the trial. |  | Cambridgeshire, reflecting higher venue hire costs in Cambridgeshire. On average, six participants attended the two HoS iterations held in Bournemouth and four attended the two HoS iterations held in Cambridgeshire. The HoS intervention would cost the health care payer, on average, £327 per participant in Bournemouth and £657 in Cambridgeshire. The cost could be as low as £245 per participant at a full capacity of 8 people.<br>**Conclusions:**<br>The intervention led to minimal QALY gain when compared to usual care and led to no significant change in health care resource use. Potential cost drivers for the intervention were inpatient and outpatient appointments and contacts with a social worker. |
| Kalra et al, 2004<br>UK<br>To evaluate the effectiveness of training care givers in reducing burden of stroke in patients and their care givers. | **Study design**<br>Randomised controlled trial<br>The LSCTC was developed and evaluated by Kalra et al, 2004 who assessed the intervention in a single London hospital. Forster et al, 2013 assessed LSCTC across multiple hospitals Stroke Rehabilitation Units (SRUs).<br>**Dates of data collection**<br>Not stated | **Sample size**<br>Structured caregiver training (n=134), Conventional caregiver instruction (n=134)<br>**Participants**<br>Stroke patients and their care givers.<br>**Setting**<br>Stroke rehabilitation unit.<br>**Intervention deliverers/organization**<br>Professional staff in stroke rehabilitation unit | JB1 checklist for RCTs<br>**Quality – 10/13**<br>**High quality** | **Resource use**<br>Caregiver training was associated with significant cost reductions over one year (£10 133 (SD £8676) v £13 794 (SD £10 510); P = 0.001), mainly because of lower hospital costs (£8987 (SD £7368) v £12 383 (SD £9104)). Although non-hospital costs in the 12 months after stroke (£1145 (SD £2553) v £1411 (SD £2742)) were similar, a trend towards lesser use of personal, domestic, and respite care became obvious in the training group.<br>**Main study** |

|  |  |  |  |
| --- | --- | --- | --- |
| | **Data collection methods**<br>Baseline assessment questionnaires before randomization.<br>**Outcomes of interest (relevant to the review question):**<br>Health care utilisation<br>**Primary and secondary outcomes of the study:**<br>Cost to health and social services, caregiving burden, patients' and care givers' functional status, psychological state and patients' institutionalisation or mortality at one year.<br>**Outcome measures**<br>Surveys of disability of the Office for Population Censuses and Surveys were used to collect health and social care costs. Patients' assessments included demographics, stroke subtype,<sup>19</sup> Barthel index,<sup>20</sup> and estimations of premorbid function and quality of life; we used the Frenchay activity index<sup>20</sup> and the EuroQol visual analogue scale.<sup>21</sup> Caregiver assessments included details of demographics and accommodation, health profile, functional status, and quality of life assessments. | **Type of intervention [exposure]**<br>In addition to conventional support, care givers allocated to caregiver training received:<br>Instruction by appropriate professionals on common stroke related problems and their prevention, management of pressure areas and prevention of bed sores, continence, nutrition, positioning, gait facilitation, and advice on benefits and local services. "Hands-on" training in lifting and handling techniques, facilitation of mobility and transfers, continence, assistance with personal activities of daily living and communication, tailored to the needs of individual patients. Training started when patients' rehabilitation needs had stabilised, and discharge was contemplated. Care givers received three to five sessions depending on need; each session lasted 30-45 minutes. Care givers' competencies were assessed at the end of training. In addition, the hospital team conducted a "follow through" session at home to adapt skills learnt to the home environment.<br>**Comparator intervention or control**<br>Information given on stroke and its consequences, prevention, and management options.<br>Involvement in goal setting for rehabilitation and discharge planning.<br>Encouragement to attend nursing and therapy activities to learn about patients' abilities and informal instruction on facilitating transfers, mobility, and activities of daily living tasks.<br>Advice on community services, benefits, and allowances, including contact information for voluntary support services for care givers. | The costs of care over one year for patients whose care givers had received training were significantly lower (£10 133 v £13 794 (\$18 087 v \$24 619; €15 204 v €20 697); P = 0.001). Trained care givers experienced less caregiving burden (care giver burden score 32 v 41; P = 0.0001), anxiety (anxiety score 3 v 4; P = 0.0001) or depression (depression score 2 v 3; P = 0.0001) and had a higher quality of life (EuroQol score 80 v 70; P = 0.001). Patients' mortality, institutionalisation, and disability were not influenced by caregiver training. However, patients reported less anxiety (3 v 4.5; P < 0.0001) and depression (3 v 4; P < 0.0001) and better quality of life (65 v 60; P = 0.009) in the caregiver training group.<br>**Conclusions**<br>Training care givers during patients' rehabilitation reduced costs and caregiver burden while improving psychosocial outcomes in care givers and patients at one year.<br>Training care givers in skills essential for the day-to-day management of disabled stroke survivors is likely to have a role in reducing the burden of care but has not been investigated. |

**Table 5:**
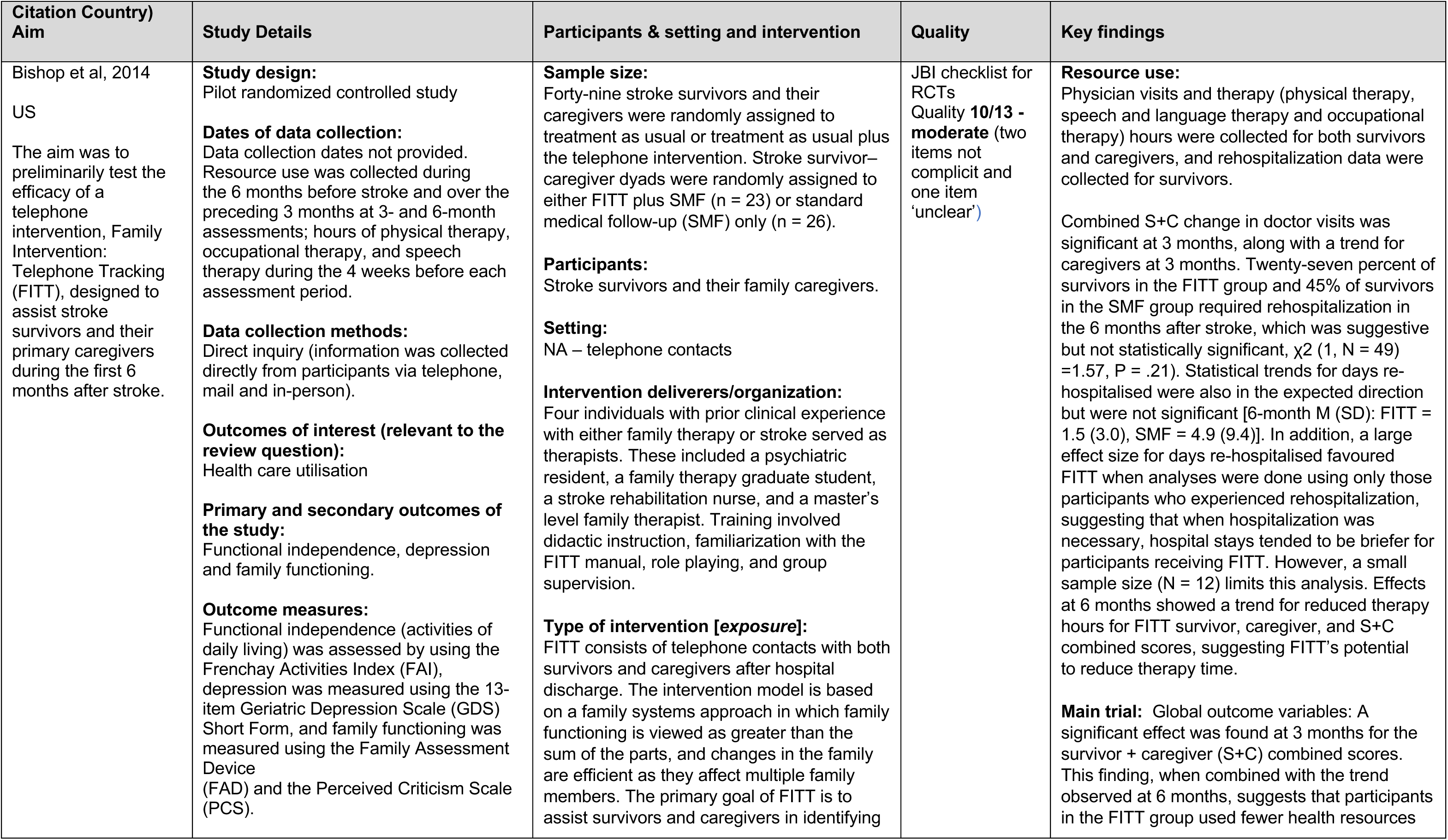
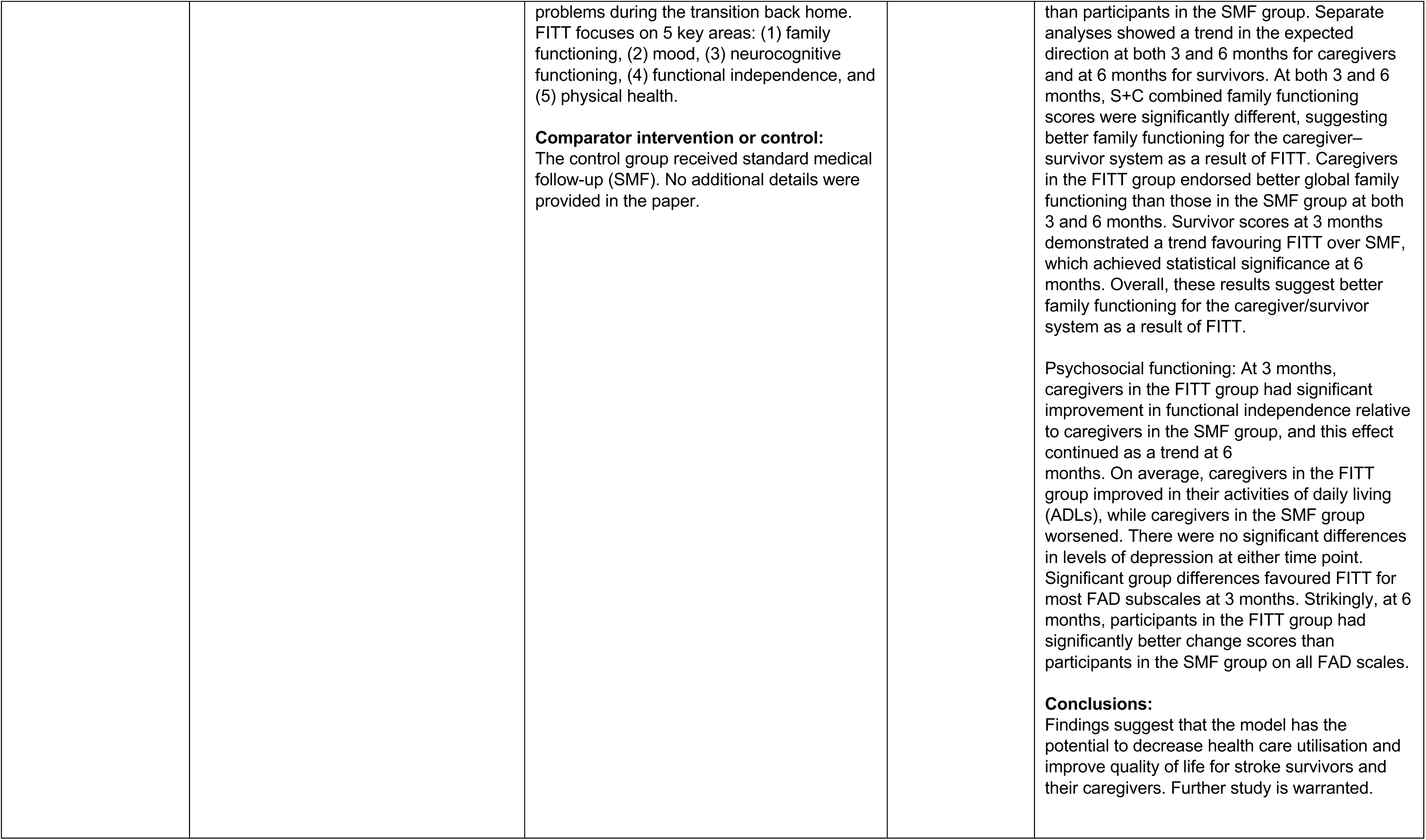
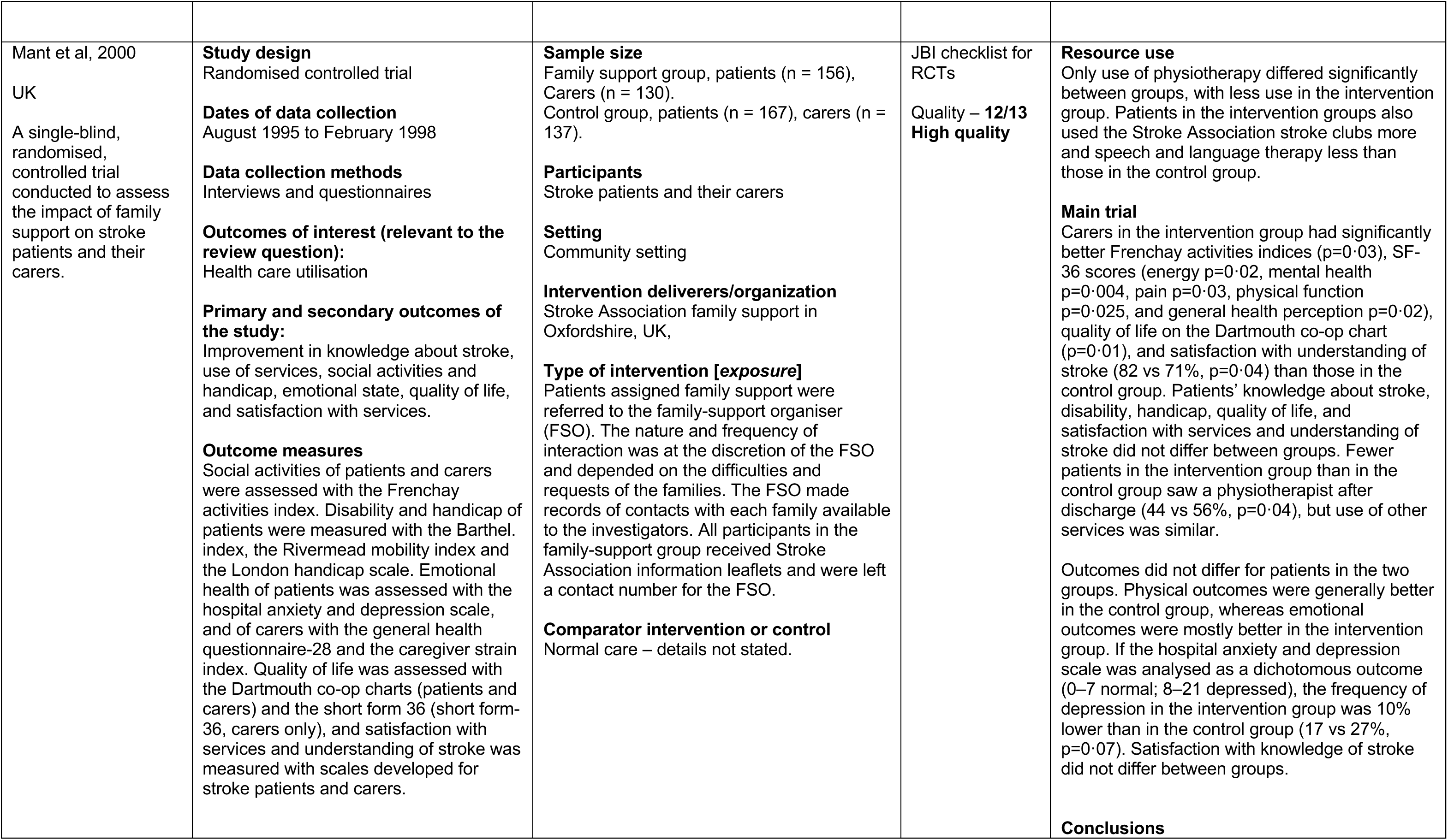
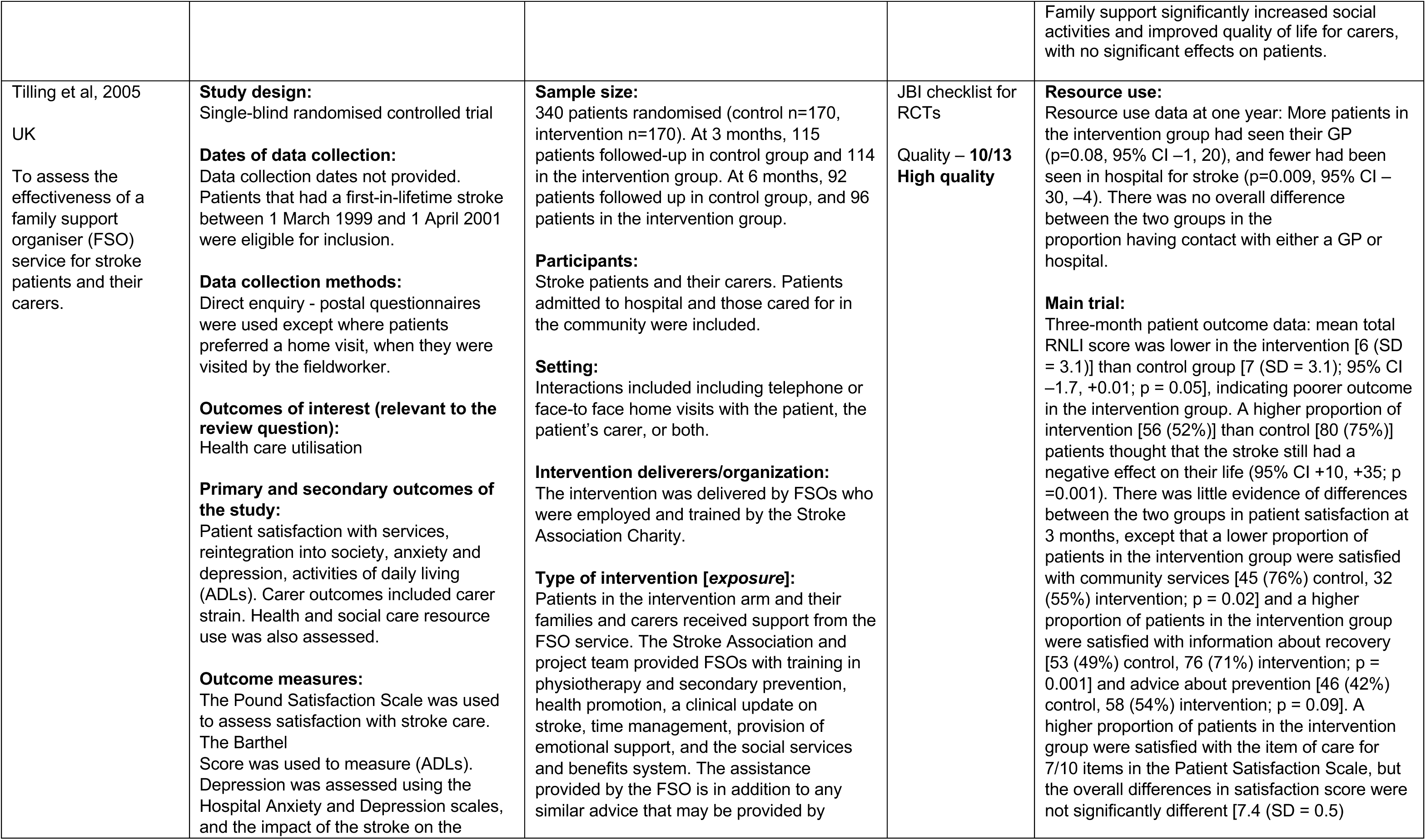
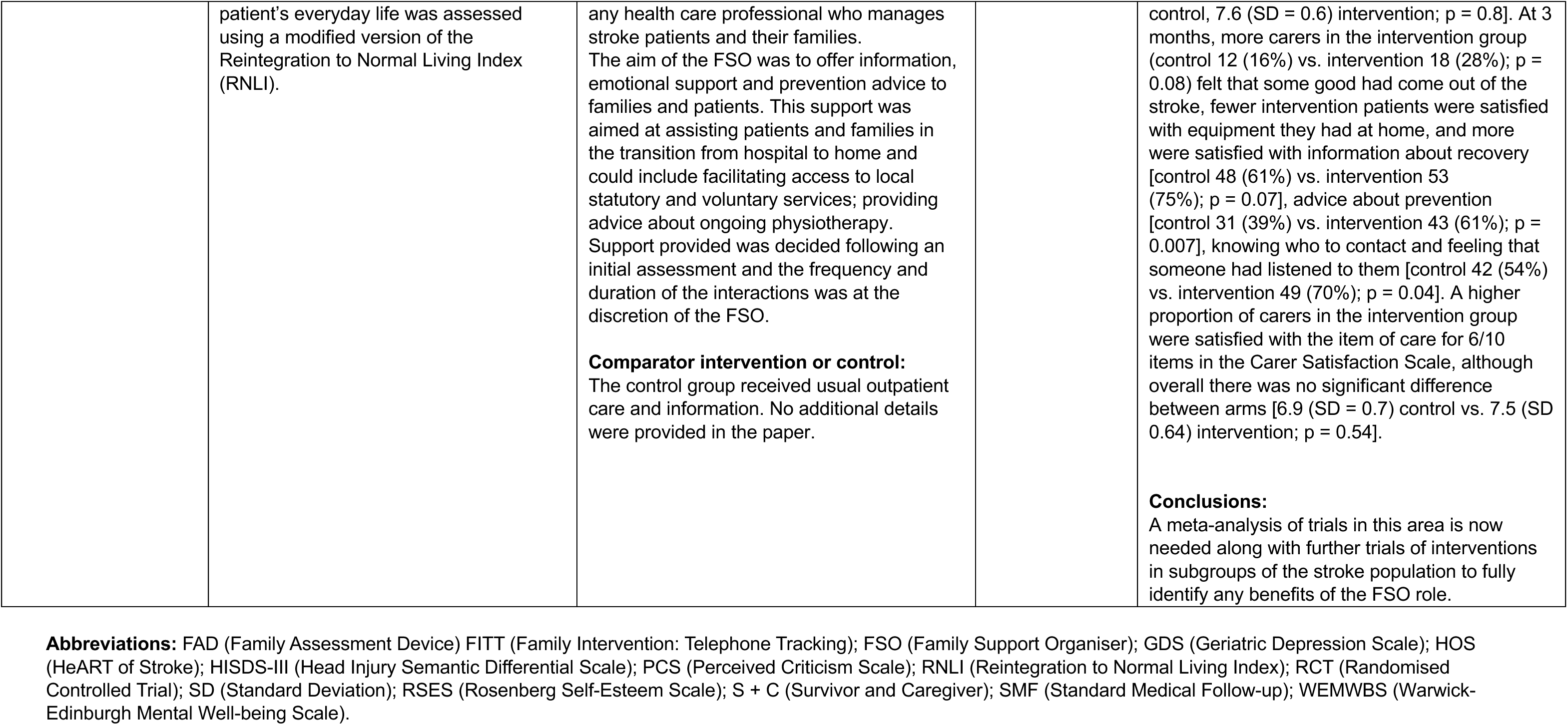
Evidence from randomised controlled trials of life after stroke services (n = 3)

| Citation Country)<br>Aim | Study Details | Participants & setting and intervention | Quality | Key findings |
| --- | --- | --- | --- | --- |
| Bishop et al, 2014<br>US<br>The aim was to preliminarily test the efficacy of a telephone intervention, Family Intervention: Telephone Tracking (FITT), designed to assist stroke survivors and their primary caregivers during the first 6 months after stroke. | **Study design:**<br>Pilot randomized controlled study<br>**Dates of data collection:**<br>Data collection dates not provided. Resource use was collected during the 6 months before stroke and over the preceding 3 months at 3- and 6-month assessments; hours of physical therapy, occupational therapy, and speech therapy during the 4 weeks before each assessment period.<br>**Data collection methods:**<br>Direct inquiry (information was collected directly from participants via telephone, mail and in-person).<br>**Outcomes of interest (relevant to the review question):**<br>Health care utilisation<br>**Primary and secondary outcomes of the study:**<br>Functional independence, depression and family functioning.<br>**Outcome measures:**<br>Functional independence (activities of daily living) was assessed by using the Frenchay Activities Index (FAI), depression was measured using the 13-item Geriatric Depression Scale (GDS) Short Form, and family functioning was measured using the Family Assessment Device (FAD) and the Perceived Criticism Scale (PCS). | **Sample size:**<br>Forty-nine stroke survivors and their caregivers were randomly assigned to treatment as usual or treatment as usual plus the telephone intervention. Stroke survivor–caregiver dyads were randomly assigned to either FITT plus SMF (n = 23) or standard medical follow-up (SMF) only (n = 26).<br>**Participants:**<br>Stroke survivors and their family caregivers.<br>**Setting:**<br>NA – telephone contacts<br>**Intervention deliverers/organization:**<br>Four individuals with prior clinical experience with either family therapy or stroke served as therapists. These included a psychiatric resident, a family therapy graduate student, a stroke rehabilitation nurse, and a master's level family therapist. Training involved didactic instruction, familiarization with the FITT manual, role playing, and group supervision.<br>**Type of intervention [exposure]:**<br>FITT consists of telephone contacts with both survivors and caregivers after hospital discharge. The intervention model is based on a family systems approach in which family functioning is viewed as greater than the sum of the parts, and changes in the family are efficient as they affect multiple family members. The primary goal of FITT is to assist survivors and caregivers in identifying | JB1 checklist for RCTs<br>Quality **10/13 - moderate** (two items not complicit and one item 'unclear') | **Resource use:**<br>Physician visits and therapy (physical therapy, speech and language therapy and occupational therapy) hours were collected for both survivors and caregivers, and rehospitalization data were collected for survivors.<br>Combined S+C change in doctor visits was significant at 3 months, along with a trend for caregivers at 3 months. Twenty-seven percent of survivors in the FITT group and 45% of survivors in the SMF group required rehospitalization in the 6 months after stroke, which was suggestive but not statistically significant, $\chi^2$ (1, N = 49) = 1.57, P = .21). Statistical trends for days re-hospitalised were also in the expected direction but were not significant [6-month M (SD): FITT = 1.5 (3.0), SMF = 4.9 (9.4)]. In addition, a large effect size for days re-hospitalised favoured FITT when analyses were done using only those participants who experienced rehospitalization, suggesting that when hospitalization was necessary, hospital stays tended to be briefer for participants receiving FITT. However, a small sample size (N = 12) limits this analysis. Effects at 6 months showed a trend for reduced therapy hours for FITT survivor, caregiver, and S+C combined scores, suggesting FITT's potential to reduce therapy time.<br>**Main trial:** Global outcome variables: A significant effect was found at 3 months for the survivor + caregiver (S+C) combined scores. This finding, when combined with the trend observed at 6 months, suggests that participants in the FITT group used fewer health resources |
|  |  | problems during the transition back home. FITT focuses on 5 key areas: (1) family functioning, (2) mood, (3) neurocognitive functioning, (4) functional independence, and (5) physical health.<br>**Comparator intervention or control:**<br>The control group received standard medical follow-up (SMF). No additional details were provided in the paper. |  | than participants in the SMF group. Separate analyses showed a trend in the expected direction at both 3 and 6 months for caregivers and at 6 months for survivors. At both 3 and 6 months, S+C combined family functioning scores were significantly different, suggesting better family functioning for the caregiver-survivor system as a result of FITT. Caregivers in the FITT group endorsed better global family functioning than those in the SMF group at both 3 and 6 months. Survivor scores at 3 months demonstrated a trend favouring FITT over SMF, which achieved statistical significance at 6 months. Overall, these results suggest better family functioning for the caregiver/survivor system as a result of FITT.<br>Psychosocial functioning: At 3 months, caregivers in the FITT group had significant improvement in functional independence relative to caregivers in the SMF group, and this effect continued as a trend at 6 months. On average, caregivers in the FITT group improved in their activities of daily living (ADLs), while caregivers in the SMF group worsened. There were no significant differences in levels of depression at either time point. Significant group differences favoured FITT for most FAD subscales at 3 months. Strikingly, at 6 months, participants in the FITT group had significantly better change scores than participants in the SMF group on all FAD scales.<br>**Conclusions:**<br>Findings suggest that the model has the potential to decrease health care utilisation and improve quality of life for stroke survivors and their caregivers. Further study is warranted. |
| Mant et al, 2000<br>UK<br>A single-blind, randomised, controlled trial conducted to assess the impact of family support on stroke patients and their carers. | **Study design**<br>Randomised controlled trial<br>**Dates of data collection**<br>August 1995 to February 1998<br>**Data collection methods**<br>Interviews and questionnaires<br>**Outcomes of interest (relevant to the review question):**<br>Health care utilisation<br>**Primary and secondary outcomes of the study:**<br>Improvement in knowledge about stroke, use of services, social activities and handicap, emotional state, quality of life, and satisfaction with services.<br>**Outcome measures**<br>Social activities of patients and carers were assessed with the Frenchay activities index. Disability and handicap of patients were measured with the Barthel index, the Rivermead mobility index and the London handicap scale. Emotional health of patients was assessed with the hospital anxiety and depression scale, and of carers with the general health questionnaire-28 and the caregiver strain index. Quality of life was assessed with the Dartmouth co-op charts (patients and carers) and the short form 36 (short form-36, carers only), and satisfaction with services and understanding of stroke was measured with scales developed for stroke patients and carers. | **Sample size**<br>Family support group, patients (n = 156), Carers (n = 130).<br>Control group, patients (n = 167), carers (n = 137).<br>**Participants**<br>Stroke patients and their carers<br>**Setting**<br>Community setting<br>**Intervention deliverers/organization**<br>Stroke Association family support in Oxfordshire, UK,<br>**Type of intervention [exposure]**<br>Patients assigned family support were referred to the family-support organiser (FSO). The nature and frequency of interaction was at the discretion of the FSO and depended on the difficulties and requests of the families. The FSO made records of contacts with each family available to the investigators. All participants in the family-support group received Stroke Association information leaflets and were left a contact number for the FSO.<br>**Comparator intervention or control**<br>Normal care – details not stated. | JB1 checklist for RCTs<br>Quality – **12/13**<br>**High quality** | **Resource use**<br>Only use of physiotherapy differed significantly between groups, with less use in the intervention group. Patients in the intervention groups also used the Stroke Association stroke clubs more and speech and language therapy less than those in the control group.<br>**Main trial**<br>Carers in the intervention group had significantly better Frenchay activities indices (p=0.03), SF-36 scores (energy p=0.02, mental health p=0.004, pain p=0.03, physical function p=0.025, and general health perception p=0.02), quality of life on the Dartmouth co-op chart (p=0.01), and satisfaction with understanding of stroke (82 vs 71%, p=0.04) than those in the control group. Patients' knowledge about stroke, disability, handicap, quality of life, and satisfaction with services and understanding of stroke did not differ between groups. Fewer patients in the intervention group than in the control group saw a physiotherapist after discharge (44 vs 56%, p=0.04), but use of other services was similar.<br>Outcomes did not differ for patients in the two groups. Physical outcomes were generally better in the control group, whereas emotional outcomes were mostly better in the intervention group. If the hospital anxiety and depression scale was analysed as a dichotomous outcome (0–7 normal; 8–21 depressed), the frequency of depression in the intervention group was 10% lower than in the control group (17 vs 27%, p=0.07). Satisfaction with knowledge of stroke did not differ between groups.<br>**Conclusions** |
|  |  |  |  | Family support significantly increased social activities and improved quality of life for carers, with no significant effects on patients. |
| Tilling et al, 2005<br>UK<br>To assess the effectiveness of a family support organiser (FSO) service for stroke patients and their carers. | **Study design:**<br>Single-blind randomised controlled trial<br>**Dates of data collection:**<br>Data collection dates not provided. Patients that had a first-in-lifetime stroke between 1 March 1999 and 1 April 2001 were eligible for inclusion.<br>**Data collection methods:**<br>Direct enquiry - postal questionnaires were used except where patients preferred a home visit, when they were visited by the fieldworker.<br>**Outcomes of interest (relevant to the review question):**<br>Health care utilisation<br>**Primary and secondary outcomes of the study:**<br>Patient satisfaction with services, reintegration into society, anxiety and depression, activities of daily living (ADLs). Carer outcomes included carer strain. Health and social care resource use was also assessed.<br>**Outcome measures:**<br>The Pound Satisfaction Scale was used to assess satisfaction with stroke care. The Barthel Score was used to measure (ADLs). Depression was assessed using the Hospital Anxiety and Depression scales, and the impact of the stroke on the | **Sample size:**<br>340 patients randomised (control n=170, intervention n=170). At 3 months, 115 patients followed-up in control group and 114 in the intervention group. At 6 months, 92 patients followed up in control group, and 96 patients in the intervention group.<br>**Participants:**<br>Stroke patients and their carers. Patients admitted to hospital and those cared for in the community were included.<br>**Setting:**<br>Interactions included including telephone or face-to face home visits with the patient, the patient's carer, or both.<br>**Intervention deliverers/organization:**<br>The intervention was delivered by FSOs who were employed and trained by the Stroke Association Charity.<br>**Type of intervention [exposure]:**<br>Patients in the intervention arm and their families and carers received support from the FSO service. The Stroke Association and project team provided FSOs with training in physiotherapy and secondary prevention, health promotion, a clinical update on stroke, time management, provision of emotional support, and the social services and benefits system. The assistance provided by the FSO is in addition to any similar advice that may be provided by | JB1 checklist for RCTs<br>Quality – **10/13**<br>**High quality** | **Resource use:**<br>Resource use data at one year: More patients in the intervention group had seen their GP ($p=0.08$, 95% CI $-1$, $20$), and fewer had been seen in hospital for stroke ($p=0.009$, 95% CI $-30$, $-4$). There was no overall difference between the two groups in the proportion having contact with either a GP or hospital.<br>**Main trial:**<br>Three-month patient outcome data: mean total RNLI score was lower in the intervention [6 (SD = 3.1)] than control group [7 (SD = 3.1); 95% CI $-1.7$, $+0.01$; $p = 0.05$], indicating poorer outcome in the intervention group. A higher proportion of intervention [56 (52%)] than control [80 (75%)] patients thought that the stroke still had a negative effect on their life (95% CI $+10$, $+35$; $p = 0.001$). There was little evidence of differences between the two groups in patient satisfaction at 3 months, except that a lower proportion of patients in the intervention group were satisfied with community services [45 (76%) control, 32 (55%) intervention; $p = 0.02$] and a higher proportion of patients in the intervention group were satisfied with information about recovery [53 (49%) control, 76 (71%) intervention; $p = 0.001$] and advice about prevention [46 (42%) control, 58 (54%) intervention; $p = 0.09$]. A higher proportion of patients in the intervention group were satisfied with the item of care for 7/10 items in the Patient Satisfaction Scale, but the overall differences in satisfaction score were not significantly different [7.4 (SD = 0.5)] |

|  |  |  |  |
| --- | --- | --- | --- |
|  | patient's everyday life was assessed using a modified version of the Reintegration to Normal Living Index (RNLI). | any health care professional who manages stroke patients and their families. The aim of the FSO was to offer information, emotional support and prevention advice to families and patients. This support was aimed at assisting patients and families in the transition from hospital to home and could include facilitating access to local statutory and voluntary services; providing advice about ongoing physiotherapy. Support provided was decided following an initial assessment and the frequency and duration of the interactions was at the discretion of the FSO.<br>**Comparator intervention or control:**<br>The control group received usual outpatient care and information. No additional details were provided in the paper. | control, 7.6 (SD = 0.6) intervention; $p = 0.8$]. At 3 months, more carers in the intervention group (control 12 (16%) vs. intervention 18 (28%); $p = 0.08$) felt that some good had come out of the stroke, fewer intervention patients were satisfied with equipment they had at home, and more were satisfied with information about recovery [control 48 (61%) vs. intervention 53 (75%); $p = 0.07$], advice about prevention [control 31 (39%) vs. intervention 43 (61%); $p = 0.007$], knowing who to contact and feeling that someone had listened to them [control 42 (54%) vs. intervention 49 (70%); $p = 0.04$]. A higher proportion of carers in the intervention group were satisfied with the item of care for 6/10 items in the Carer Satisfaction Scale, although overall there was no significant difference between arms [6.9 (SD = 0.7) control vs. 7.5 (SD 0.64) intervention; $p = 0.54$].<br>**Conclusions:**<br>A meta-analysis of trials in this area is now needed along with further trials of interventions in subgroups of the stroke population to fully identify any benefits of the FSO role. |

**Abbreviations:** FAD (Family Assessment Device) FITT (Family Intervention: Telephone Tracking); FSO (Family Support Organiser); GDS (Geriatric Depression Scale); HOS (HeART of Stroke); HISDS-III (Head Injury Semantic Differential Scale); PCS (Perceived Criticism Scale); RNLI (Reintegration to Normal Living Index); RCT (Randomised Controlled Trial); SD (Standard Deviation); RSES (Rosenberg Self-Esteem Scale); S + C (Survivor and Caregiver); SMF (Standard Medical Follow-up); WEMWBS (Warwick-Edinburgh Mental Well-being Scale).

### 6.3 Quality appraisal

**Table 6:**
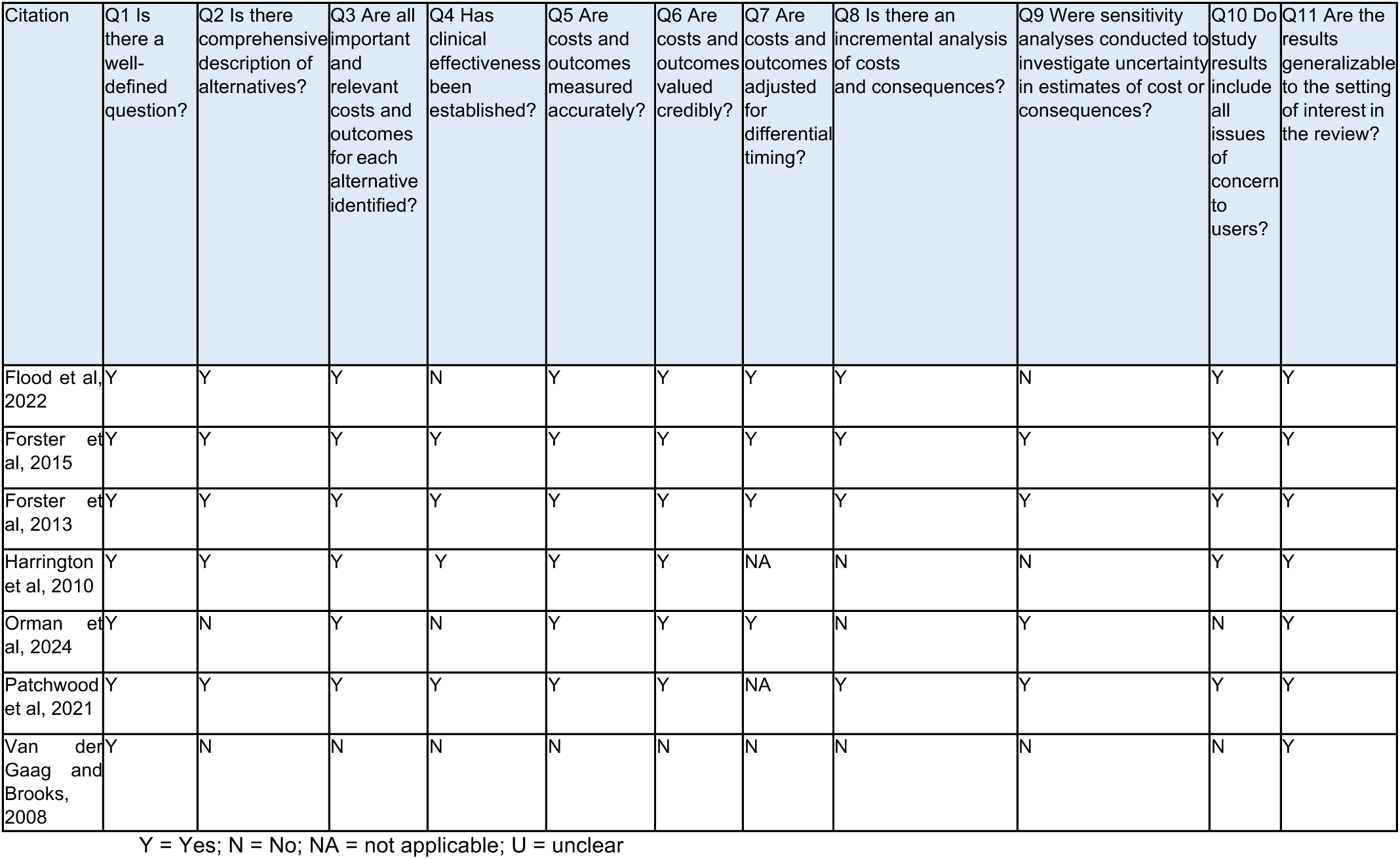
JBI Critical appraisal checklist for economic evaluations.

**Table 7:**
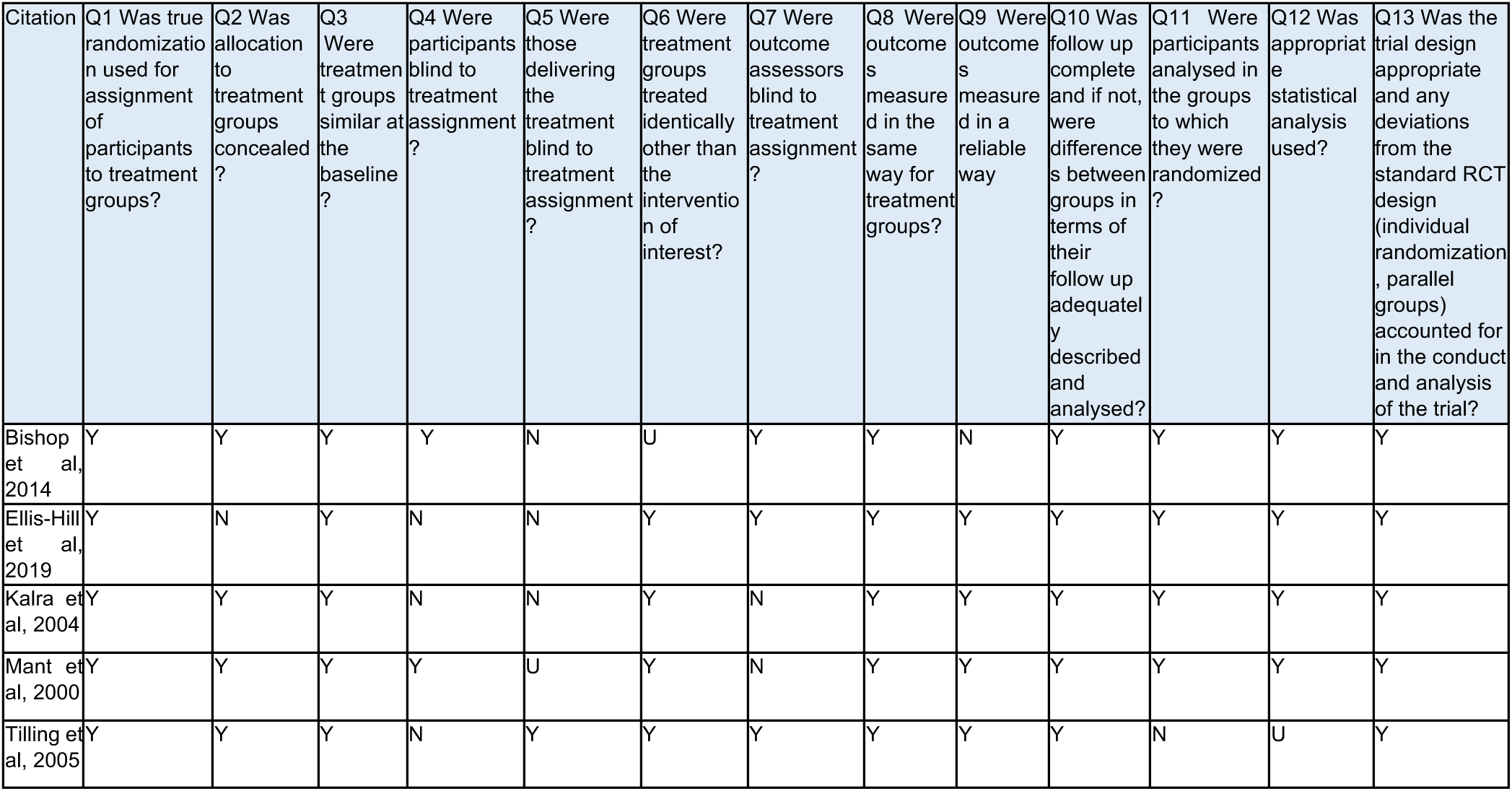
JBI critical appraisal checklist for randomised controlled trials.

## Acknowledgements

The authors would like to thank Angela Contestabile, Katie Chapelle, Josie Hoskins, Heidi James, Robert Hall, Jonathan Hewitt, Llinos Wyn Parry, Lynn Preece, and Niki Turner for their time, expertise, and contributions during stakeholder meetings in guiding the focus of the review and interpretation of findings. We would also like to thank Elizabeth Gillen and Juliet Hounsome, Information Specialists at Cardiff University, for their expert contribution to the search strategy development and database searches. Finally, we would like to thank Catherine Lawrence for invaluable critical feedback on drafts of this review.

## 8 APPENDIX

**APPENDIX 1:**
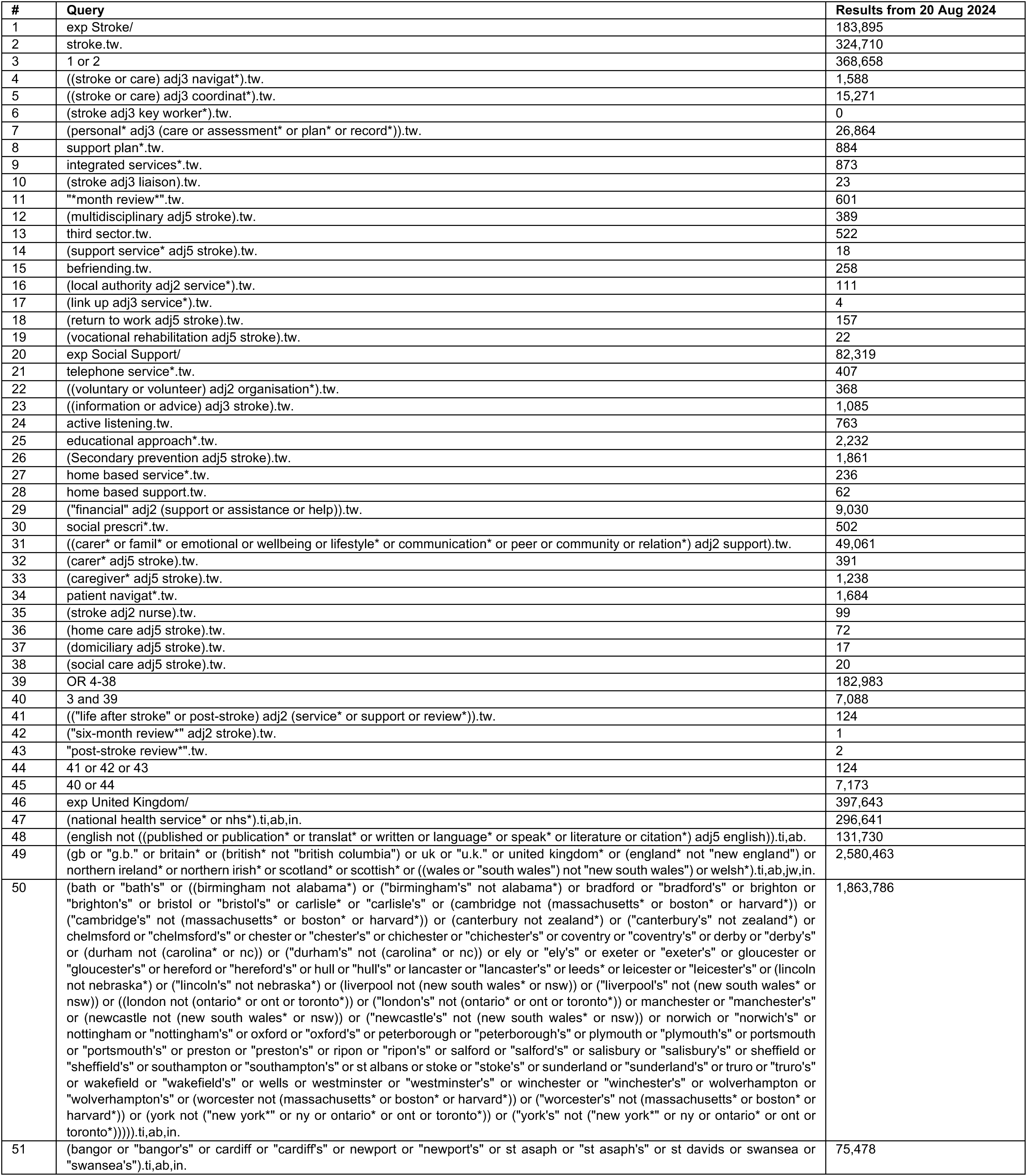
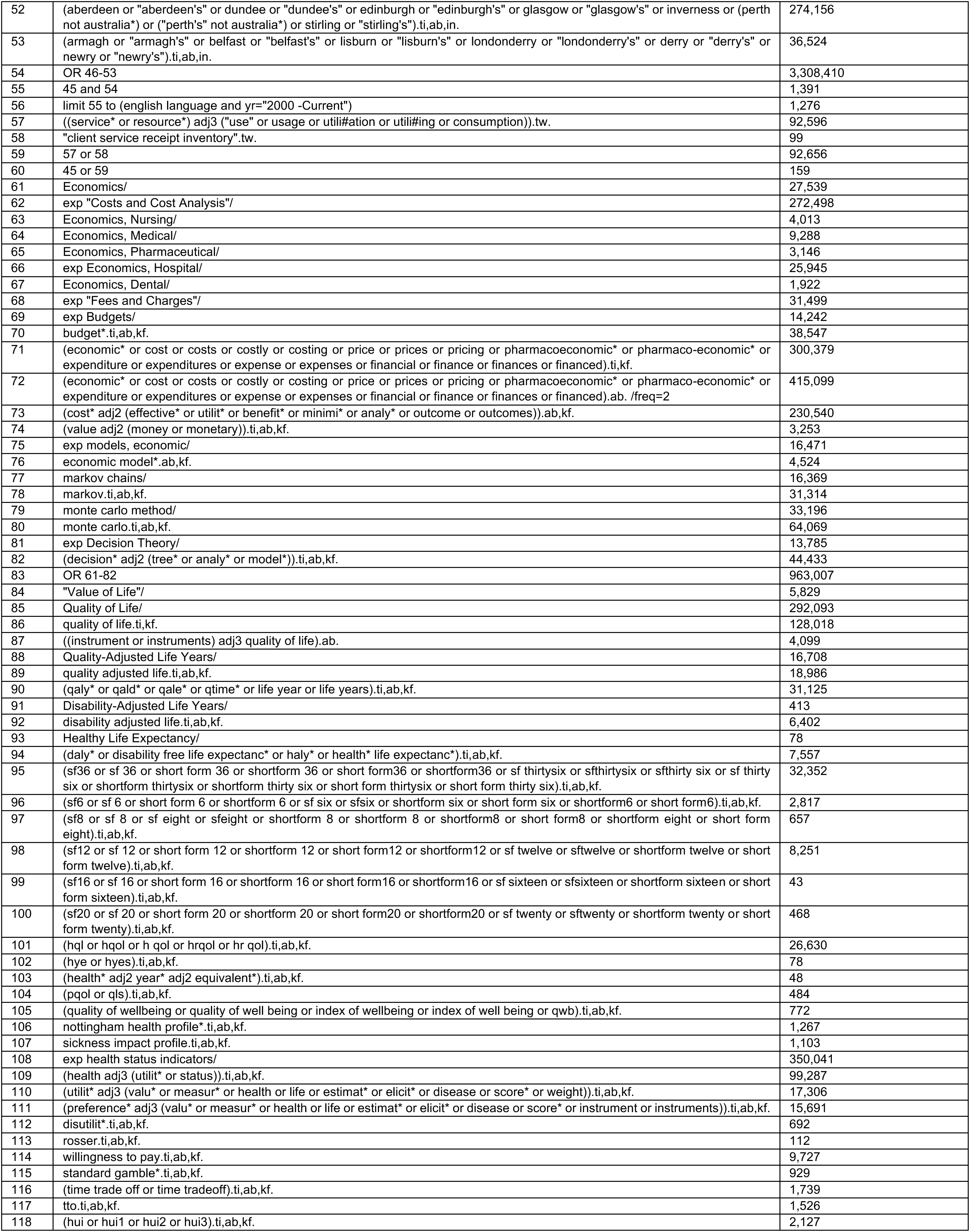
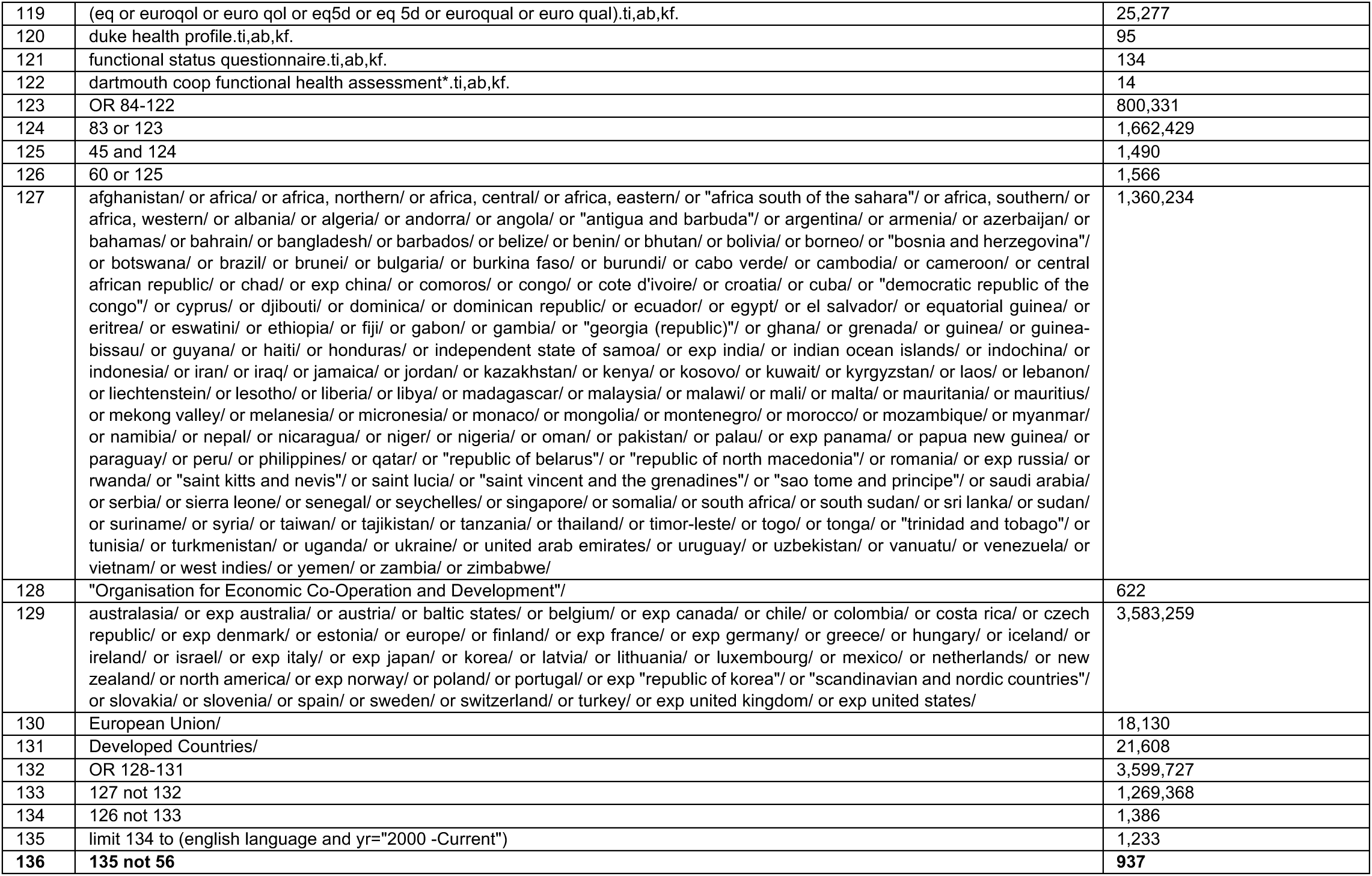
search strategy for Medline via Ovid.

### The Health and Care Research Wales Evidence Centre

Our dedicated team works together with Welsh Government, the NHS, social care, research institutions and the public to deliver vital research to tackle health and social care challenges facing Wales.

Funded by Welsh Government, through Health and Care Research Wales, the Evidence Centre answers key questions to improve health and social care policy and provision across Wales.

Along with our collaborating partners, we conduct reviews of existing evidence and new research, to inform policy and practice needs, with a focus on ensuring real-world impact and public benefit that reaches everyone.

Director: Professor Adrian Edwards

Associate Directors: Dr Alison Cooper, Dr Natalie Joseph-Williams, Dr Ruth Lewis

